# Exploring selection bias in COVID-19 research: Simulations and prospective analyses of two UK cohort studies

**DOI:** 10.1101/2021.12.10.21267363

**Authors:** Louise AC Millard, Alba Fernández-Sanlés, Alice R Carter, Rachael Hughes, Kate Tilling, Tim P Morris, Daniel Major-Smith, Gareth J Griffith, Gemma L Clayton, Emily Kawabata, George Davey Smith, Deborah A Lawlor, Maria Carolina Borges

## Abstract

**Background:** Non-random selection into analytic subsamples could introduce selection bias in observational studies of SARS-CoV-2 infection and COVID-19 severity (e.g. including only those have had a COVID-19 PCR test). We explored the potential presence and impact of selection in such studies using data from self-report questionnaires and national registries.

**Methods:** Using pre-pandemic data from the Avon Longitudinal Study of Parents and Children (ALSPAC) (mean age=27.6 (standard deviation [SD]=0.5); 49% female) and UK Biobank (UKB) (mean age=56 (SD=8.1); 55% female) with data on SARS-CoV-2 infection and death-with-COVID-19 (UKB only), we investigated predictors of selection into COVID-19 analytic subsamples. We then conducted empirical analyses and simulations to explore the potential presence, direction, and magnitude of bias due to selection when estimating the association of body mass index (BMI) with SARS-CoV-2 infection and death-with-COVID-19.

**Results:** In both ALSPAC and UKB a broad range of characteristics related to selection, sometimes in opposite directions. For example, more educated participants were more likely to have data on SARS-CoV-2 infection in ALSPAC, but less likely in UKB. We found bias in many simulated scenarios. For example, in one scenario based on UKB, we observed an expected odds ratio of 2.56 compared to a simulated true odds ratio of 3, per standard deviation higher BMI.

**Conclusion:** Analyses using COVID-19 self-reported or national registry data may be biased due to selection. The magnitude and direction of this bias depends on the outcome definition, the true effect of the risk factor, and the assumed selection mechanism.

**Key messages:**

- Observational studies assessing the association of risk factors with SARS-CoV-2 infection and COVID-19 severity may be biased due to non-random selection into the analytic sample.
- Researchers should carefully consider the extent that their results may be biased due to selection, and conduct sensitivity analyses and simulations to explore the robustness of their results. We provide code for these analyses that is applicable beyond COVID-19 research.

## SECTION 1: INTRODUCTION

Understanding characteristics affecting a person’s risk of SARS-CoV-2 infection and COVID-19 severity is critical to inform policies aimed at reducing viral transmission and preventing severe illness and death. Due to the extensive data collected prior to the COVID-19 pandemic, existing population-based studies are invaluable resources to investigate risk and prognostic factors for SARS-CoV-2 infection and COVID-19, respectively.

Analyses using large-scale observational studies are often conducted on non-random subsamples of the intended target population (due to non-random study recruitment, and participants lost to follow-up or with missing data [1, 2]), and often involve variables measured with error. In non-random subsamples, when both the exposure and outcome, or a consequence of these, influence the probability of an individual being selected into the study, bias can occur (referred to as collider bias). This can induce an association between the exposure and outcome when none exists in the whole sample, or attenuate, inflate or even reverse the estimated effect of the exposure on the outcome in the selected subsample [3, 4]. Furthermore, even in representative subsamples, estimates of association might be biased in the presence of measurement error (i.e., mismeasurements or misclassifications) [5].

Selection bias may be a particular cause for concern in research analysing risk factors for SARS-CoV-2 infection or severe COVID-19 illness. These studies frequently rely on samples of individuals who either volunteered to participate in COVID-19 sub-studies, were tested for SARS-CoV-2 infection in national testing programmes or were admitted to hospital. Misclassification of SARS-CoV-2 infection status is another key potential source of bias in these studies. In particular, owing to limited testing and reporting of infection status, several studies have used “population comparison groups”, which include individuals with unknown infection status (as well as those known to not have an infection) in the comparison group [6].

In this study, we use pre-pandemic information from two UK-based observational population studies – the Avon Longitudinal Study of Parents and Children (ALSPAC) and UK Biobank (UKB), with two aims. First, we aimed to investigate factors associated with having data on SARS-CoV-2 infection and on COVID-19 severity. Second, we aimed to explore potential bias from these selection mechanisms and from the use of different comparison groups when estimating the association of factors influencing the risk of SARS-CoV-2 infection (‘risk factors’) and the severity of COVID-19 disease (‘prognostic factors’), using body mass index (BMI) as an illustrative example in empirical analyses and simulations.

## SECTION 2: EMPIRICAL COHORT ANALYSES

### Overview

Using pre-pandemic data from ALSPAC and UKB, we first investigate whether sociodemographic, behavioural, and health-related characteristics predict being selected into the subsample with data on SARS-CoV-2 infection (ALSPAC and UKB) and death-with-COVID-19 as a proxy for COVID-19 severity (UKB only). We then explored the impact of selection and misclassification bias on the estimated effect of BMI with SARS-CoV-2 infection and death-with-COVID-19 by comparing different sets of comparison groups.

### Methods

#### Prospective cohort studies and SARS-CoV-2 subsamples

##### ALSPAC

The multigenerational ALSPAC birth cohort recruited 14,541 pregnancies (∼75% response of eligible women; Generation-0 [G0] mothers), who gave birth to 14,062 children (Generation-1 [G1]) in the former county of Avon in the South West of England in 1991-1992 [7, 8]. These mothers and children have since been followed with regular assessments (including questionnaires, anthropometric and physical measurements). Since the initial G1 children were aged 7-years, a further 913 eligible children (born in the South West of England in the same years as the initial recruited children) were enrolled [9]. Hereafter we will refer to the participants where the G1 index child was alive at one year of age and who did not withdraw consent for their data to be used (14,849 G1 participants and 14,282 G0 mothers). Ethical approval was obtained from the ALSPAC Ethics and Law Committee and the local research ethics committees. Informed consent for the use of data collected via questionnaires and clinics was obtained from participants following the recommendations of the ALSPAC Ethics and Law Committee at the time (details and reference numbers of all ethics approvals can be found at http://www.bristol.ac.uk/media-library/sites/alspac/documents/governance/Research%20Ethics%20Committee%20approval%20references.pdf). The study website contains details of all the data available through a fully searchable data dictionary and variable search tool: http://www.bristol.ac.uk/alspac/researchers/our-data/. The work we present here was approved by the ALSPAC Ethics and Law Committee under project B3543.

Since April 2020, participants were sent four questionnaires to collect self-reported information relevant for studies of the COVID-19 pandemic and its consequences (including general health, seasonal symptoms, recent travel, impact of the pandemic on behaviours, mental health, well-being, healthcare/key worker status, and living arrangements during the pandemic). As part of these questionnaires’ participants were asked: “*Do you think that you have, or have had, COVID-19?*”. Supplementary Figures 1a and 1b show participant flow diagrams for the G1 and G0 mothers cohorts. These data were collected and managed using the REDCap electronic data capture system hosted at the University of Bristol [10].

#### Analytical subsamples

This study focuses on the first (Q1) or second (Q2) COVID-19 questionnaires, sent between 9^th^ April and 14^th^ May 2020, and between 26^th^ May and 5^th^ July 2020, respectively [11, 12]. We refer to ALSPAC participants who replied to either Q1 or Q2 (or both) as the SARS-CoV-2 subsample. We hypothesised that selection pressures affecting reporting a SARS-CoV-2 infection would differ during (Q1) and after (Q2) the first UK lockdown. We carried out analyses separately for questionnaires Q1 and Q2. In these questionnaires participants could indicate their COVID-19 status: a) yes, confirmed by a positive test, b) yes, doctor’s suspicion, c) yes, own suspicion, or d) do not think they had COVID-19. Of the 14,849 G1 participants recruited at baseline, 2,966 and 2,704, respectively, replied to the question referring to their COVID-19 status in Q1 and Q2. Of the 14,282 G0 mothers recruited at baseline, 2,685 and 2,632, respectively, replied to the question referring to their COVID-19 status in Q1 and Q2. These subsamples are hereafter referred to as the “assessed” subsamples.

### UK Biobank

UKB recruited 503,317 UK adults (aged 37-73) from 22 centres across England, Wales and Scotland between 2006 and 2010 (5.5% response rate) [13]. Participants attended baseline assessment centres, which collected data via touch-screen questionnaires, face-to-face interviews, physical measurements, and biological samples. Some participants have been followed up with further assessments (e.g. questionnaires, imaging studies and more recently serology tests for SARS-CoV-2) [13, 14]. Here, we used data from baseline assessment centres, linked hospital episode statistics (HES), mortality statistics, and Public Health England (PHE) polymerase chain reaction (PCR) test results for active SARS-CoV-2 infection. Ethical approval for UKB project 16729 was provided by UK Biobank, although no specific approval is required for analyses relating to COVID-19.

The SARS-CoV-2 subsample in UKB refers to all participants with a PCR test (either positive or negative) for SARS-CoV-2 infection and/or COVID-19 mentioned on their death certificate. Data on SARS-CoV-2 PCR tests conducted in England by PHE include the date the test was carried out, the specimen type (e.g., upper respiratory tract), the laboratory, the origin (e.g., hospital inpatient or community-based testing) and the result (positive or negative). At the time of analysis, PCR test data were available for the period from 16^th^ March 2020 to 1^st^ February 2021.

COVID-19 deaths were defined as participants with COVID-19 recorded on their death certificate, with ICD-10 code U07.1 (laboratory confirmed COVID-19) or ICD-10 code U07.2 (a clinical or epidemiological diagnosis of COVID-19) [15]. COVID-19 could be either the primary or contributory cause of death (i.e., they could have died *from* COVID-19 or *with* COVID-19). To encompass both definitions, we refer to death-with-COVID-19 throughout.

#### Analytical subsamples

Testing data are available separately for England, Scotland, and Wales. As each nation set their own restrictions, including lockdowns and capacity for testing, we hypothesise that selection mechanisms will differ for England, Scotland, and Wales. Therefore, UK Biobank participants who attended a baseline assessment centre in Wales or Scotland were excluded. We hypothesised that selection pressures affecting receiving a test for SARS-CoV-2 infection will strongly differ before and after mass-testing becomes available. For this reason we stratify our analyses into predictors of infection/death-with-COVID-19 pre-mass testing (excluding those who died before the 1^st^ of January 2020, and not recording infections/deaths after 18^th^ May 2020) and post-mass testing (excluding all those who died prior to 18^th^ May 2020, and including all subsequent SARS-CoV-2 tests and deaths until 1^st^ February 2021) [16]. Of the 502,117 participants in UK Biobank at the time of our analysis, 28,537 died before 1^st^ January 2020 and a further 1,811 participants died between the 1^st^ of January 2020 and the 18^th^ of May 2020. The resultant sample sizes were 421,037 and 419,226 for pre- and post- mass testing analytical samples, respectively (see participant flow diagram in Supplementary Figure 1c). Of these, 4,869 (1%) and 56,616 (14%) had a PCR test, referred to hereafter as the “assessed” subsamples.

#### Study variables

##### Pre-pandemic candidate predictors of selection

We selected *a priori* a range of sociodemographic factors, behavioural measures, physical measures, and comorbidities as putative predictors of selection into SARS-CoV-2 subsamples, as detailed in **Supplementary Table 1**. In ALSPAC, these variables were derived using data from questionnaires and follow-up visits collected between 1991 and 2017 (previously defined elsewhere [17]), while in UKB they were derived using data from baseline assessment centres (2006-2010) and linked longitudinal HES data (available from 1997 to December 2020).

#### Definition of SARS-CoV-2 infection (ALSPAC and UK Biobank) and COVID-19 death (UK Biobank)

##### ALSPAC

SARS-CoV-2 infection cases, denoted SARS-CoV-2 (+), were defined as G1 young adults and G0 mothers who, in COVID-19 questionnaires Q1 or Q2, reported getting a SARS-CoV-2 infection that was either confirmed by a positive test, indicated by a doctor or suspected by the participant [11, 12]. We included own-suspected cases in this definition because in April 2020 tests were scarce.

SARS-CoV-2 infection comparison groups were defined in two ways: 1) in the assessed sample, as those participants who reported not having had a SARS-CoV-2 infection (denoted SARS-CoV-2 (-)), and 2) in the whole sample, as those participants who reported not having had a SARS-CoV-2 infection or who did not have data on SARS-CoV-2 infection (i.e., either they did not respond to the relevant questions or did not receive or return the questionnaire) (hereafter referred to as “everyone else”).

COVID-19 severity could not be studied in ALSPAC. Although Q2 included questions about hospitalization due to COVID-19 (A4-A7), an insufficient number of participants had reported being hospitalized and we do not have access to death registry data in this cohort [11].

### UK Biobank

SARS-CoV-2 infection cases, denoted SARS-CoV-2 (+), were defined as those with either a positive PCR test result or with COVID-19 recorded on their death certificate (either ICD-10 code U07.1 or U07.2).

SARS-CoV-2 infection comparison groups were defined in two ways: 1) in the assessed sample, as those participants with a negative PCR test result and who did not have a death recorded with COVID-19 as a cause (denoted SARS-CoV-2 (-)) and 2) in the whole sample, as those with a negative PCR test result or no PCR test result who did not have a death recorded with COVID-19 as a cause (hereafter referred to as “everyone else”).

We consider death-with-COVID-19 as a severe COVID-19 endpoint. At the time of analysis, mortality data was available until 18^th^ December 2020. Comparisons groups for death-with-COVID-19 were defined in two ways: 1) in the SARS-CoV-2 (+) subsample, as participants with a positive PCR test who did not die with COVID-19 or 2) in the whole sample, as participants with either a positive PCR test but who did not die with COVID-19, with a negative PCR test, or who were not assessed (i.e. did not have a PCR test).

#### Statistical analyses

##### Assessing candidate predictors of selection into SARS-CoV-2 subsamples

We used univariable logistic regression models to estimate the association between each candidate predictor of selection (**Supplementary table 1**) and the outcomes describing selection into subsamples of SARS-CoV-2 infection and death-with-COVID-19 analyses (Table 1) as described below. We used four comparisons to better understand processes influencing being selected into the SARS-CoV-2 subsamples in ALSPAC and UKB (Table 1): i) having data on SARS-CoV-2 infection compared to having no data (assessed vs non-assessed), ii) assessed as having had SARS-CoV-2 infection compared to having no data (SARS-CoV-2 (+) vs non-assessed), iii) assessed as *not* having had a SARS-CoV-2 infection compared to having no data (SARS-CoV-2 (-) vs non-assessed), and iv) assessed as having had a SARS-CoV-2 infection compared to assessed as *not* having had a SARS-CoV-2 infection (SARS-CoV-2 (+) vs SARS-CoV-2 (-)). Similar definitions were initially proposed by Chadeau-Hyam *et al*. [18] to help identify individual characteristics influencing someone’s risk of being assessed (indicative of non-random sample selection) from someone’s risk of being classified as a case once assessed (indicative of a characteristic influencing SARS-CoV-2 infection risk). For instance, associations which are of similar direction and magnitude between comparison “ii” (SARS-CoV-2 (+) vs non-assessed) and “iii” (SARS-CoV-2 (-) vs non-assessed) might suggest that the association of a candidate predictor of selection with reporting or testing positive for SARS-CoV-2 is due to confounding or selection among assessed individuals rather than a causal effect on susceptibility to SARS-CoV-2 infection.

**Table 1:**
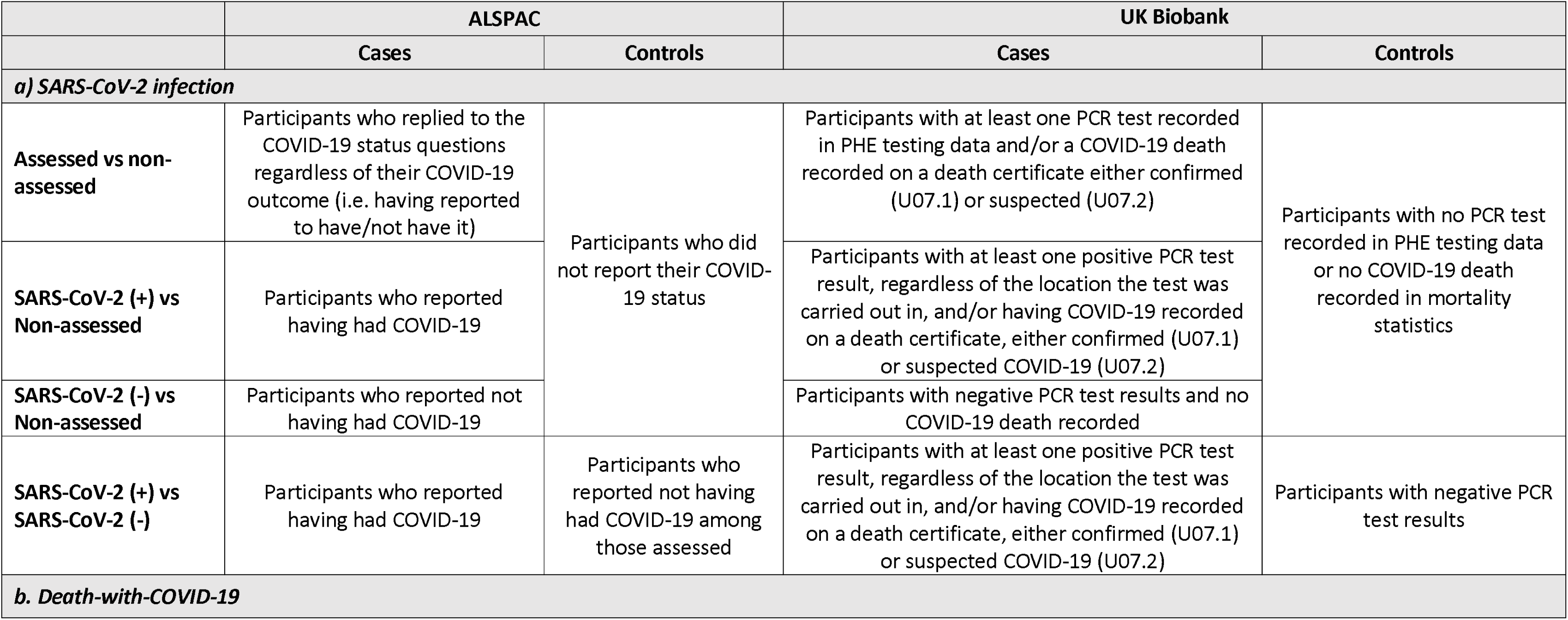
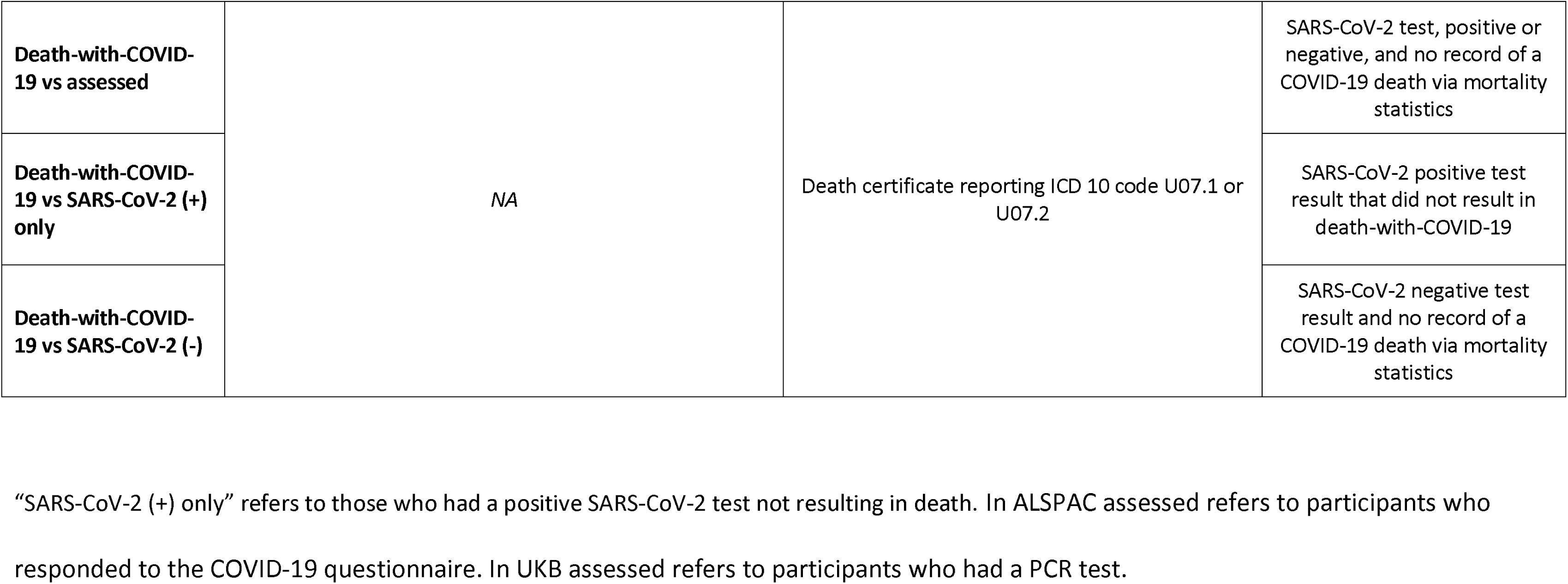
Definition of outcomes related to being selected into the subsamples with SARS-CoV-2 and COVID-19 data used to assess predictors of selection (aim 1)

We derived three comparisons to better understand processes influencing selection bias in analyses of COVID-19 death in UKB (Error! Reference source not found.): i) death-with-COVID-19 compared to assessed as positive for SARS-CoV-2 infection (death vs SARS-CoV-2 (+)), ii) death-with-COVID-19 compared to the tested sample (death vs assessed) and iii) death-with-COVID-19 compared to testing negative for SARS-CoV-2 infection (death vs SARS-CoV-2 (-)). Again, such comparisons can help tease apart factors associated with death-with-COVID-19 due to selection bias (either due to selection into the tested sample, as in the infection scenario above, or due to conditioning on infection [19]) or due to influencing SARS-CoV-2 risk or risk of death-with-COVID-19. For example, associations that are of similar direction and magnitude between comparison “i” (death vs SARS-CoV-2 (+)) and “ii” (death vs assessed) might suggest that the association of a candidate predictor of selection with death is not biased due to conditioning on infection (which could influence estimates from comparison “i”) or by a causal effect on susceptibility to SARS-CoV-2 infection (which could influence estimates from comparison “ii”).

In both ALSPAC and UKB we used test-wise deletion, where the candidate predictor of selection under study was required to have complete data.

##### Exploring bias due to selection in the association of BMI with SARS-CoV-2 infection and death-with-COVID-19

We used logistic regression models to estimate the association of BMI with SARS-CoV-2 infection (in ALSPAC and UKB) and COVID-19 death (in UKB only) following the case and comparison groups definitions described in detail in the ‘Definition of SARS-CoV-2 infection and COVID-19 death’ subsection. We carried out complete case analyses, where included individuals had no missing data for all exposures, outcomes, and covariates. We adjusted the models for potential confounders as defined in Figure 1: age, sex, smoking status, educational attainment, and deprivation indices (Index of Multiple Deprivation in ALSPAC and Townsend deprivation index in UKB).

**Figure 1:**
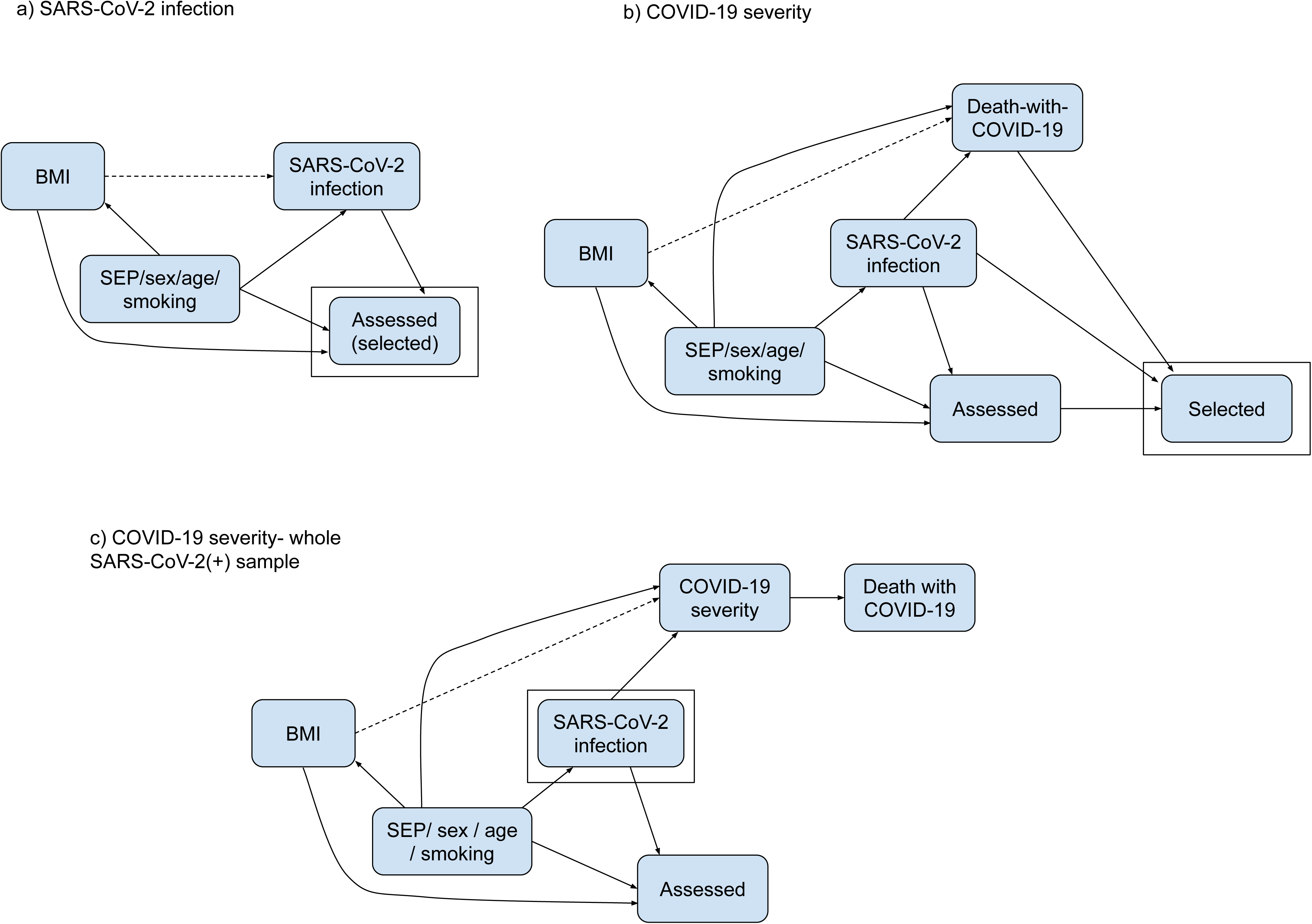
Directed acyclic graphs depicting assumed causal model for empirical and simulation scenarios. a) SARS-CoV-2 infection; b) Death-with-COVID-19. Dashed lines indicate the causal effect we are estimating. a) Simulations based on ALSPAC and UKB data. Participants were assessed (and hence selected), if they self-reported having had a SARS-CoV-2 infection in ALSPAC, or had a SARS-CoV-2 PCR test result in UKB. b) Simulations based on UKB data only. Participants were selected if they were assessed (as in (a)) and tested positive, or if they died with COVID-19. An arrow from node A to node B in a DAG indicates that A is a direct cause of B (i.e., A affects B not only through another node in the DAG). DAGs do not describe how this effect occurs, i.e., the specific model describing this relationship, including whether nodes interact in their effects. For example, in DAG (b), infection is a direct cause of death-with-COVID-19 as a person can only die with COVID-19 if they are infected. Thus, infection interacts with all other direct effects of death-with-COVID-19. For example, smoking directly affects the risk of dying with COVID-19 only among those with a SARS-CoV-2 infection (i.e., the effect of smoking on death-with-COVID-19 depends on SARS-CoV-2 infection status).

### Results

#### Descriptive analyses

##### ALSPAC

Among the 2,966 participants assessed for COVID-19 in Q1, 2,122 (72%) were females and their mean age was 27.6 (standard deviation [SD] = 0.54) years (**Supplementary table 2A**). With the exceptions of age and sex, all candidate predictors of selection had missing data, ranging from 11% for the urban/rural index (calculated in 2014) to 74% for education level (reported when they were approximately 27 years old) (**Supplementary table 3**). **Supplementary tables 2B and 3** also include the characteristics of G0 mothers.

##### UK Biobank

Of the 421,037 UK Biobank participants included in the pre-mass testing analyses, 55% were female with a mean age of 56.3 years (SD = 8.1). Pre-mass testing, 1% of participants had a recorded SARS-CoV-2 test or death, where 30% of the tests were positive. Post mass testing, this increased to 14% for tests or deaths, where 23% of the tests were positive. Prevalence of comorbidities ranged from 13% for previous autoimmune diagnoses, up to 44% for a cardiovascular diagnosis (**Supplementary table 2C**). Few candidate predictors of selection had missing data (**Supplementary Table 3**).

#### Assessing candidate predictors of selection into SARS-CoV-2 subsamples

##### ALSPAC

###### Selection into the subsample for SARS-CoV-2 infection analyses

Most of the candidate predictors of selection were associated with having self-reported SARS-CoV-2 infection data (i.e., being assessed) from COVID-19 questionnaire Q1 among G1 participants. Being female, older, having higher education attainment, living in non-urban areas, suffering from adverse mental health outcomes and having higher BMI and diastolic blood pressure were associated with a higher odds of being assessed (odds ratio [OR] ranging from 1.08 for age [95% CI: 1.04; 1.13] to 3.29 for sex [95% CI: 3.01; 3.59]). Being non-white, a current smoker, having a history of alcohol abuse, living in more deprived areas and suffering from some autoimmune comorbidities were associated with a lower odds of being assessed (OR ranging from 0.60 for ethnicity [95% CI: 0.47; 0.75] to 0.76 for Index of Multiple Deprivation (IMD) [95% CI: 0.74; 0.78]) (Figure 2).

**Figure 2:**
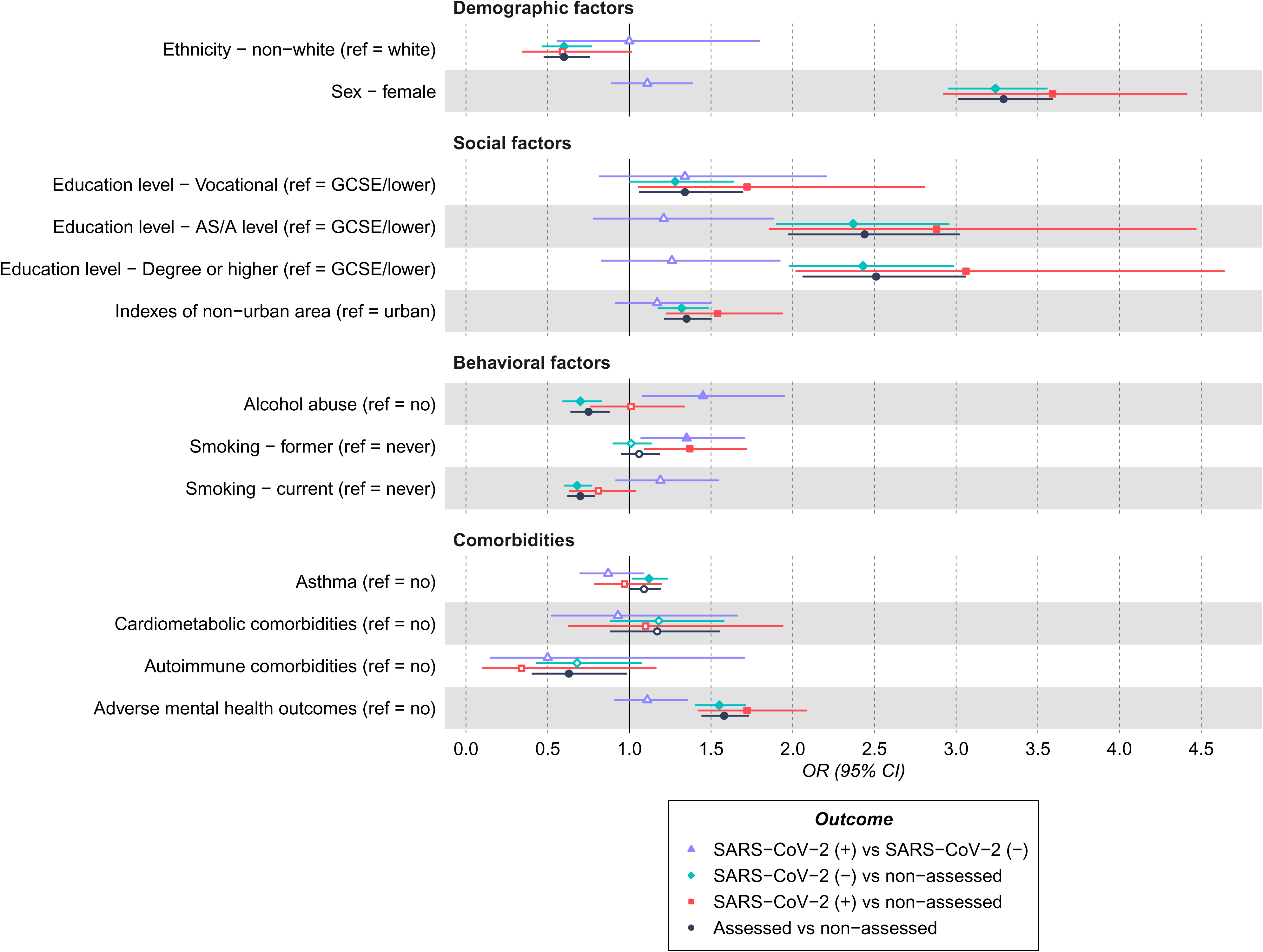
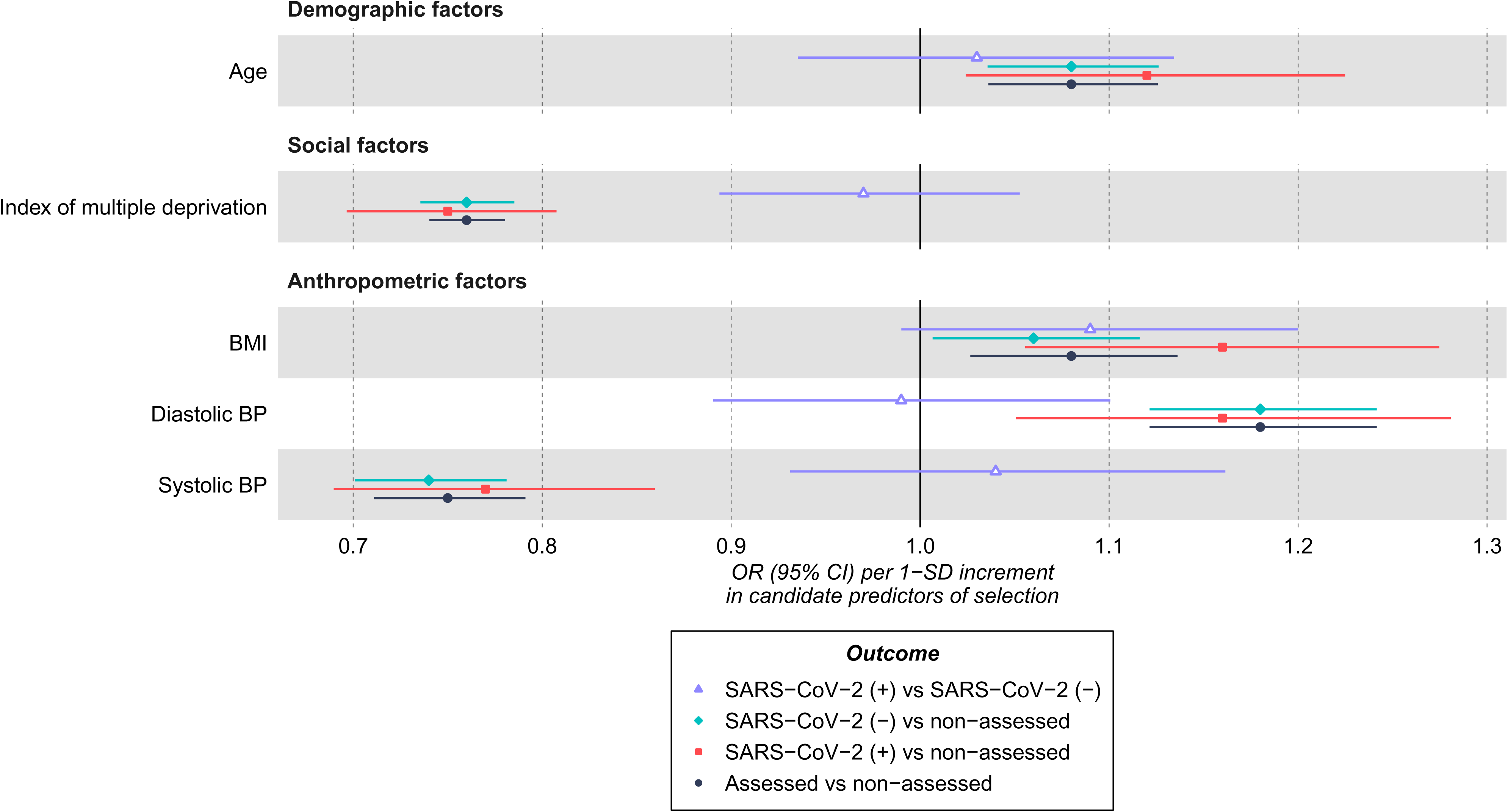
Forest plots of the association between the candidate predictors of selection and outcomes related to SARS-CoV-2 infection in the ALSPAC G1 cohort – questionnaire 1. a) Categorical variables; b) Continuous variables. ORs and their 95% confidence intervals are shown for categorical variables (a) and continuous variables (b). Estimates for continuous candidate predictors are per 1 standard deviation for each predictor except for IMD which is given per 1 higher quantile.

Similar results were obtained from analyses comparing SARS-CoV-2 (+) or SARS-CoV-2 (-), respectively, to non-assessed participants. However, when SARS-CoV-2 (+) participants were compared with SARS-CoV-2 (-) participants (i.e. restricting the analyses to the participants assessed for SARS-CoV-2 infection), only smoking and alcohol abuse were associated with higher odds of SARS-CoV-2 infection (OR = 1.38 [95% CI: 1.10; 1.75] for smoking; OR = 1.45 [95% CI: 1.07; 1.94] for alcohol abuse) (Figure 2).

The pattern of associations between candidate predictors and outcomes related to selection into SARS-CoV-2 infection analyses were consistent between the two questionnaires (**Supplementary figure 2**), as well as between the G1 and G0 mothers cohorts (**Supplementary figure 3**), with a few exceptions. Exceptions included BMI and comorbidities, where in contrast to G1, higher BMI and comorbidities were associated with lower and higher odds of being assessed for SARS-CoV-2 infection among the G0 mothers.

##### UK Biobank

###### Selection into the subsample for SARS-CoV-2 infection analyses

Except for diastolic blood pressure, all variables were associated with being assessed (tested) for SARS-CoV-2 infection pre-mass testing. Variables associated with a higher odds of being assessed include being older, reporting non-white ethnicity, being a former or current smoker, having higher BMI and pre-existing comorbidities (e.g. OR = 3.15 for a previous cardiovascular diagnosis [95% CI: 2.96; 3.35]). Being female, living in a rural area and having higher educational attainment were associated with a lower odds of being assessed (e.g. OR = 0.92 for vocational qualifications (compared with GCSE or less) [95% CI: 0.85; 0.98]) (Figure 3).

**Figure 3:**
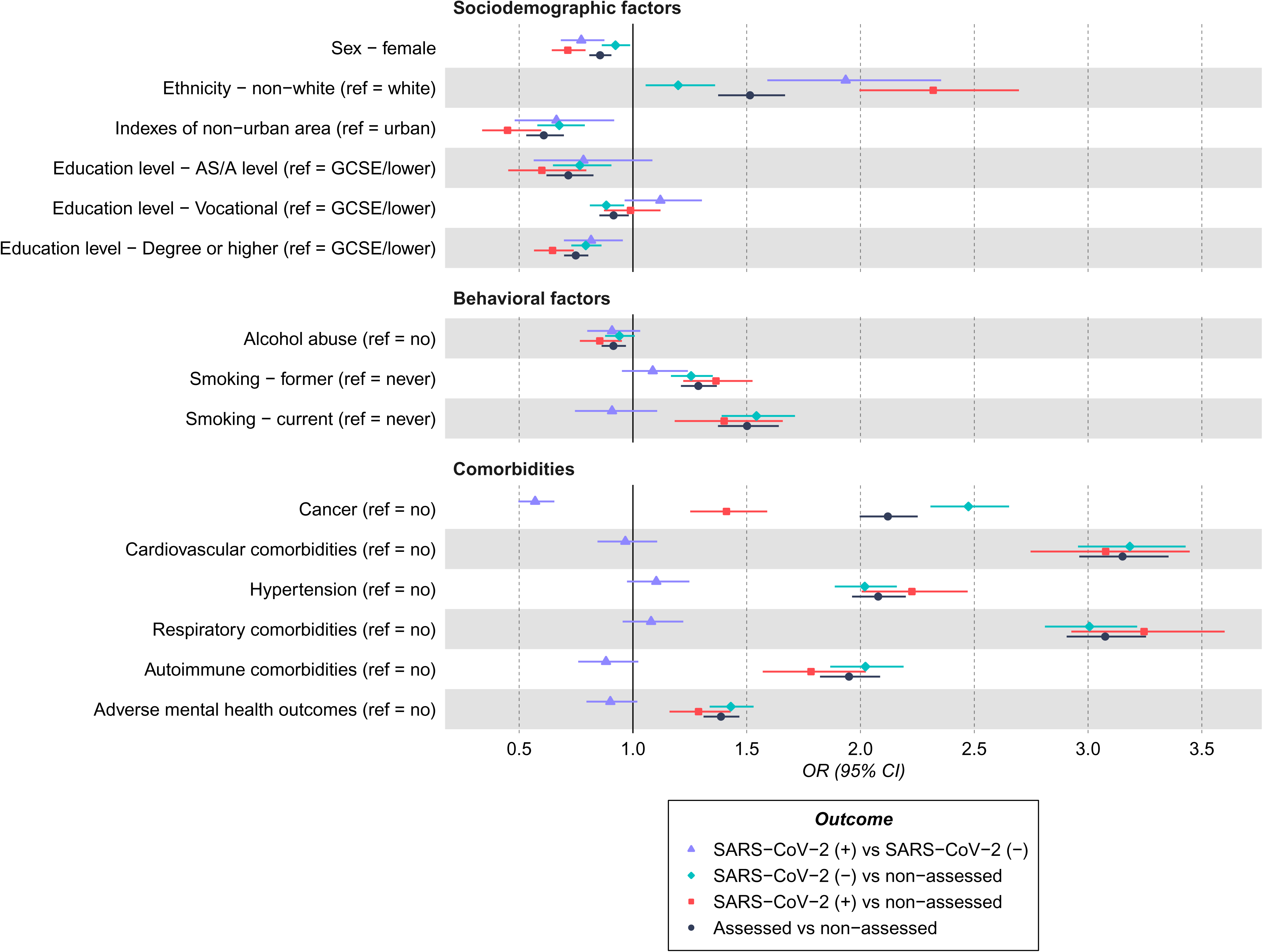
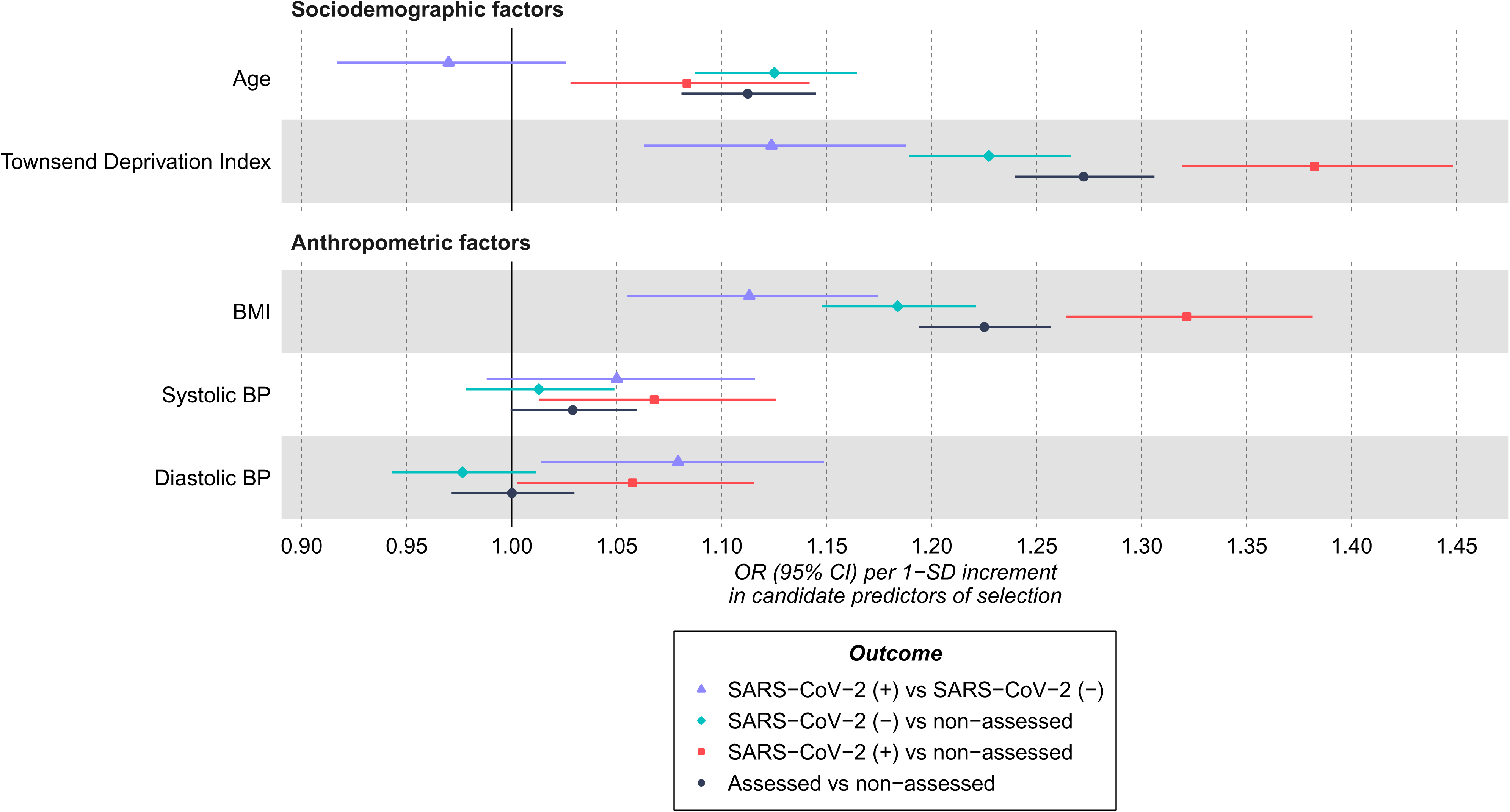
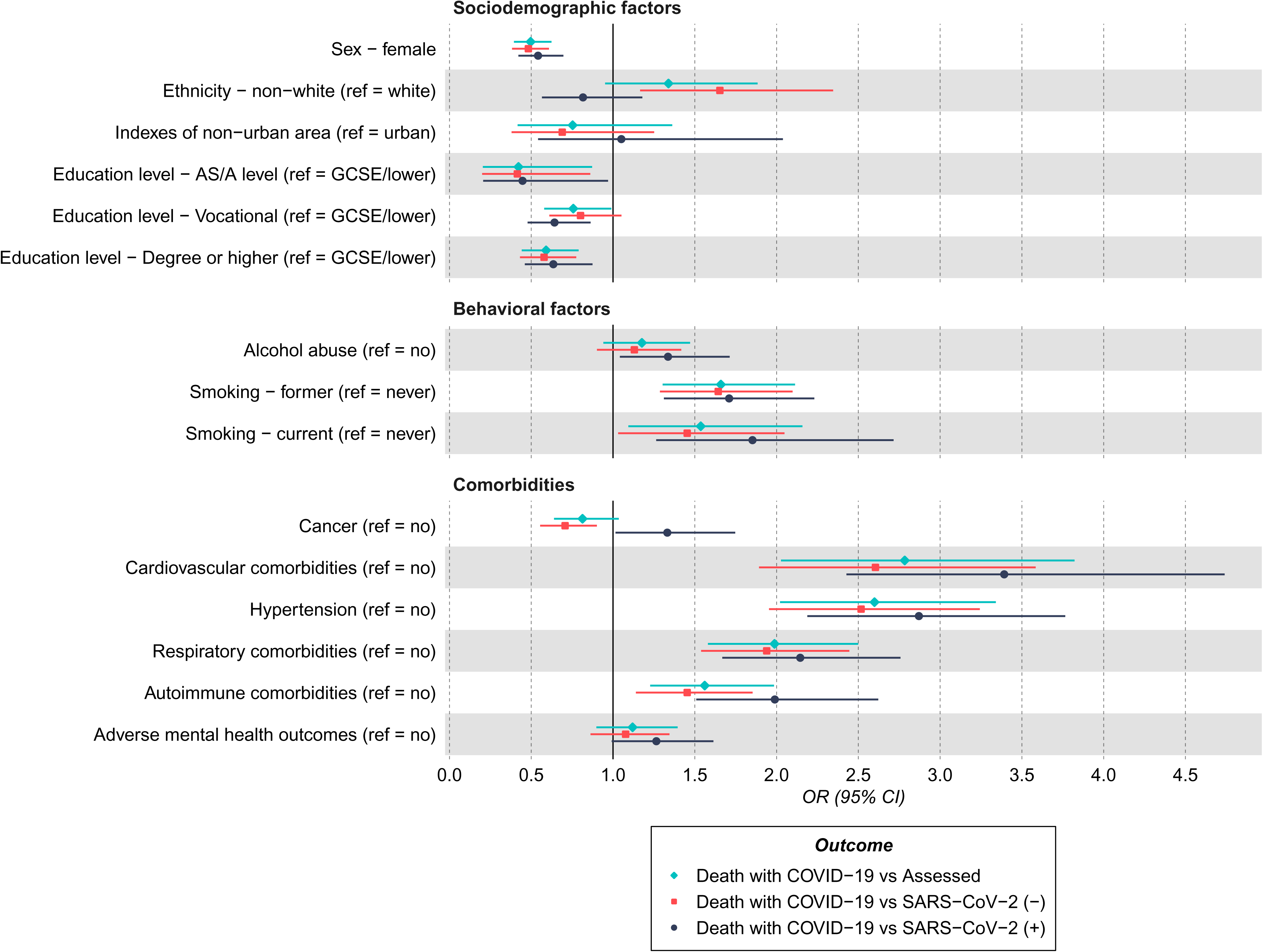
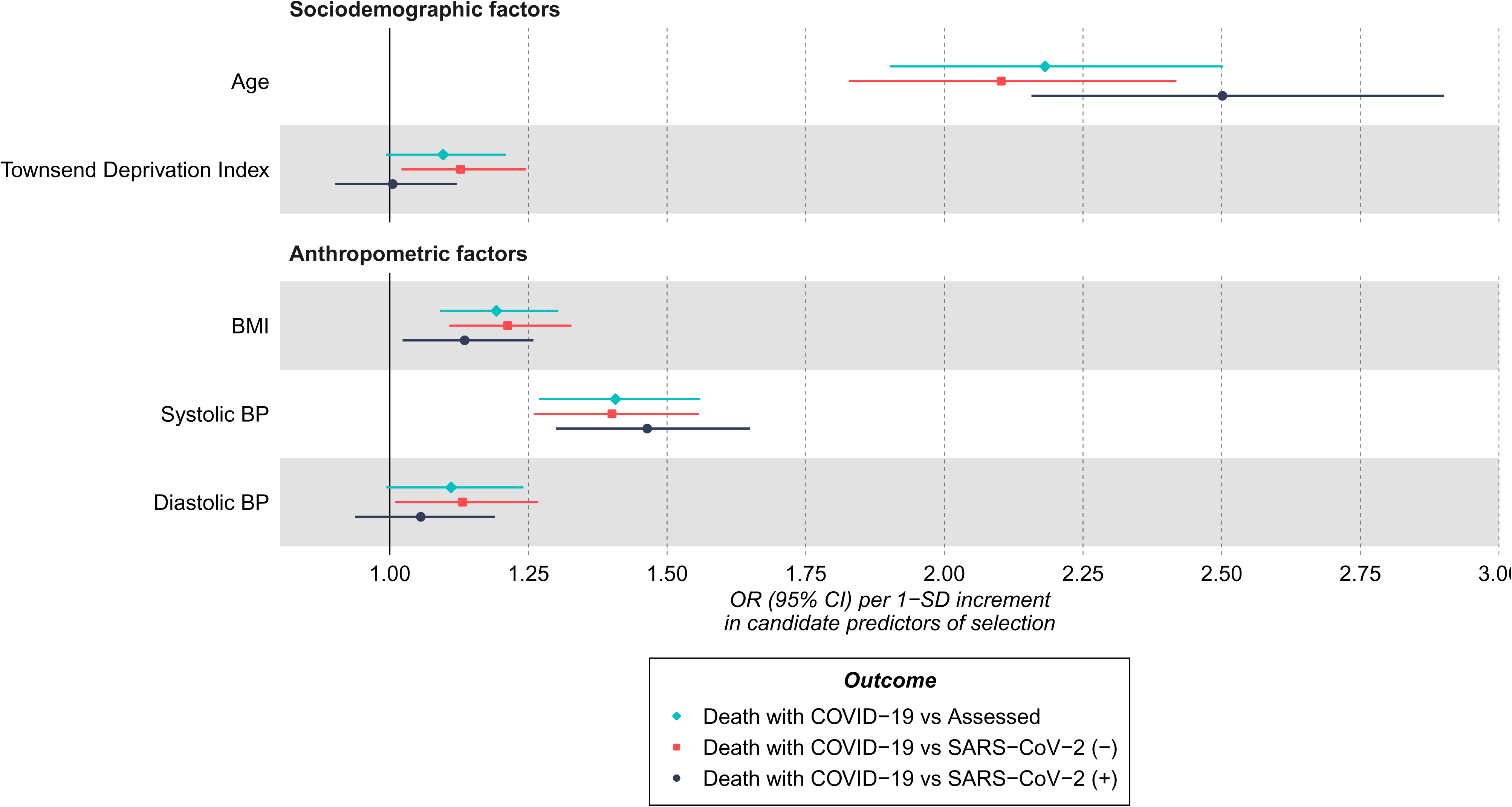
Forest plots of the association between the candidate predictors of selection and outcomes related to SARS-CoV-2 infection and death-with-COVID-19 in UKB – pre-mass testing 1. a) SARS-CoV-2 infection - Categorical variables; b) SARS-CoV-2 infection - Continuous variables; c) Death with COVID-19 - Categorical variables; d) Death with COVID-19 - Continuous variables. ORs and their 95% confidence intervals are shown separately for SARS-CoV-2 infection (a, b) and death-with-COVID-19 (c, d). Estimates for continuous candidate predictors are per 1 standard deviation for each predictor.

These results were directionally consistent with results comparing non-assessed participants with both those testing i) SARS-CoV-2 (+) and ii) SARS-CoV-2 (-). When restricting to the assessed subsample, all sociodemographic, behavioral and anthropometric variables, with the exception of current smoking and age, were associated with testing SARS-CoV-2 (+) compared with testing SARS-CoV-2 (-) pre-mass testing. No pre-existing comorbidities, except for cancer, were associated with testing SARS-CoV-2 (+) versus SARS-CoV-2 (-) (Figure 3).

The directions of these effects were similar between the pre- and post-mass testing analyses. However, for some variables, such as socioeconomic indicators and comorbidities, the magnitude of the associations for being assessed (versus not being assessed) were lower post-mass testing. For example, the association for being assessed vs not being assessed per unit increase in Townsend deprivation index decreased from OR = 1.27 (95% CI: 1.24 to 1.32) before mass testing to 1.08 (95% CI: 1.07 to 1.09) following the introduction of mass testing. In addition, when restricting the sample to those assessed for SARS-CoV-2, pre-existing comorbidities were associated with a lower odds of testing SARS-CoV-2 (+) in post-mass testing in contrast to pre-mass testing (**Supplementary Figure 4**).

###### Selection into the subsample for death-with-COVID-19 analyses (among assessed participants)

Compared with SARS-CoV-2 (+) not resulting in death-with-COVID-19, being male, having lower educational attainment, being a former or current smoker, having pre-existing comorbidities, being older and having a higher BMI were all associated with increased odds of dying with COVID-19. These results were similar when comparing those who died with COVID-19 with i) tested participants (both SARS-CoV-2 (+) and (-)) and ii) SARS-CoV-2 (-) participants confirmed via PCR tests (Figure 3).

#### Exploring bias due to selection in the association of BMI with SARS-CoV-2 infection and death-with-COVID-19

##### ALSPAC

###### BMI and risk of SARS-CoV-2 infection

Per standard deviation increase in BMI, the OR for SARS-CoV-2 infection among G1 participants was 1.08 [95% CI: 0.96; 1.21] in the assessed subsample and 1.10 [95% CI: 0.98-1.23] when considering the whole sample (Figure 4). Corresponding results for Q2 for G1 participants and G0 mothers are available in **Supplementary figures 5** and **6**.

**Figure 4:**
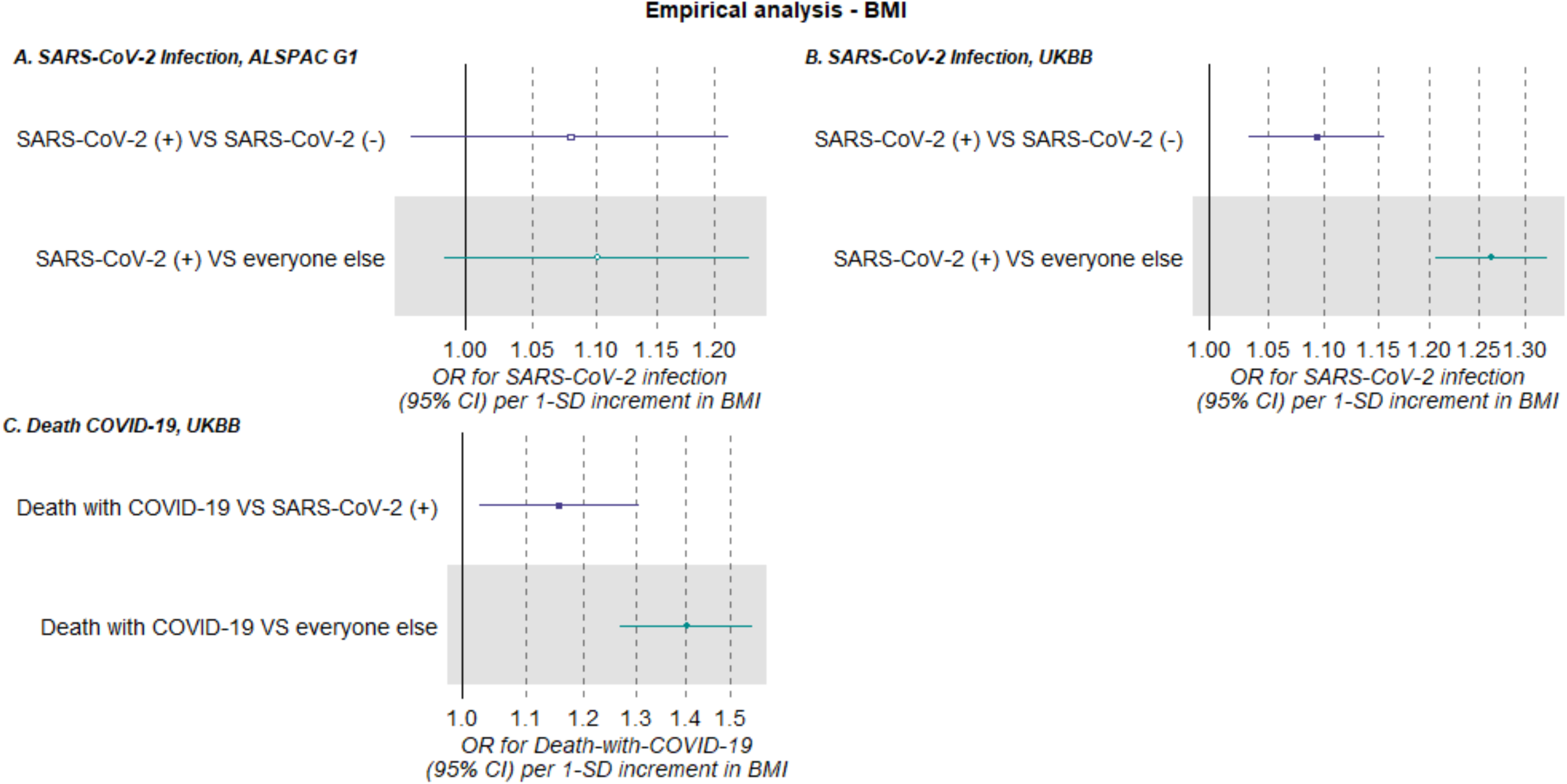
Forest plots of the association between BMI and COVID-19-related outcomes. a) SARS-CoV-2 infection in the ALSPAC G1 cohort reported in questionnaire Q1; SARS-CoV-2 (+) vs SARS-CoV-2 (-) N=1,915, SARS-CoV-2 (+) vs everyone else N=2,983. b) SARS-CoV-2 infection in UKB recorded pre-mass testing; SARS-CoV-2 (+) vs SARS-CoV-2 (-) N=4,662, SARS-CoV-2 (+) vs everyone else N=409,487. c) death-with-COVID-19 in UKB recorded pre-mass testing; Death-with-COVID-19 vs SARS-CoV-2(+) not resulting in death-with-COVID-19 N=1,375, Death-with-COVID-19 vs everyone else N=409,487. Models were adjusted for age, sex, smoking, education and proxies of socioeconomic position. ‘Everyone else’ control group includes those tested and SARS-CoV-2 (-) and those not tested.

##### UK Biobank

###### BMI and risk of SARS-CoV-2 infection

Higher BMI was associated with higher odds of SARS-CoV-2 (+) in both comparison groups; effect estimates were of smaller magnitude in the analysis restricting to the assessed subsample (OR = 1.09 [95% CI: 1.03, 1.16]) compared to when considering the whole sample (OR = 1.26 [95% CI: 1.21, 1.32) (Figure 4). Similar effect estimates were identified when analyses were stratified by sex (**Supplementary figure 7**).

The association between BMI and SARS-CoV-2 (+) in the assessed sample was similar in the post-mass testing period (OR = 1.07 [95% CI: 1.05, 1.10]), however the association in the whole sample analysis was attenuated towards the null (OR = 1.17 [95% CI: 1.15, 1.19]) (**Supplementary figure 7**).

###### BMI and risk of death-with-COVID-19

Higher BMI was also associated with higher odds of death-with-COVID-19, after adjusting for age, sex, smoking, and SEP compared with both i) SARS-CoV-2 (+) not resulting in death-with-COVID-19 (OR = 1.16 [95% CI: 1.03, 1.31]) and ii) everyone else (i.e., SARS-CoV-2 (+), SARS-CoV-2 (-), and non-assessed) (OR = 1.40 [95% CI: 1.27, 1.55]) (Figure 4). The association between BMI and death-with-COVID-19 in both the assessed subsample and whole sample analyses increased following the introduction of mass testing (**Supplementary figure 8**).

## SECTION 3: SIMULATIONS

### Overview

We explored possible bias in studies of SARS-CoV-2 infection and COVID-19 death through simulation studies of the estimated effect of BMI on SARS-CoV-2 infection and death-with-COVID-19 as an exemplar. These simulation studies were informed by data from ALSPAC and UKB, as well as external information from population-based surveys, to derive plausible parameters for the simulated data. We report the aims, data-generating mechanisms, estimands, methods, and performance measures of our simulations – the ADEMP approach [20].

### Methods

#### Simulation A: Estimating the association of BMI with assessed as positive for SARS-CoV-2 infection

##### Aim

We aimed to assess the bias that may occur when estimating the association of BMI on SARS-CoV-2 infection (conditional on confounding factors) when only a subsample of participants has a SARS-CoV-2 infection assessment (i.e. self-reported having had COVID-19 in ALSPAC or had a SARS-CoV-2 PCR test in UKB), using data from ALSPAC (COVID-19 questionnaire 1, G1 cohort) and UKB (pre-mass testing) to inform plausible parameters.

##### Data generating mechanism

The data generating mechanism is based on the directed acyclic graph (DAG) shown in Figure 1a. We used simulated dataset sample sizes of 14,849, and 421,122 for the ALSPAC and UKB simulations to reflect the empirical data. The parameters of the models used to generate the simulated data were based on a combination of estimated values from ALSPAC and UKB, and statistics extracted from the published literature. We repeated the simulations assuming a) no effect of BMI on SARS-CoV-2 infection and b) a strong effect (OR=3) of BMI on SARS-CoV-2 infection. Selection in simulation A is defined as participants that were assessed for SARS-CoV-2 infection. We induce selection bias by including an additive interaction effect on the log probability scale of BMI and SARS-CoV-2 infection on selection (described further in Supplementary sections S1 and S2). We simulated three scenarios to induce different magnitudes of selection bias, with the following effect of BMI and SARS-CoV-2 infection on selection: 1) no interaction (main effects only on the log probability scale), 2) ‘plausible’ interaction effect (log RR=0.0527 in ALSPAC and -0.162 in UKB-based scenarios), and 2) ‘extreme’ interaction effect (log RR=0.135 in ALSPAC and -0.245 in UKB-based scenarios). For each of these scenarios the main effect and intercept were adjusted such that the total effect of BMI and SARS-CoV-2 infection on selection remained constant (see Supplementary Section S4 for further details). Further details of our approach and the specific models used to generate these data are provided in **Supplementary sections S1-S4** and **Supplementary tables 4** and **5**.

##### Target estimand

The estimand of interest was the OR of SARS-CoV-2 infection per SD increase in BMI, conditional on confounding factors (but not conditional on selection).

##### Methods

We evaluate two approaches to estimate the association of BMI with SARS-CoV-2 infection using logistic regression:

a. Using only the assessed subsample, with the outcome defined as SARS-CoV-2 (+) versus SARS-CoV-2 (-).
b. Using the whole sample, with the outcome defined as those assessed and SARS-CoV-2 (+) versus everyone else (i.e. assessed as SARS-CoV-2 (-) and the non-assessed).

We use Wald-type confidence intervals on the log-odds scale with the SE taken from the inverse estimated information matrix.

##### Performance measure

We estimated the bias (and Monte Carlo standard error (MCSE)) of the estimated effect of BMI on SARS-CoV-2 infection, compared to the true value, for each method described above. We estimated the coverage, i.e. the proportion of simulations where the confidence intervals included the true value. These performance measures were generated across 1000 simulation repetitions. Further details of the parameter values used are provided in Supplementary section S5.

#### Simulation B: Estimating the association of BMI with death-with-COVID-19

##### Aim

We aimed to assess bias due to selection in estimates of the association of BMI with death-with-COVID-19, as a proxy for severe COVID-19 (recalling that simulation A considered infection).

##### Data generating mechanism

The data generating mechanism was based on the DAG shown in Figure 1b. The confounders, BMI, SARS-CoV-2 infection and being assessed (selection in Simulation A) were generated as described in our SARS-CoV-2 infection simulation above. Selection in simulation B is defined as participants that were either assessed and were SARS-CoV-2 (+), or those who died with COVID-19. The parameters of the models used to generate the simulated data were based on a combination of estimated values from ALSPAC and UKB, and statistics extracted from the published literature.

In a similar way to the SARS-CoV-2 infection simulations, we repeated the simulation assuming: 1) no effect of BMI on death-with-COVID-19, and 2) OR=3 effect of BMI on death-with-COVID-19. Further details of our approach and the specific models used to generate these data are provided in **Supplementary sections S1-S4** and **Supplementary tables 4** and **5**.

##### Target estimand

OR of death-with-COVID-19 per SD increase in BMI, conditional on confounding factors and having a SARS-CoV-2 infection (but not conditional on being assessed).

##### Methods

We evaluated two approaches to estimate the association of BMI with death-with-COVID-19 using logistic regression:

a. Using only the selected subsample of those assessed and SARS-CoV-2 (+), with the outcome defined as those who died with COVID-19 versus those who were SARS-CoV-2 (+) but did not die with COVID-19.
b. Using the whole sample (i.e. those both with and without assessment for SARS-CoV-2 infection) with the outcome defined as those who died with COVID-19 versus everyone else.

##### Performance measure

We assessed bias and coverage (and MCSE), compared to the true value, for each of the above methods, as in simulation A.

### Results

All simulations using the confounder adjusted model in the full sample (i.e. no selection) were unbiased, with coverage between 0.933 (MCSE=0.0079) and 0.958 (MCSE=0.063).

#### Simulations estimating the association of BMI with SARS-CoV-2 infection and death-with-COVID-19 outcomes

##### ALSPAC

The results of our simulations of SARS-CoV-2 infection based on ALSPAC are shown in Table 2 (histograms of estimates shown in **Supplementary figure 9**).

**Table 2:**
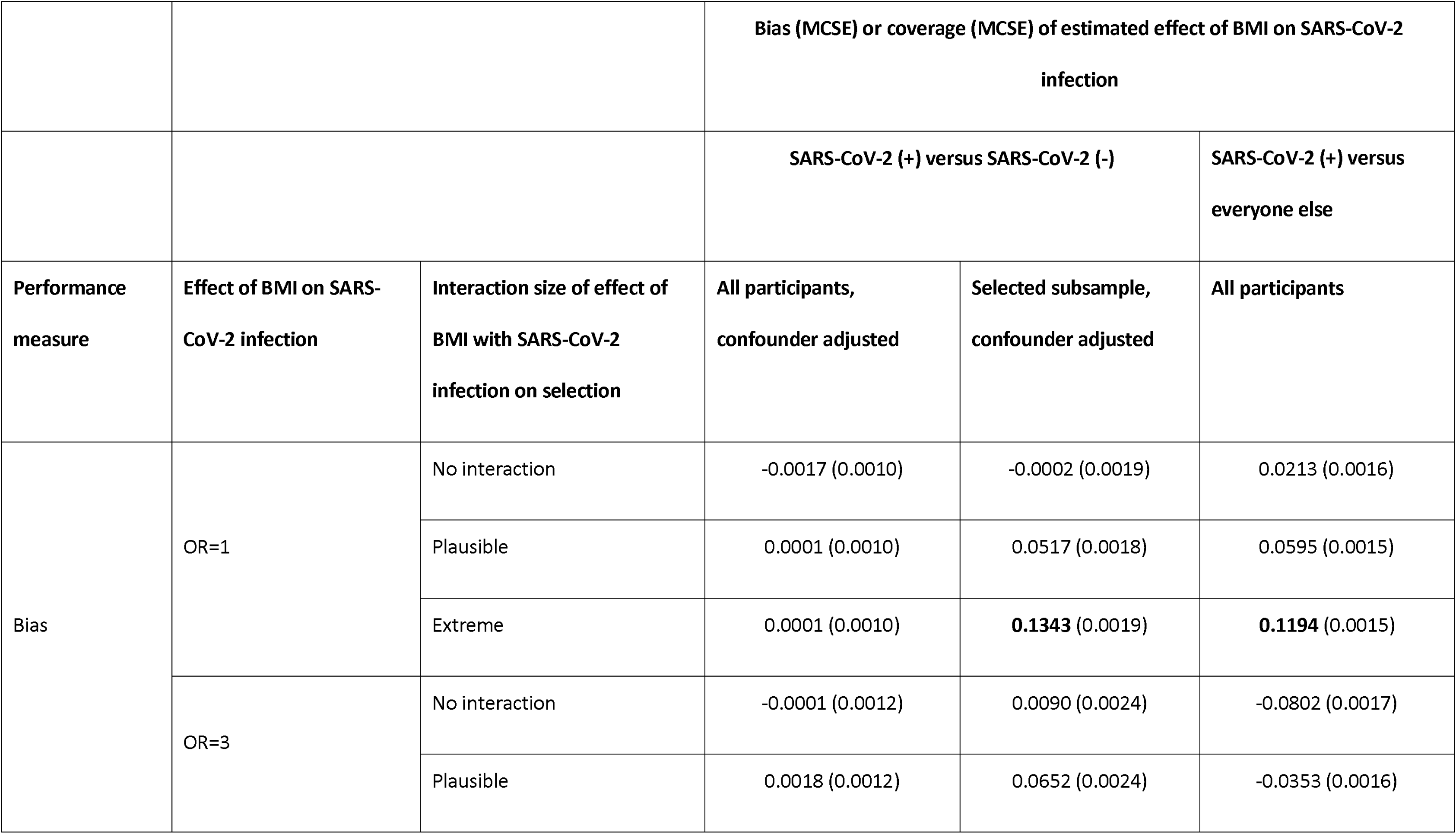
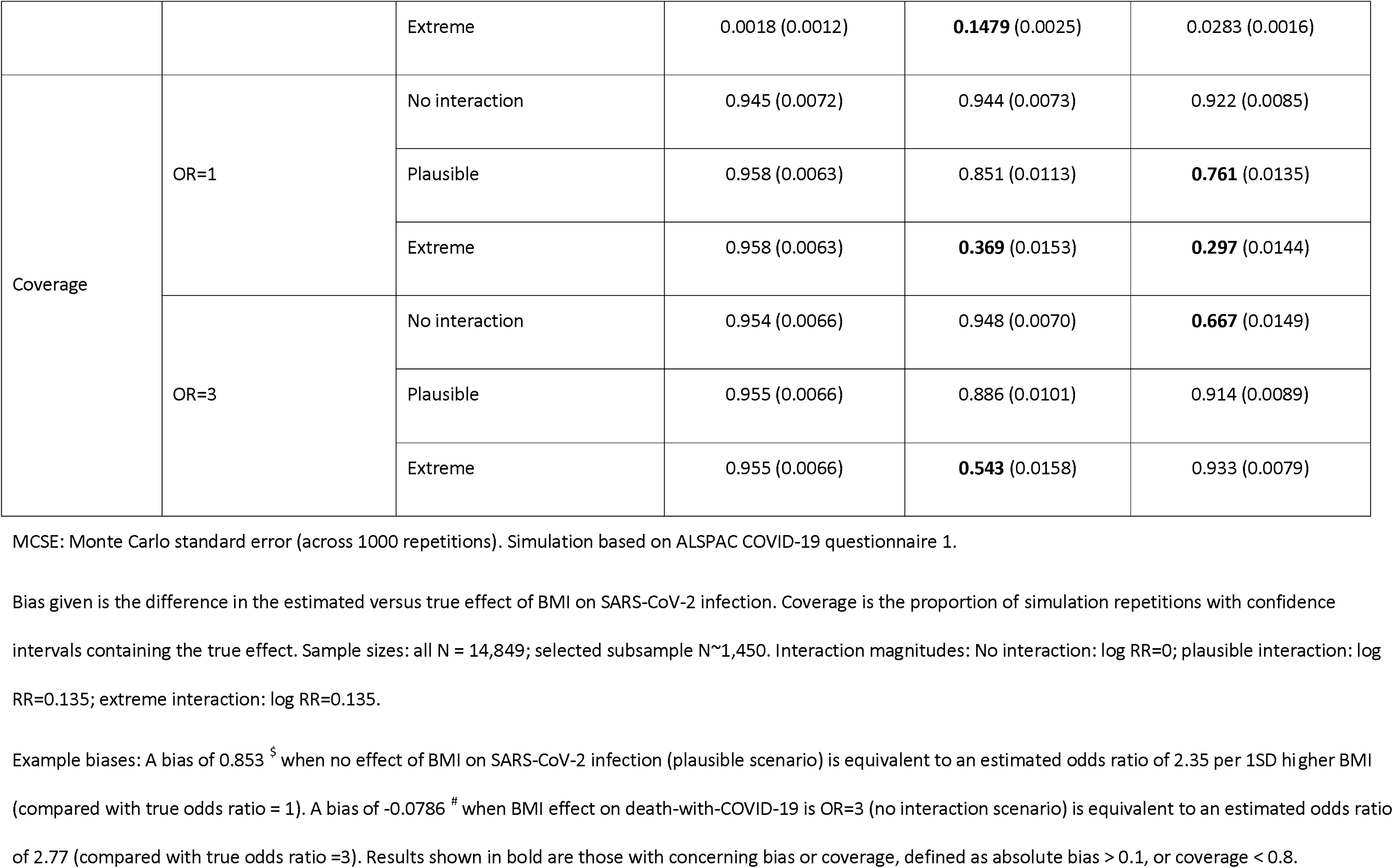
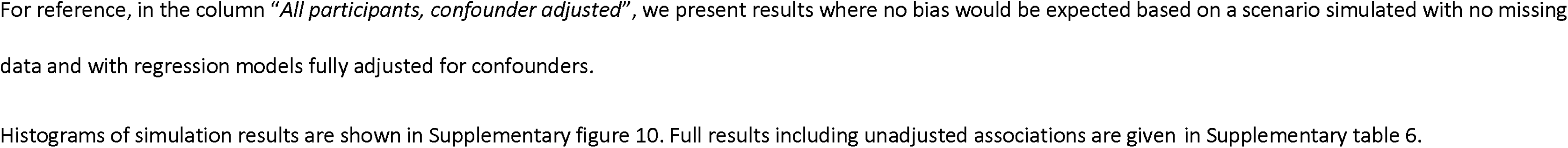
Results of simulations of SARS-CoV-2 infection based on ALSPAC G1 cohort.

When assuming independent effects on selection of BMI and SARS-CoV-2 infection (i.e. no additive interaction on the log probability scale), estimates using the SARS-CoV-2 (+) versus SARS-CoV-2 (-) outcome definition (selected subsample only) were unbiased as expected when assuming no effect of BMI on SARS-CoV-2 infection (see Supplementary figure 10a for an illustration of why this is the case for analyses using logistic regression). Unexpectedly, when assuming an effect of BMI on SARS-CoV-2 infection we found positive bias. However, this bias disappeared when increasing the sample size, suggesting it is due to near separation – that is, when a combination of covariates almost perfectly predicts the outcome [21, 22] – rather than bias due to use of a non-random subsample (see **Supplementary table 8**). In the scenarios with an interaction effect of BMI and SARS-CoV-2 infection on being assessed, bias using the SARS-CoV-2 (+) versus SARS-CoV-2 (-) outcome was positive and strengthened as the magnitude of the interaction increased.

Bias for the SARS-CoV-2 (+) versus everyone else outcome was positive when there was no effect of BMI on SARS-CoV-2 infection, but negative when there was an effect, when there was no interaction in the effects of BMI and SARS-CoV-2 infection on being assessed i.e. in the presence of misclassification bias only (expected ORs of exp(0.0213)=1.02 compared to a true odds ratio of 1, and exp(ln(3)-0.0802)=2.77 compared to a true OR of 3). As the magnitude of the (positive) interaction effect increased (i.e. increasing amounts of selection bias in addition to misclassification bias), bias became less negative/more positive i.e. decreasing in magnitude of negative bias as the interaction magnitude increased then increasing magnitude of positive bias as the interaction magnitude increased further expected ORs of 1.06 and 2.90 for the plausible interaction effect sizes, when BMI does not and does affect SARS-CoV-2 infection, respectively). Coverage varied greatly depending on the scenario, ranging between 0.369 and 0.948 for the SARS-CoV-2 (+) versus SARS-CoV-2 (-) outcome, and between 0.297 and 0.933 for the SARS-CoV-2 (+) versus everyone outcome. In general, coverage reduced (away from 0.95) as the size of the interaction increased, apart from the extreme example using the SARS-CoV-2 (+) versus everyone else outcomes assuming an effect of BMI on SARS-CoV-2 infection for which coverage increased (towards 0.95).

#### UK Biobank

The results of our simulations of SARS-CoV-2 infection based on UKB data are shown in Table 3a (histograms of estimates shown in Supplementary figure 11a). Results for SARS-CoV-2 (+) versus SARS-CoV-2 (-) for the ‘no interaction’ scenario were similar to that in ALSPAC (see **Supplementary table 9** for results with larger sample). In the scenarios with an interaction effect of BMI and SARS-CoV-2 infection on being assessed we found negative bias (the opposite direction to ALSPAC because the interaction was in the opposite direction) that strengthened as the interaction magnitude increased. Bias for the SARS-CoV-2 (+) versus everyone else outcome was positive when there was no interaction in the effect of BMI and SARS-CoV-2 infection on being tested i.e. with misclassification bias only (e.g. bias = 0.1637 ([MCSE=0.0012] when assuming no effect of BMI on SARS-CoV-2 infection). As the magnitude of the (negative) interaction effect increased (i.e. increasing amounts of selection bias in addition to misclassification bias), bias became less positive/more negative (e.g. bias of 0.0259 (MCSE=0.0012) and -0.0386 (MCSE (0.0012) for the plausible and extreme interaction effect sizes when assuming no effect of BMI on SARS-CoV-2 infection). As with the ALSPAC simulations, coverage varied greatly depending on the scenario.

**Table 3:**
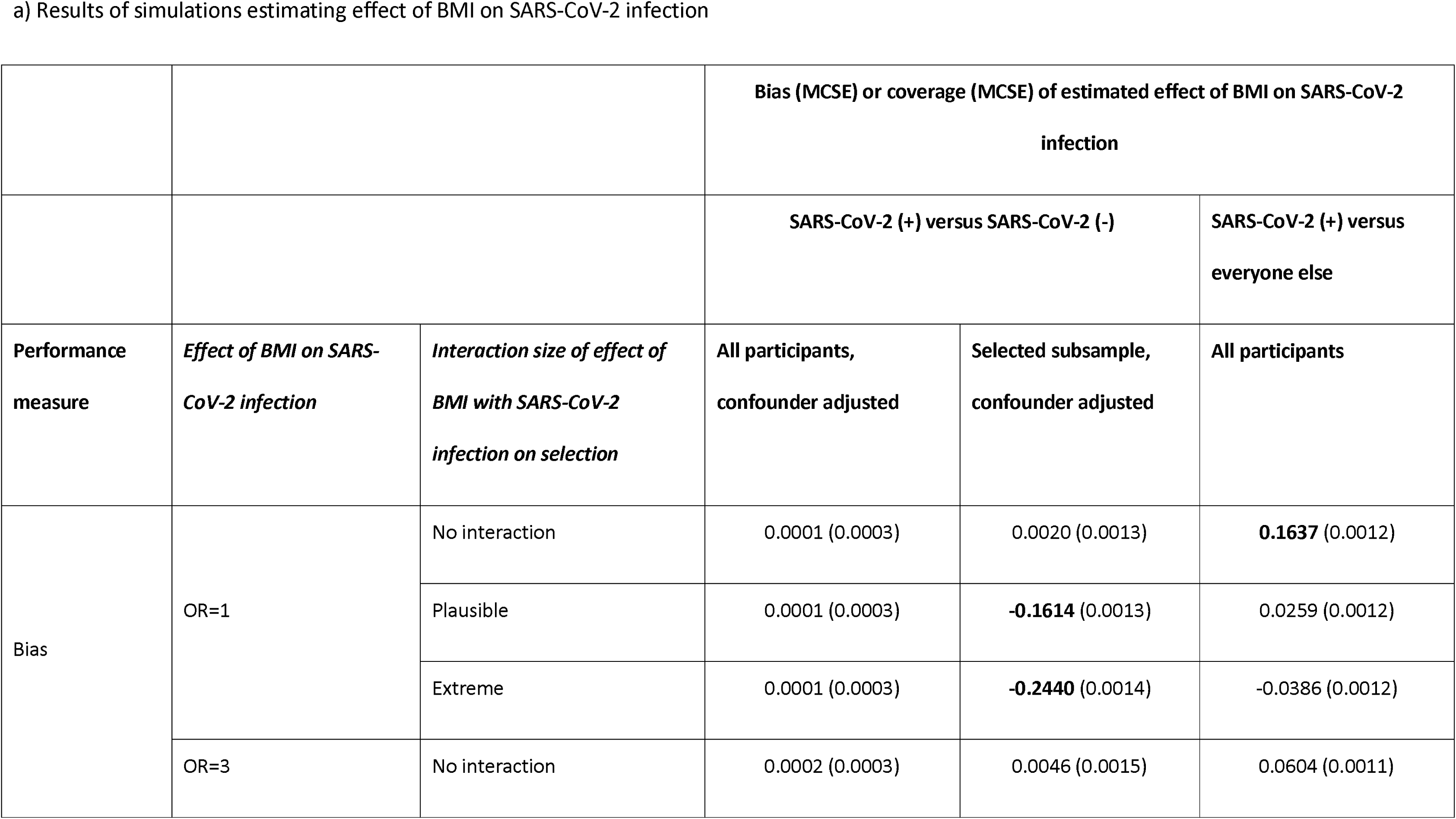
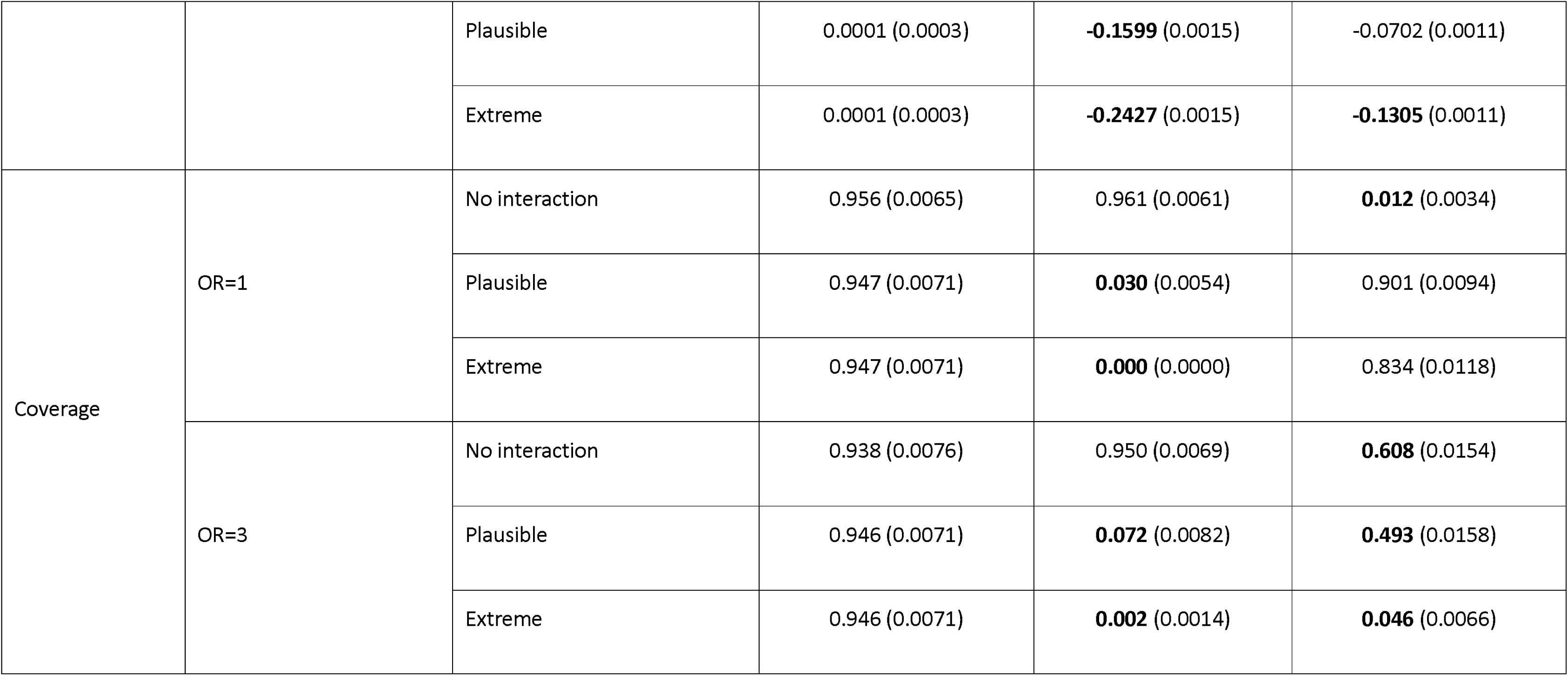
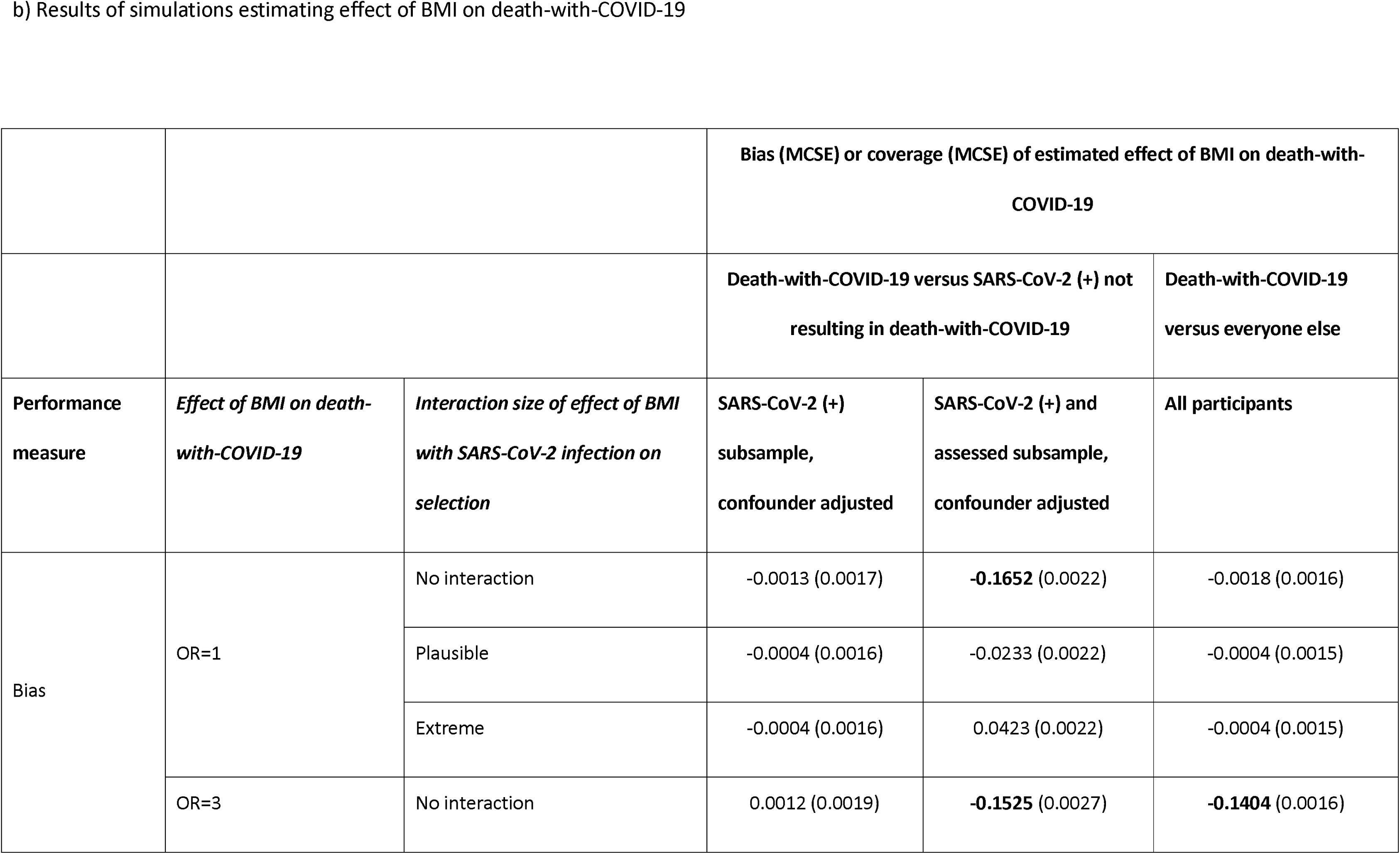
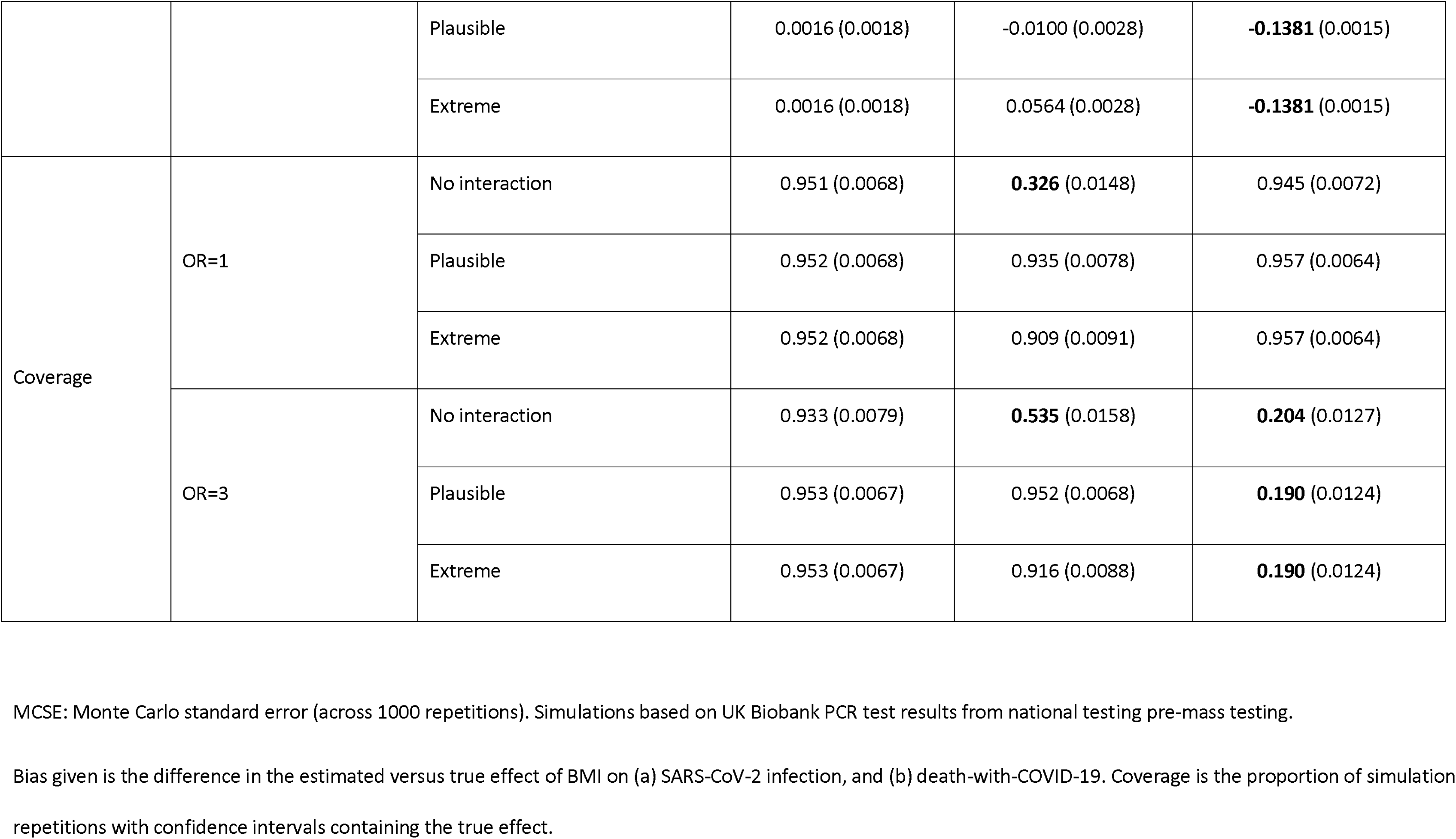
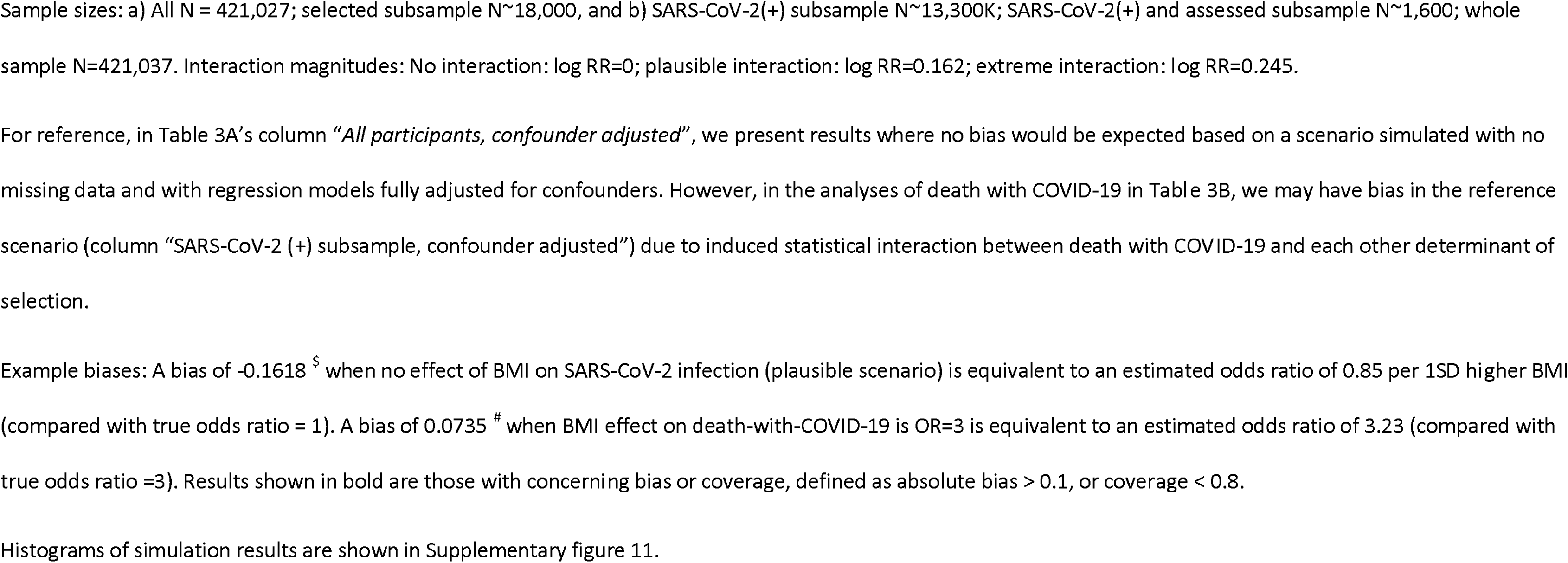
Results of simulations of SARS-CoV-2 infection and death-with-COVID-19 based on UK Biobank data.

Results of our simulations of death-with-COVID-19 are shown in Table 3b (histograms of estimates shown in **Supplementary figure 11b**). When using the death-with-COVID-19 vs SARS-CoV-2 (+) not resulting in death-with-COVID-19 outcome definition and assuming no interaction effect of BMI and SARS-CoV-2 infection on being tested (but with an interaction of death with COVID-19 with each other determinant of selection), estimates had negative bias and poor coverage (e.g. bias = -0.1652 [MCSE=0.0022] and 0.326 [MCSE=0.0148] coverage when assuming no effect of BMI on death-with-COVID-19). When including an interaction, bias became more positive and coverage improved (e.g. bias = -0.0233 (MCSE=0.0022) with 0.935 [MCSE=0.0078] coverage for the plausible scenario with no effect of BMI on death-with-COVID-19).

For the death-with-COVID-19 versus everyone else outcome we found little evidence of bias and good coverage when there was no effect of BMI on death-with-COVID-19 (e.g. bias = - 0.0004 [MCSE=0.0015], coverage=0.957 [MCSE=0.0064] for the ‘plausible’ interaction magnitude), and negative bias with poor coverage when BMI affected death-with-COVID-19 (OR=3) across the different interaction magnitudes (e.g. bias = -0.1381 [MCSE=0.0015], coverage=0.190 [MCSE=0.0124] for the ‘plausible’ interaction magnitude).

In general, the comparison group with least bias (and hence better coverage) depended on the particular assumptions used for the data generating mechanism. For example, when assuming no effect of SARS-CoV-2 infection and plausible interaction magnitude (for the effect of BMI and infection on infection), estimates of bias were comparable in the scenario based on ALSPAC data (bias=0.0517 [MCSE=0.0018] comparing SARS-CoV-2 (+) to SARS-CoV-2 (-) versus 0.0595 [MCSE=0.0015] comparing SARS-CoV-2 (+) to everyone else). In contrast, in the scenario based on UK Biobank the SARS-CoV-2 (+) versus SARS-CoV-2 (-) (selected subsample only) outcome definition had greater bias compared to the SARS-CoV-2 (+) versus everyone else definition (bias=-0.1614 [MCSE=0.0013] versus 0.0259 [MCSE=0.0012]).

Bias-eliminated coverage were all near to 0.95 (bias-eliminated coverage for all confounder adjusted estimates between 0.936 (MCSE=0.0077) and 0.963 (MCSE=0.0060)), confirming the poor coverage in some scenarios was driven solely by bias (**Supplementary tables 6 and 7**).

## SECTION 4: DISCUSSION

We found that a wide range of factors, including sociodemographic, behavioural, and health-related variables, were associated with being selected into the COVID-19 analytical subsamples in both ALSPAC and UKB. Some factors predicted selection in different directions and/or magnitudes between the two studies, possibly due to contrasting data collection mechanisms, characteristics of the target population or pre-pandemic selection pressures. In addition, we observed within-cohort differences in some factors predicting selection depending on the time of data collection and participant generations (in ALSPAC) considered.

As an example, we investigated the potential impact of selection on the association between BMI and COVID-19 outcomes in empirical analyses and simulations. In empirical analyses, estimates were imprecise in ALSPAC but suggested higher BMI was associated with higher odds of SARS-CoV-2 infection and death-with-COVID-19 in UKB. The magnitude of bias estimated in simulations varied widely depending on the specific data generating mechanism. The simulation results were used to see whether the empirical estimates can be explained by the bias seen in the simulated scenarios. For example, in UKB we estimated a larger association of BMI on SARS-CoV-2 infection using the SARS-CoV-2 (+) versus everyone else definition compared to SARS-CoV-2 (+) versus SARS-CoV-2 (-) (OR=1.26 [95% CI: 1.21-1.32] and OR=1.09 [95% CI: 1.03-1.16], respectively). In our ‘plausible’ simulation scenario assuming an effect of BMI on SARS-CoV-2 infection (OR=3) we found a smaller negative bias in the cases versus everyone, consistent with this (bias -0.16 [MCSE=0.0015] and -0.07 [MCSE=0.0011] respectively).

In general, the bias for the SARS-CoV-2 infection simulations for both comparison groups (i.e. SARS-CoV-2 (-) or everyone else) depends on the direction and magnitude of the interaction effect of BMI and infection on selection. For the everyone else comparison group, differential misclassification, where the SARS-CoV-2(+) non-assessed participants are included in the comparison group, means that the direction of the bias overall also depends on the BMI and infection distributions and the effects of selection across these. Furthermore, for the death-with-COVID-19 analyses, including all participants who died with COVID-19 statistically induces an interaction between death-with-COVID-19 and all other determinants of selection (e.g. BMI affects selection only in those who did not die with COVID-19 such the BMI’s effect on selection is modified by death-with-COVID-19), which also induces bias in the estimated effect of BMI on death-COVID-19. We provide further details and intuition for these biases in **Supplementary section 6** and **Supplementary figure 10**.

### Results in context

Previous studies have investigated factors predicting selection into COVID-19 studies in ALSPAC [17] and UKB [18, 23]. We previously reported a range of factors influencing selection into the COVID-19 analytical ALSPAC sample [17], which could be related to pre-pandemic loss to follow-up or non-participation in the COVID-19 questionnaires. Using UKB data, studies have reported a range of sociodemographic and health-related factors associated with being more likely to obtain a test for SARS-CoV-2 as well as to a higher risk of SARS-CoV-2 infection [18, 23]. We have built on this previous work to i) compare potential selection bias in an analytical sample (ALSPAC) with a different selection mechanism to UKB, ii) explore the potential presence of selection bias in analyses of COVID-19 death and iii) conduct simulations to explore the consequences of selection bias on empirical analyses.

In agreement with our findings, other studies have reported that higher BMI is associated with higher odds SARS-CoV-2 infection and COVID-19 disease severity [24–27]. However, our study indicates that the estimates reported in these studies may be impacted by selection bias.

### Strengths and limitations

We used both empirical analyses and simulations to comprehensively investigate the potential presence and impact of selection bias in COVID-19 studies. We used two cohorts with pre-pandemic data allowing us to identify potential determinants of selection. We were able to compare across these cohorts that have contrasting sources of COVID-19 data (from questionnaires in ALSPAC and national registries in UKB). In addition, a strength of our simulations is that we based most of the parameters on either cohort data or other secondary sources to try to reflect realistic scenarios.

In the analyses presented here we make several assumptions about or simplifications of the data. Both ALSPAC and UKB are subject to pre-pandemic selection bias due to non-random recruitment into these studies and loss to follow-up, which we do not account for here.

Overall, we considered misclassification of the comparison groups (e.g. infected as non-infected) but not of the case groups (e.g. non-infected as infected). This may be particularly problematic for self-reported COVID-19 data and cause of death attributed to COVID-19 early in the pandemic [23]. We have focussed analyses here on the first wave of the COVID-19 pandemic in the UK. Selection bias may change over time as the pandemic progresses, which may explain some of the differences between ALSPAC and UKB. In ALSPAC, the comparison of SARS-CoV-2 (+) with everyone else, including participants who did not reply to the questionnaires, may be unlikely to be conducted in practice, but is included here for completion (to be consistent with UKB analyses).

While we used parameters in the simulations that were estimated, where possible, using ALSPAC and UKB data, in some cases it was not possible to estimate some parameters with much degree of certainty. For example, we used smoking as a proxy for SARS-CoV-2 infection to identify possible magnitudes of the interaction between BMI and SARS-CoV-2 infection in their effect on selection, and estimating this interaction in ALSPAC and UKB meant these estimates, particularly in ALSPAC, were imprecise. In addition, in our simulations of death-with-COVID-19 we did not explore the potential impact of *index event bias*, which is a special case of selection bias that can occur when conditioning on disease incidence [19].

### Implications for COVID-19 research

A better understanding of risk and prognostic factors for COVID-19 will help to identify interventions to reduce the risk of SARS-CoV-2 infection and COVID-19 severity. Given logistic and ethical issues involving clinical trials, most evidence on risk and prognostic factors for the disease comes from observational studies [28]; however, teasing apart causal from non-causal relationships in such studies is notoriously difficult due to confounding, selection and measurement error. Our findings suggest that sample selection pressures can substantially differ between and within studies and can depend on a number of factors, such as the data collection mechanism, sample ascertainment, timepoint, geographical location, and characteristics of the target population. In addition, this study illustrates that bias due to sample selection and misclassification can distort relationships between risk/prognostic factors and disease in an unpredictable way. This indicates that there is no “one-size-fits-all” solution and that, where possible, individual studies should use longitudinal pre-pandemic data to thoroughly investigate whether and how selection pressures and misclassification may bias their results and to inform sensitivity analyses to mitigate these biases (e.g. inverse probability weighting, multiple imputation). Study specific sensitivity analyses and study-informed simulations can be used to explore the potential direction and magnitude of bias.

## Data Availability

The UK Biobank data underlying the results presented in the study are available from UK Biobank [www.ukbiobank.ac.uk] for researchers who meet the criteria for access to these confidential data. The ALSPAC data underlying the results presented in the study are available from ALSPAC [http://www.bristol.ac.uk/alspac/] for researchers who meet the criteria for access to these confidential data.

## Code Availability

All code for both the empirical cohort analysis and simulation study is available at [https://github.com/MRCIEU/COVIDITY_selbias/]. Git tag v0.1 corresponds to the version presented here.

## Funding

This work was conducted as part of the BHF National Institute of Health Research (NIHR) COVIDITY flagship project.

The UK Medical Research Council and Wellcome (Grant ref: 217065/Z/19/Z) and the University of Bristol provide core support for ALSPAC. This work was supported by the Wellcome Trust’s “Longitudinal Population Study Covid-19 Steering Group and Secretariat” (221574/Z/20/Z, a Strategic Support Science Grant); the Elizabeth Blackwell Institute for Research at the University of Bristol for the Questionnaire ‘COVID1’; and the University of Bristol Faculty Director’s Discretionary Fund for the Questionnaire ‘COVID2’. A comprehensive list of grant funding is available on the ALSPAC website (http://www.bristol.ac.uk/alspac/external/documents/grantacknowledgements.pdf).

This work was also supported by the Bristol British Heart Foundation (BHF) Accelerator Award (AA/18/7/34219, which supports AF-S and ARC ; the University of Bristol and Medical Research Council (MRC) Integrative Epidemiology Unit (MC_UU_00011/1, MC_UU_00011/3, MC_UU_00011/6, supporting LACM, AF-S, ARC, DM-S, GJG, GLC, GDS, DAL, KT, and MCB); RH and EK were supported by a Sir Henry Dale Fellowship jointly funded by the Wellcome Trust and the Royal Society (Grant Number 215408/Z/19/Z); the European Union’s Horizon 2020 research and innovation programme under grant agreement No 733206 (LifeCycle), which supports GLC; the University of Bristol (Vice-Chancellor’s Fellowships to LACM and MCB); the Economic and Social Research Council (ES/T009101/1, postdoctoral fellowship to GJG); the MRC (CH/F/20/90003, to DAL); the NIHR (NF-0616-10102, to DAL). TPM was funded by the Medical Research Council (MC_UU_12023/21, MC_UU_12023/29 and MC_UU_00004/07). The funders had no role in study design, data collection and analysis, decision to publish, or preparation of the manuscript. This research was funded in whole, or in part, by the Wellcome Trust. For the purpose of Open Access, the author has applied a CC BY public copyright licence to any Author Accepted Manuscript version arising from this submission.

This publication is the work of the authors and Louise AC Millard, Alba Fernández-Sanlés, Alice R Carter and Maria Carolina Borges will serve as guarantors for the contents of this paper.

## Acknowledgements

We are extremely grateful to all the families who took part in this study, the midwives for their help in recruiting them, and the whole ALSPAC team, which includes interviewers, computer and laboratory technicians, clerical workers, research scientists, volunteers, managers, receptionists and nurses.

We are extremely grateful to all the UK Biobank participants who took part in this study, and the whole study team. This work was carried out using UK Biobank project 16729.

## Author contributions

LACM, AF-S, ARC and MCB all drafted the manuscript. LACM conducted simulation analyses. AF-S conducted ALSPAC analyses. ARC conducted UKB analyses. All authors contributed to the design of the study. All authors critically reviewed and revised the manuscript.

## Declarations of Interest

TPM receives personal income from consulting for Kite Pharma, Inc. KT has acted as a consultant for CHDI foundation. DAL acknowledges support from Roche Diagnostics and Medtronic Ltd for research unrelated to that presented here. All other authors declare they have no conflict of interest, financial or otherwise.

## Notes

### Author Declarations

Ethical approval was obtained from the ALSPAC Ethics and Law Committee and the local research ethics committees. Informed consent for the use of data collected via questionnaires and clinics was obtained from participants following the recommendations of the ALSPAC Ethics and Law Committee at the time (details and reference numbers of all ethics approvals can be found at http://www.bristol.ac.uk/media-library/sites/alspac/documents/governance/Research%20Ethics%20Committee%20approval%20references.pdf). The work we present here was approved by the ALSPAC Ethics and Law Committee under project B3543. Ethical approval for UKB project 16729 was provided by UK Biobank, although no specific approval is required for analyses relating to COVID-19.

## REFERENCES

1. Smith LH. Selection Mechanisms and Their Consequences: Understanding and Addressing Selection Bias. Curr Epidemiol Reports. 2020;7:179–89.

2. Cole SR, Platt RW, Schisterman EF, Chu H, Westreich D, Richardson D, et al. Illustrating bias due to conditioning on a collider. Int J Epidemiol. 2010;39:417–20.

3. Munafò MR, Tilling K, Taylor AE, Evans DM, Davey Smith G. Collider scope: When selection bias can substantially influence observed associations. Int J Epidemiol. 2018;47:226–35.

4. Griffith GJ, Morris TT, Tudball MJ, Herbert A, Mancano G, Pike L, et al. Collider bias undermines our understanding of COVID-19 disease risk and severity. Nat Commun. Nature Research; 2020;11:5749.

5. van Smeden M, Lash TL, Groenwold RHH. Reflection on modern methods: five myths about measurement error in epidemiological research. Int J Epidemiol. 2020;49:338–47.

6. COVID-19 Host Genetics Initiative. Mapping the human genetic architecture of COVID-19. Nature. 2021;

7. Boyd A, Golding J, Macleod J, Lawlor DA, Fraser A, Henderson J, et al. Cohort Profile: The ‘Children of the 90s’—the indexoffspring of the Avon Longitudinal Study of Parents and Children. Int J Epidemiol [Internet]. Oxford University Press; 2013 [cited 2021 Nov 18];42:111. Available from: /pmc/articles/PMC3600618/

8. Fraser A, Macdonald-wallis C, Tilling K, Boyd A, Golding J, Davey smith G, et al. Cohort Profile: The Avon Longitudinal Study of Parents and Children: ALSPACmothers cohort. Int J Epidemiol [Internet]. Oxford University Press; 2013 [cited 2021 Nov 18];42:97. Available from: /pmc/articles/PMC3600619/

9. Northstone K, Lewcock M, Groom A, Boyd A, Macleod J, Timpson N, et al. The Avon Longitudinal Study of Parents and Children (ALSPAC): an update on the enrolled sample of index children in 2019 [version 1; peer review: 2 approved]. Wellcome Open Res. 2019;4:51.

10. Harris PA, Taylor R, Thielke R, Payne J, Gonzalez N, Conde JG. Research Electronic Data Capture (REDCap) - A metadata-driven methodology and workflow process for providing translational research informatics support. J Biomed Inform [Internet]. NIH Public Access; 2009 [cited 2021 Nov 18];42:377. Available from: /pmc/articles/PMC2700030/

11. Northstone K, Smith D, Bowring C, Wells N, Crawford M, Haworth S, et al. The Avon Longitudinal Study of Parents and Children - A resource for COVID-19 research: Questionnaire data capture May-July 2020. Wellcome Open Res. 2020;5:210.

12. Northstone K, Howarth S, Smith D, Bowring C, Wells N, Timpson NJ. The Avon Longitudinal Study of Parents and Children - A resource for COVID-19 research: Questionnaire data capture April-May 2020. Wellcome Open Res. 2020;5:127.

13. Fry A, Littlejohns TJ, Sudlow C, Doherty N, Adamska L, Sprosen T, et al. Comparison of Sociodemographic and Health-Related Characteristics of UK Biobank Participants With Those of the General Population. Am J Epidemiol. 2017;186:1026–34.

14. Baseline assessments [Internet]. Available from: https://www.ukbiobank.ac.uk/enable-your-research/about-our-data/baseline-assessment

15. World Health Organization. International guidelines for certification and classification (coding) of COVID-19 death [Internet]. 2020. Available from: https://cdn.who.int/media/docs/default-source/classification/icd/covid-19/guidelines-cause-of-death-covid-19-20200420-en.pdf?sfvrsn=35fdd864_2

16. Department of Health and Social Care. Everyone in the United Kingdom with symptoms now eligible for coronavirus tests [Internet]. 2021 [cited 2021 Nov 1]. Available from: https://www.gov.uk/government/news/everyone-in-the-united-kingdom-with-symptoms-now-eligible-for-coronavirus-tests?utm_source=932565f9-f9d7-45ec-b964-d9f353f71948&utm_medium=email&utm_campaign=govuk-notifications&utm_content=daily

17. Fernández-Sanlés A, Smith D, Clayton GL, Northstone K, Carter AR, Millard LAC, et al. Bias from questionnaire invitation and response in COVID-19 research: an example using ALSPAC [version 1; peer review: 1 approved]. Wellcome Open Res. 2021;6:184.

18. Chadeau-Hyam M, Bodinier B, Elliott J, Whitaker MD, Tzoulaki I, Vermeulen R, et al. Risk factors for positive and negative COVID-19 tests: A cautious and in-depth analysis of UK biobank data. Int J Epidemiol. Oxford University Press; 2020;49:1454–67.

19. Dahabreh IJ, Kent DM. Index event bias as an explanation for the paradoxes of recurrence risk research. JAMA. 2011;305:822–3.

20. Morris TP, White IR, Crowther MJ. Using simulation studies to evaluate statistical methods. Stat Med. 2019;38:2074–102.

21. Heinze G, Schemper M. A solution to the problem of separation in logistic regression. Stat Med. 2002;21:2409–19.

22. van Smeden M, de Groot JAH, Moons KGM, Collins GS, Altman DG, Eijkemans MJC, et al. No rationale for 1 variable per 10 events criterion for binary logistic regression analysis. BMC Med Res Methodol. 2016;16:163.

23. Griffith GJ, Davey Smith G, Manley D, Howe LD, Owen G. Interrogating structural inequalities in COVID-19 Mortality in England and Wales. J Epidemiol Community Health. 2021;jech-2021-216666.

24. Leong A, Cole JB, Brenner LN, Meigs JB, Florez JC, Mercader JM. Cardiometabolic risk factors for COVID-19 susceptibility and severity: A Mendelian randomization analysis. PLoS Med. 2021;18:e1003553.

25. Gao M, Piernas C, Astbury NM, Hippisley-Cox J, O’Rahilly S, Aveyard P, et al. Associations between body-mass index and COVID-19 severity in 6.9 million people in England: a prospective, community-based, cohort study. Lancet Diabetes Endocrinol. 2021;9:350–9.

26. Williamson EJ, Walker AJ, Bhaskaran K, Bacon S, Bates C, Morton CE, et al. Factors associated with COVID-19-related death using OpenSAFELY. Nature. 2020;584:430–6.

27. Recalde M, Pistillo A, Fernandez-Bertolin S, Roel E, Aragon M, Freisling H, et al. Body Mass Index and Risk of COVID-19 Diagnosis, Hospitalization, and Death: A Cohort Study of 2 524 926 Catalans. J Clin Endocrinol Metab. 2021;dgab546.

28. Raynaud M, Zhang H, Louis K, Goutaudier V, Wang J, Dubourg Q, et al. COVID-19-related medical research: a meta-research and critical appraisal. BMC Med Res Methodol. 2021;21:1.

