## Supplementary information for "Exploring selection bias in COVID-19 research: Simulations and prospective analyses of two UK cohort studies"

### SUPPLEMENTARY TEXT

#### Supplementary Section S1: Parameters using in simulation study

a) Parameters for simulations assessing the effect of BMI on SARS-CoV-2 infection

In order to investigate the direction and magnitude of selection bias in realistic settings, we estimated the association for each edge in our DAG using ALSPAC and UKB data whenever possible, and used these parameters in our simulations. We generated the distribution of the categorical confounders (sex, education level, smoking status and deprivation level) to match distributions in each cohort, and all continuous variables (age and BMI) are standardised (see Supplementary table 4 for each variable distribution).

We estimated the association of the confounders, which included age, sex, education, residential area deprivation (Townsend Deprivation Index – TDI – in UKB and the Index of Multiple Deprivation – IMD – in ALSPAC) and smoking status, jointly, with BMI in ALSPAC and UKB using multivariable linear regression, setting the intercept and random error of this model such that BMI was generated standardised (mean=0, SD=1). We estimated the association of BMI and the confounders jointly, with selection in UK Biobank and ALSPAC using Poisson regression (see Supplementary section 2 for an explanation of this). We assumed a risk ratio (RR)=5.80 and 5.05 in ALSPAC and UK Biobank respectively, for the effect of SARS-CoV-2 infection on selection in our main analyses (see Supplementary section S3 for a description of how these values were inferred). Generating selection with a Poisson model and including only main effects will not introduce selection bias, when estimating the effect of BMI on SARS-CoV-2 infection with logistic regression (see Supplementary section S2). We therefore included an interaction term, for the effect of BMI and SARS-CoV-2 infection on selection (updating the main effects accordingly so that the marginal probabilities remained constant, see Supplementary Section 4). Since we cannot estimate this interaction using cohort data (as we do not have SARS-CoV-2 infection status among those not assessed) we instead estimate the interaction between smoking status and BMI, in their effect on selection. Among those who do not smoke a higher BMI may cause people to be more likely to have a test, while those who do smoke often have a cough so are more likely (on average) to test themselves for a SARS-CoV-2 infection irrespective of their BMI. An interaction between BMI and SARS-CoV-2 infection may work in a similar way, so we estimate the interaction between BMI and smoking in their effect on being assessed, and use this as a proxy measure for the interaction of BMI with SARS-CoV-2 infection. In ALSPAC and UKB interaction terms were log RR=0.0527 [-0.030, 0.135] and log RR=-0.162 [-0.245, -0.079], respectively. We used these estimates as the interaction terms in our main analysis, and repeated the simulations with most extreme values in the confidence intervals in the same direction as the estimate (i.e. the upper CI for ALSPAC, and the lower CI for UK Biobank) as sensitivity analyses.

We estimated the association of the confounders jointly, with SARS-CoV-2 infection in ALSPAC and UKB, using the subsample of those participants who were assessed (i.e. had self-reported having had COVID-19 in ALSPAC or with a SARS-CoV-2 PCR test result in UKB), using logistic regression. We repeated the simulations assuming a) no effect of BMI on SARS-CoV-2 infection and b) OR=3 for the effect of BMI on SARS-CoV-2 infection.

We set the intercept of the models generating selection (i.e. assessed participants) such that the proportion of participants selected were consistent with proportions estimated in each cohort (19.97% of ALSPAC participants responded to COVID-19 questionnaire 1 and 4.33% of UKB participants had a SARS-CoV-2 PCR test pre-mass testing). We set the intercept of the model generating SARS-CoV-2 infection assuming a prevalence of SARS-CoV-2 infection of 3.16% in ALSPAC prior to COVID-19 questionnaire 1, and 7.20% in UKB prior to the start of mass testing [1]. These estimates of SARS-CoV-2 infection prevalence were extracted from the REal-time Assessment of Community Transmission-2 (REACT-2) national seroprevalence study using subgroup prevalences for England, during the first wave of the pandemic (i.e. until mid-July 2020), and considering the age groups that matched the age distribution of UKB (i.e. 65–74y) and ALSPAC G1 participants (i.e. 15–44y).

b) Parameters for simulations assessing the effect of BMI on death-with-COVID-19

We estimated the association between the confounders jointly with death-with-COVID-19 in UKB, in the subsample of participants who tested positive for SARS-CoV-2. We set the intercept of this model to assume an infection fatality ratio (% COVID-19 deaths among all with a SARS-CoV-19 infection) of 3.13%, as estimated by the REACT-2 national study for England during the first wave of the pandemic (i.e. until mid-July), among people aged 65–74y, excluding care home residents [1]. To generate the selection variable we first assigned those who were assessed and tested positive as selected. We then also assigned those who died with COVID-19 as selected into our sample. This assumes that there was no missingness of deaths with COVID-19, such that there is no selection bias among this subsample, but among those who did not die with COVID-19 there may be selection bias.

#### Supplementary Section S2: Overview of simulating selection bias

For a binary outcome *Y* where we are estimating the effect of an exposure *X* using logistic regression, selection bias can only occur in our estimated odds ratio if *X* and *Y* do not independently affect selection, *S*, i.e. the following holds:

$$P(S|X,Y)\neq P(S|X)\times P(S|Y)$$

If $P\left( S | X,Y \right)=P\left( S | X \right)\times P(S|Y$), then estimates in the selected subsample where S=1 will be unbiased. The extent of the bias depends on the magnitude of the deviation from independence, which itself is determined by the specific selection mechanism involved. We can test independence using Poisson regression.

Generating a selection variable with logistic regression (implicit bias)

When generating data in a simulation, if we use a logit model then we may implicitly add an interaction on the probability scale. This is because, to give no interaction, we need this formula to hold:

$$P(S|X,Y)=P(S|X)\times P(S|Y)$$

On the log scale this is given by:

$$\log P(S|X,Y)=\log(P(S|X))+log(P(S|Y))$$

However, the logit model is given by:

$$\log\frac{p}{1-p}=\alpha_{0}+\alpha_{X}X+\alpha_{Y}Y$$

It is this difference in the left-hand side of these models that means an interaction may be implicitly induced when generating data using a logit model.

The following Stata code shows an example, where we have two fair coins, and they both affect whether a bell *b* is rung. The Poisson regression shows that there is an interaction between x and y in their effect on b, even though our logit model doesn’t explicitly include an interaction term. Then, when we test the association of y on x in the whole sample versus the subsample where b=1, we encounter selection bias.

clear all

set seed 1234

local i = 0.5

set obs 100000

* x and y are coins

gen x = rbinomial(1, 0.5)

gen y = rbinomial(1, 0.5)

* whether the bell rings or not depends on x and y

gen p = invlogit(-0.1 + 10*x - 0.2*y)

gen b = rbinomial(1, p)

* check there are some in each cell

tab x y if b == 1

* check if there is an interaction between x and y for their

* effect on b

poisson b x##y

** log risk ratio estimates from poisson model:

** x estimate: 0.74 [95% CI : 0.72, 0.76]

** y estimate: -0.12 [95% CI : -0.15, -0.10]

** x*y estimate: 0.12 [95% CI : 0.09, 0.15]

****** BIASED ASSOCIATION IN SUBSAMPLE

* check unbiased in whole sample

logistic y x

** odds ratio x estimate: 1.01 [95% CI: 0.98, 1.03]

* association in subsample where bell has rung

logistic y x if b == 1

** odds ratio x estimate: 1.14 [95% CI: 1.10, 1.17]

Generating a selection variable with explicit bias

To generate a selection outcome without an interaction, we can use the Poisson model:

$$\log P(S|X,Y)=\alpha_{0}+\alpha_{X}X+\alpha_{Y}Y$$

We can see that this has the right form, so can be conveniently used to generate probabilities (and hence a binary variable sampled from these probabilities) satisfying the independence assumption.

We can also add an interaction term into the Poisson model, to explicitly deviate from independence:

$$\log P(S|X,Y)=\alpha_{0}+\alpha_{X}X+\alpha_{Y}Y+\delta XY$$

Each parameter in this model is the risk ratio of S per 1-unit higher covariate.

Here is some Stata code demonstrating this:

clear all

set seed 1234

set obs 10000

* x and y are coins

gen x = rbinomial(1, 0.5)

gen y = rbinomial(1, 0.5)

* whether the bell rings or not depends on x and y

gen logp = -0.4 - 0.1*x - 0.2*y

** or replace with this version to include an interaction:

* gen xy = x*y

* gen logp = -0.4 - 0.1*x - 0.2*y -0.2*xy

gen p = exp(logp)

gen b = rbinomial(1, p)

* check there are some in each cell

tab x y if b == 1

* check if there is an interaction between x and y for their effect on b

poisson b x##y

** log risk ratio estimates from poisson model:

** x estimate: -0.10 [95% CI : -0.17, -0.03]

** y estimate: -0.22 [95% CI : -0.29, -0.15]

** x*y estimate: -0.00 [95% CI : -0.10, 0.10]

* check unbiased in whole sample

logistic y x

** odds ratio x estimate: 0.96 [95% CI: 0.89, 1.04]

* association in subsample where bell has rung

logistic y x if b == 1

** odds ratio x estimate: 0.96 [95% CI: 0.87, 1.07]

#### Supplementary Section S3: Deriving a feasible effect of SARS-CoV-2 infection on selection

We use estimates of the infection prevalence and the proportion of asymptomatic cases, along with the proportion of those who tested positive of those tested in each cohort, to estimate a feasible risk ratio for the effect of SARS-CoV-2 infection on being assessed. The infection prevalences used were extracted from the REal-time Assessment of Community Transmission-2 (REACT-2) national seroprevalence study using subgroup prevalences in England, during the first wave of the pandemic (i.e. until mid-July 2020), and considering the age groups that matched the age distribution of UKB (i.e. 65–74y) and ALSPAC G1 participants (i.e. 15–44y), with values of 0.0316 and 0.0720 in ALSPAC and UK Biobank scenarios, respectively [1]. We assume 20% of SARS-CoV-2 infections are asymptomatic [2]. The proportion of SARS-CoV-2 (+) among those assessed was 0.159 in ALSPAC and 0.281 in UK Biobank.

Our calculations suggest feasible risk ratios of 5.80 and 5.05 for the effect of SARS-CoV-2 infection on being assessed, in ALSPAC and UK Biobank respectively. Supplementary file S1 shows derivations of these risk ratios.

#### Supplementary section S4: Description of models used in simulations

Simulation A: Effect of BMI on SARS-CoV-2 infection

We generated the distribution of the categorical confounders (sex, education level, smoking status and deprivation level) to match distributions in each cohort. Area deprivations was categorical in the ALSPAC simulation and continuous in the UK Biobank simulation to match the variables available in these studies. See Supplementary table 4 for details of the specific parameters used to generate these variables.

The remaining variables were generated under the following model:

$$bmi= \beta_{0}+ \beta_{age}age+ \beta_{sex}sex+ \beta_{smok\_prev}smok\_prev+ \beta_{smok\_curr}smok\_curr+ {\beta_{ed\_alevel}ed\_alevel+ \beta_{ed\_voc}ed\_voc+ \beta_{ed\_deg}ed\_deg+ \beta}_{depriv}depriv+\epsilon$$

$$logit\left( p\left( infection \right) \right)= \beta_{0}+ \beta_{bmi}bmi+ \beta_{age}age+ \beta_{sex}sex+ \beta_{smok_{prev}}smok_{prev}+ \beta_{smok_{curr}}smok_{curr}$$

$$+ {\beta_{ed\_alevel}ed\_alevel+ \beta_{ed\_voc}ed\_voc+ \beta_{ed\_deg}ed\_deg+ \beta}_{depriv}depriv$$

$$\log(p(assessed))= \beta_{0}+ \beta_{bmi}bmi+ \beta_{age}age+ \beta_{sex}sex+ \beta_{smok_{prev}}smok_{prev}+ \beta_{smok_{curr}}smok_{curr}$$

$$+ {\beta_{ed\_alevel}ed\_alevel+ \beta_{ed\_voc}ed\_voc+ \beta_{ed\_deg}ed\_deg+ \beta}_{depriv}depriv+ \beta_{infection}infection$$

For this simulation selected = assessed, i.e. all those who were assessed (either responding to the questionnaire in ALSPAC or having a test in UK Biobank) are selected.

Simulation B: Effect of BMI on COVID-19 severity (UK Biobank only)

The confounders, BMI and infection were generated as in simulation A. The remaining variables were generated under the following model:

$$\log it (p(coviddeathrisk))= \beta_{0}+ \beta_{bmi}bmi+ \beta_{age}age+ \beta_{sex}sex+ \beta_{smok_{prev}}smok_{prev}+ \beta_{smok_{curr}}smok_{curr}$$

$$+ {\beta_{ed\_alevel}ed\_alevel+ \beta_{ed\_voc}ed\_voc+ \beta_{ed\_deg}ed\_deg+ \beta}_{depriv}depriv+ \beta_{infection}infectio$$

$$coviddeath=\left\{ \begin{aligned} 1:coviddeathall=1 &and infection=1 \\ 0:other&wise&&& \end{aligned} \right.$$

$$selected= \left\{ \begin{aligned} 1:coviddeath=1 \\ 1:assessed=1 & and infection=1 \\ 0:otherwise \end{aligned} \right.$$

where *coviddeathrisk* is the risk of dying from covid if you were to become infected, coviddeathall is a binary variable denoting whether a participant would die *if* they get infected, and coviddeath is a binary variable denoting whether a participant actually died with COVID-19.

Specific parameter values for our simulations based on ALSPAC

$$bmi= -0.084+ 0.0044\times age+ 0.0409\times sex+ 0.1202\times smok_{prev}+ 0.0346\times smok_{curr}+ {-0.1502\times ed_{alevel}+ 0.1064\times ed\_voc+ -0.2645*\times ed\_deg+}0.0764\times depriv + \mathcal{N}(0,0.985)$$

*Infection model for scenario where BMI affects SARS-Cov-2 infection:*

$$logit\left( p\left( infection \right) \right)=-3.475+\log\left( 3 \right)\times bmi+ 0.0297\times age+ 0.1944\times sex+ 0.3287\times smok_{prev}+ 0.2191\times smok_{curr}$$

$$+ {0.3179\times ed\_alevel+ 0.3313\times ed\_voc+ 0.3436\times ed\_deg+}-0.0497\times depriv$$

*Infection model for scenario where BMI does not affect SARS-Cov-2 infection:*

$$logit\left( p\left( infection \right) \right)=-2.997+ 0.0297\times age+ 0.1944\times sex+ 0.3287\times smok_{prev}+ 0.2191\times smok_{curr}$$

$$+ {0.3179\times ed\_alevel+ 0.3313\times ed\_voc+ 0.3436\times ed\_deg+}-0.0497\times depriv$$

*Assessed model without interaction:*

$$\log(p(assessed))=-2.75+ 0.0208\times bmi+ 0.0162\times age+ -0.2689\times sex+ 0.0228\times smok_{prev}+ -0.0594\times smok_{curr}+0.3387\times ed_{alevel}$$

$$+ 0.1095\times ed\_voc+0.3408\times ed\_deg-0.0175\times depriv+ log(5.80)\times infection$$

*Assessed model with ‘plausible’ interaction:*

For those with no SARS-CoV-2 infection:

$$\log(p(assessed))=a_{0}+ 0.005\times bmi+ 0.0162\times age+ -0.2689\times sex+ 0.0228\times smok_{prev}+ -0.0594\times smok_{curr}+0.3387\times ed_{alevel}+ 0.1095\times ed_{voc}$$

$$+0.3408\times ed\_deg-0.0175\times depriv$$

For those with a SARS-CoV-2 infection:

$$\log(p(assessed))= a_{0}+ 0.0577\times bmi+ 0.0162\times age+ -0.2689\times sex+ 0.0228\times smok_{prev}+ -0.0594\times smok_{curr}+0.3387\times ed_{alevel}+ 0.1095\times ed_{voc}$$

$$+0.3408\times ed\_deg-0.0175\times depriv+ c$$

Where

$$a_{0}= A - b_{1}\times\mu_{0}$$

$$c=B - b_{2}\times\mu_{1}- a_{0}$$

Given that

$$A=a_{0}+ b'\times\mu_{0}$$

$$B=a_{0}+ c^{'}+b'\times\mu_{1}$$

Where b is the beta for BMI in the ‘no interaction’ model (0.0208 in this example), c’ is the beta for covid in the ‘no interaction’ model (log(5.80) in this case), and $b_{1}$and $b_{2}$ are the betas for BMI in the interaction, for no infection and infection, respectively (0.005 and 0.0577 in this case).

*Assessed model with ‘extreme’ interaction:*

For those with no SARS-CoV-2 infection:

$$\log(p(assessed))=a_{0}+ -0.019\times bmi+ 0.0162\times age+ -0.2689\times sex+ 0.0228\times smok_{prev}+ -0.0594\times smok_{curr}+0.3387\times ed_{alevel}+ 0.1095\times ed_{voc}$$

$$+0.3408\times ed\_deg-0.0175\times depriv$$

For those with a SARS-CoV-2 infection:

$$\log(p(assessed))= a_{0}+ 0.116\times bmi+ 0.0162\times age+ -0.2689\times sex+ 0.0228\times smok_{prev}+ -0.0594\times smok_{curr}+0.3387\times ed_{alevel}+ 0.1095\times ed_{voc}$$

$$+0.3408\times ed\_deg-0.0175\times depriv+ c$$

Where $a_{0}$ and c are calculated as above.

Specific parameter values for our simulations based on UK Biobank

$$bmi= 0.0106+ 0.0214\times age+ 0.1606\times sex+ 0.1049\times smok_{prev}+ 0.1074\times smok_{curr}+ -0.1561 \times ed\_alevel$$

$$+-0.0290\times ed_{voc}+ -0.2739\times ed_{deg}+0.0881\times depriv + \mathcal{N}(0,0.983)$$

*Infection model for scenario where BMI affects SARS-Cov-2 infection:*

$$logit\left( p\left( infection \right) \right)=-4.100+ \log\left( 3 \right)\times bmi-0.0702\times age+ 0.2838\times sex+ 0.0436\times smok_{prev}-0.2052 \times smok_{curr}- 0.2221\times ed_{alevel}$$

$$+ 0.1172\times ed\_voc-0.2043\times ed\_\deg+ 0.1231\times depriv$$

Infection model for *scenario where BMI does not affect SARS-Cov-2 infection:*

$$logit\left( p\left( infection \right) \right)=-3.526- 0.0702\times age+ 0.2838\times sex+ 0.0436\times smok_{prev}-0.2052 \times smok_{curr}- 0.2221\times ed_{alevel}+ 0.1172\times ed_{voc}$$

$$-0.2043\times ed\_\deg+ 0.1231\times depriv$$

*Assessed model without interaction:*

$$\log(p(assessed))=-4.610+ 0.1617\times bmi+ 0.1047\times age -0.1079\times sex+ 0.1625\times smok_{prev}+ 0.2835\times smok_{curr}-0.2084\times ed_{alevel}$$

$$-0.0105\times ed\_voc -0.1443\times ed\_\deg+0.2088\times depriv+ log(5.05)\times infection$$

*Assessed model with ‘plausible’ interaction:*

For those with no SARS-CoV-2 infection:

$$\log(p(assessed))=a_{0}+ 0.187\times bmi+ 0.1047\times age -0.1079\times sex+ 0.1625\times smok_{prev}+ 0.2835\times smok_{curr} -0.2084\times ed_{alevel}-0.0105\times ed_{voc}$$

$$-0.1443\times ed\_\deg+0.2088\times depriv$$

For those with a SARS-CoV-2 infection:

$$\log(p(assessed))=a_{0}+ 0.025\times bmi+ 0.1047\times age -0.1079\times sex+ 0.1625\times smok_{prev}+ 0.2835\times smok_{curr}-0.2084\times ed_{alevel}-0.0105\times ed_{voc}$$

$$-0.1443\times ed_{\deg+0}.2088\times depriv+c$$

Where $a_{0}$ and c are calculated as above.

*Assessed model with ‘extreme interaction:*

For those with no SARS-CoV-2 infection:

$$\log(p\left( assessed \right))= a_{0}+ 0.205\times bmi+ 0.1047\times age -0.1079\times sex+ 0.1625\times smok_{prev}+ 0.2835\times smok_{curr}-0.2084\times ed_{alevel}-0.0105\times ed_{voc}$$

$$-0.1443\times ed\_\deg+0.2088\times depriv$$

For those with a SARS-CoV-2 infection:

$$\log(p\left( assessed \right))=a_{0}-0.04\times bmi+ 0.1047\times age -0.1079\times sex+ 0.1625\times smok_{prev}+ 0.2835\times smok_{curr}-0.2084\times ed_{alevel}-0.0105\times ed_{voc}$$

$$-0.1443\times ed_{\deg+0}.2088\times depriv+c$$

Where $a_{0}$ and c are calculated as above.

#### Supplementary section S5: Simulation performance measures

We estimated the bias of the estimated effect of BMI on SARS-CoV-2 infection, compared to the true value, for each method described above:

$$\hat{bias}=\frac{1}{n_{sim}} \sum_{i=1}^{n_{sim}} (\hat{\theta}_{i}- \theta)$$

where $\hat{\theta}_{i}$ is the log odds of the estimated effect of BMI on SARS-CoV-2 infection of the i^th^ repetition, and $\theta$ is the true log odds effect (either log(1) or log(3)). The number of simulation repetitions $n_{sim}$ was set to 1000. We calculated the Monte Carlo standard error (SE) of the bias estimate as:

${SE}_{bias}= \sqrt{\frac{1}{n_{sim}(n_{sim}-1)} \sum_{i=1}^{n_{sim}} {{(\hat{\theta}}_{i}- \bar{\theta})}^{2}}$

Where $\bar{\theta}$ is the mean of $\hat{\theta}_{i}$ of all 1000 simulation repetitions.

We estimated the coverage of the confidence intervals:

$$\hat{cov}=\frac{1}{n_{sim}} \sum_{i=1}^{n_{sim}} {1(\hat{\theta}}_{low,i} \leq\theta\leq\hat{\theta}_{high,i})$$

Where $\hat{\theta}_{low,i}$ and $\hat{\theta}_{high,i}$ are the lower and upper bounds of the 95% confidence interval. We calculated the Monte Carlo standard error (SE) of coverage:

$${SE}_{coverage}= \sqrt{\frac{\hat{cov}\times(1-\hat{cov})}{n_{sim}}}$$

#### Supplementary section S6: Intuition of bias direction and magnitude

With an interaction, bias was positive in both ALSPAC and UKB because both these scenarios had a positive interaction term. In general bias for this outcome definition will be in the direction of, and proportional to, the risk ratio of this interaction (i.e. the interaction term). This is illustrated in Supplementary figure 3a and 3c. For the SARS-CoV-2 (+) vs everyone else outcome definition, for the ‘no interaction’ scenarios bias was induced due to the misclassification of cases as controls. The direction of this bias depended on the assumed effect of BMI on SARS-CoV-2 infection and the cohort underlying the simulation. In general, the bias for this ‘no interaction’ version depends on the distribution of participants across BMI and the degree of selection across these values, as illustrated in Supplementary figure 3b. When including an interaction term and using the SARS-CoV-2 (+) vs everyone else outcome definition, the bias became increasingly positive. This is because when both misclassification bias and selection bias are at play, the total bias depends on the magnitude of each of these, as illustrated in Supplementary figure 3b and 3d.

For the death-with-COVID-19 simulations based on UK Biobank the inclusion of all who had a death-with-COVID-19 in the analytic sample induced a negative bias. In general, including all with death with COVID-10 will induce negative bias (or less positive bias) when BMI has a positive effect on selection (and vice-versa) as illustrated in Supplementary figure 3e.

### Supplementary Tables

#### Supplementary Table 1: Definition of variables describing selection into COVID-19 subsamples in ALSPAC and UK Biobank

| **Candidate predictors of selection** | **Variables used** | | |
| --- | --- | --- | --- |
|  | **ALSPAC G1** | **ALSPAC G0 Mothers** | **UK Biobank** |
| *Sociodemographic factors* | | | |
| Age | Continuous | | |
|  | Age at the time of completion of the COVID-19 questionnaires in months | Age at the time of completion of the COVID-19 questionnaires in years | Age at baseline in years |
| Sex | Binary (male; female) | | |
|  | Recorded at birth/baseline | NA | Self-reported at baseline |
| Ethnicity | Binary (white; non-white) | | |
|  | Reported by mother in questionnaire C during pregnancy | Self-reported in questionnaire C during pregnancy | Self-reported at baseline  White (British, Irish or Other White background); all other ethnic backgrounds |
| Education level | Categorical  (GCSE/CSE/O level/Lower; Vocational; AS/A level; Degree/Higher) ^1^ | | |
|  | Self-reported in YPF questionnaire and during ALSPAC-G2^2^ enrolment (most recent used) | Self-reported in questionnaire C during pregnancy | Self-reported at baseline |
| Index of Deprivation | Ordinal (treated as continuous) | | Continuous |
|  | Quintiles of Index of Multiple Deprivation Score 2010 at timepoint Jan 2014 (1-5; 1, Least deprived; 5, Most deprived) | Quintiles of Index of Multiple Deprivation Score 2010 at timepoint Jan 2014 (1-5; 1, Least deprived; 5, Most deprived) | Townsend deprivation index of home postcode at baseline |
| Urban/Rural Index | Binary (Urban; Non-urban) | | |
|  | 2001 Census urban/rural indicator at timepoint Jan 2014 | 2001 Census urban/rural indicator at timepoint Jan 2014 | Urban or town; Rural (village or hamlet) based on baseline residence  Note: some values in the field relate to Scottish participants which are not included in these analyses |
| *Behavioural factors* | | | |
| Alcohol abuse | Binary (yes; no) | | |
|  | Yes (Mild (2-3 criteria) & Moderate (4-5 criteria) & Severe alcohol use disorder (≥6 criteria))  VS  No (0-1 criteria)  Alcohol Use Disorder (Diagnostic and Statistical Manual of Mental Disorders Fifth Edition [DSM-5]) since 2014 (“Focus@24” clinic and questionnaire “Life at 22+”) | Yes (hazardous (8-15) & harmful (16-19) & high (20-40) risk))  VS  No (Low (0-7))  Alcohol Use Disorders Identification Test (AUDIT), total score since 2010 (questionnaires T and V) | Yes (≥15 units per week)  VS  No (<15 units per week)  Units per week estimated from reported weekly consumption of beers, spirits and wine defined previously in [3] |
| Smoking status | Categorical (never smoked; former smoker; current smoker) | | |
|  | - Self-reported since 2012 (questionnaires “Your Changing Life”, “It’s All About You”, “Your Life Now (21+)”, “Me @ 23+”, “Life @ 24+”), and - Recorded from clinics since 2008 (“Teen Focus 4” and “Focus@24”)   Most recent record used. | Self-reported since 2010 (questionnaires T and V). Most recent record used. | Self-report smoking status at baseline |
| *Anthropometric factors* | | | |
| Body Mass Index (BMI) | Continuous (kg/m2) | | |
|  | Clinics since 2008 (“Teen Focus 4”, “Focus@24” and G2 clinic data). Most recent record used. | Clinics since 2008 (“Focus on Mothers 1”, “Focus on Mothers 2”, “Focus on Mothers 3” and “Focus on Mothers 4”). Most recent record used. | Baseline measures of height and weight to calculate BMI |
| Systolic Blood Pressure (SBP) | Continuous (mm Hg) | | |
|  | Clinics since 2008 (“Teen Focus 4”, “Focus@24” and G2 clinic data). Most recent record used. | Clinics since 2008 (“Focus on Mothers 1”, “Focus on Mothers 2”, “Focus on Mothers 3” and “Focus on Mothers 4”). Most recent record used. | Average of two automated baseline readings |
| Diastolic Blood Pressure (DBP) | Continuous (mm Hg) | | |
|  | Clinics since 2008 (“Teen Focus 4”, “Focus@24” and G2 clinic data). Most recent record used. | Clinics since 2008 (“Focus on Mothers 1”, “Focus on Mothers 2”, “Focus on Mothers 3” and “Focus on Mothers 4”). Most recent record used. | Average of two automated baseline readings |
| *Comorbidities* | | | |
| Cardiometabolic (ALSPAC) and  cardiovascular comorbidities (UKB) | Binary (yes; no) | | |
|  | Any diabetes, hypertension or cardiovascular disease since 2014   - self-reported in questionnaire “Life at 22+”, and - recorded from clinics “Teen Focus 4” and “Focus@24”” | Any diabetes, hypertension or cardiovascular disease   - self-reported since pregnancy (in questionnaires D, K, L, N, P, R, S, T and V), and - recorded from clinics since 2008 (“Focus on Mothers 1”, “Focus on Mothers 2”, “Focus on Mothers 3” and “Focus on Mothers 4”) | ICD9/ICD10 code for cardiovascular illness in hospital inpatient records |
| Hypertension | NA | | Binary (yes; no) |
|  | NA | NA | ICD-9/ICD-10 code for hypertension in hospital inpatient records |
| Respiratory comorbidities  (asthma and  lung-related disorders) | Binary (yes; no) | | |
|  | Asthma   - reported by mother or participant since 1999 (questionnaires “My Son / Daughter’s Well-being”, “Your Daughter/Son 16+ Years On”, “Life of a 16+ Teenager”, “Your Changing Life” and “Life at 22+”), and - recorded from clinics since 2005 (“Teen Focus 2” and “Teen Focus 3”) | Self-reported asthma and lung-related disorders since pregnancy (questionnaires D, N, R, S, T and VV) | ICD-9/ICD-10 code for respiratory disorders in hospital inpatient records |
| Autoimmune comorbidities | Binary (yes; no) | | |
|  | Self-reported in questionnaire “Life at 22+” | NA | ICD-9/ICD-1010 code for autoimmune disorders in hospital inpatient records |
| Cancer | Binary (yes; no) | | |
|  | NA | Self-reported since 1997 (questionnaires L, P, S, T and V) | All self-reported cancers at baseline or ICD-9/ICD-1010 code for cancer in hospital inpatient records |
| Adverse mental health outcomes  (depression, anxiety  and psychiatric disorders) | Binary (yes; no) | | |
|  | Any depression, anxiety or psychiatric disorder since 2013 (questionnaires “Plans and Aspirations: Online Survey”, “Your Changing Life”, “Your Life Now (21+)”, “Life at 22+”, “Me @ 23+” and “Life @ 25+”; and clinics “Teen Focus 4” and “Focus@24”)   - Short Mood and Feelings Questionnaire (SMFQ), total score (no depression if score ≤10; depression if score >10) - Generalised Anxiety Disorder 7-item Assessment (EPDS), total score (no generalised anxiety disorder if score ≤10; generalised anxiety disorder if score >10) - ICD-10 diagnosis of depression (Clinical Interview Schedule -Revised) - ICD-10 diagnosis of generalised anxiety disorder (Clinical Interview Schedule -Revised) - Rating of >=1 psychotic experience during psychosis-like symptom (PLIKS) interview | Any depression, anxiety or psychiatric disorder since 1999 (questionnaires N, R, S, T and V)   - self-reported depression, anxiety or psychiatric disorders - Edinburgh Postnatal Depression Scale (GAD-7), total score (no depression if score ≤11; depression if score >11) - State-trait Anxiety Inventory (STAI), total score (no anxiety if score ≤45; anxiety if score >45) | Self-reported seen a GP/psychiatrist for nerves, anxiety, tension or depression at baseline  Self-reported major depressive disorder or anxiety disorder or ICD-9/ICD-10 code for major depressive disorder or anxiety disorder in hospital inpatient records |

ICD-9: International Classification of Diseases, Ninth Revision; ICD-10: International Classification of Diseases, Tenth Revision.

^1^ GCSE, General Certificate of Secondary Education (mandatory qualifications taken at age 16, since 1988); CSE, Certificate of Secondary Education (qualifications taken at age 16 between 1965 and 1987; optional before 1973, but mandatory afterwards [unless taking O-levels]; O-level, Ordinary level qualifications (optional qualifications at age 16, between 1951 and 1987); Vocational qualifications (optional qualifications after age 16); AS/A-level, Advanced subsidiary/Advanced level qualifications (optional qualifications after age 16);

^2^ ALSPAC-G2: Second generation of ALSPAC (i.e. children of the original ALSPAC index child) not included in the current study.

#### Supplementary table 2: Distribution of characteristics of cohort study participants

**a) ALSPAC G1 participants**

| Variables | Whole sample | | | Selected subsample Q1 | | Selected subsample Q2 | | | | | |
| --- | --- | --- | --- | --- | --- | --- | --- | --- | --- | --- | --- |
|  | N | Mean (SD) / N (%) | | N | Mean (SD) / N (%) | N | | | | | Mean (SD) / N (%) |
| ***SARS-CoV-2/COVID-19 outcomes*** | | | | | | | | | | | |
| **Assessed for SARS-CoV-2 infection** | | | | | | | | | | | |
| Non-assessed | 14,849 | Q1: 11,883 (80.0%)  Q2: 12,145 (81.8%) | 2,966 | | 0 (0.0%) | | | 2,704 | | 0 (0.0%) | |
| Assessed |  | Q1: 2,966 (20.0%)  Q2: 2,704 (18.2%) |  |  | 2,966 (100.0%) | | |  |  | 2,704 (100.0%) | |
| **SARS-CoV-2 infection** | | | | | | | | | | | |
| *SARS-CoV-2 (+)* | 14,849 | 472 (3.2%)  478 (3.2%) | 2,966 | | 472 (15.9%) | | 2,704 | | 478 (17.7%) | | |
| *SARS-CoV-2 (-)* |  | 2,494 (16.8%)  2,226 (15.0%) |  |  | 2,494 (84.1%) | |  |  | 2,226 (82.3%) | | |
| ***Sociodemographic variables*** | | | | | | | | | | | |
| **Age** | 14,849 | Q1: 27.6 (0.5)  Q2: 27.8 (0.6) | | 2,966 | 27.6 (0.5) | 2,704 | | | | | 27.8 (0.6) |
| **Sex (female)** | 14,849 | 7,270 (49.0%) | | 2,966 | 2,122 (71.5%) | | 2,704 | | | | 1,910 (70.6%) |
| **Ethnicity** | | | | | | | | | | | |
| *White* | 12,045 | 11,436 (94.9%) | | 2,641 | 2,552 (96.6%) | 2,418 | | | | | 2,336 (96.6%) |
| *Other than White* |  | 609 (5.1%) | |  | 89 (3.4%) |  |  |  |  |  | 82 (3.4%) |
| **Education** | | | | | | | | | | | |
| *GCSE/none* | 3,885 | 529 (13.6%) | | 2,297 | 223 (9.7%) | 2,115 | | | | | 191 (9.0%) |
| *Vocational* |  | 581 (15.0%) | |  | 287 (12.5%) |  |  |  |  |  | 264 (12.5%) |
| *AS/A level* |  | 1,049 (27.0%) | |  | 671 (29.2%) |  |  |  |  |  | 637 (30.1%) |
| *Degree/higher* |  | 1,726 (44.4%) | |  | 1,116 (48.6%) |  |  |  |  |  | 1,023 (48.4%) |
| **Index of Multiple Deprivation** | | | | | | | | | | | |
| *Quintile 1 (least deprived)* | 13,023 | 3,889 (29.9%) | | 2,746 | 1,074 (39.1%) | 2,496 | | | | | 959 (38.4%) |
| *Quintile 2* |  | 2,971 (22.8%) | |  | 706 (25.7%) |  |  |  |  |  | 647 (25.9%) |
| *Quintile 3* |  | 2,354 (18.1%) | |  | 474 (17.3%) |  |  |  |  |  | 420 (16.8%) |
| *Quintile 4* |  | 2,156 (16.6%) | |  | 323 (11.8%) |  |  |  |  |  | 314 (12.6%) |
| *Quintile 5 (most deprived)* |  | 1,653 (12.7%) | |  | 169 (6.2%) |  |  |  |  |  | 156 (6.3%) |
| **Urban/Rural Index** | | | | | | | | | | | |
| *Urban* | 13,224 | 11,048 (83.6%) | | 2,779 | 2,226 (80.1%) | 2,529 | | | | | 2,031 (80.3%) |
| *Non-urban* |  | 2,176 (16.5%) | |  | 553 (19.9%) |  |  |  |  |  | 498 (19.7%) |
| ***Behavioural factors*** | | | | | | | | | | | |
| **Alcohol abuse** | | | | | | | | | | | |
| *No* | 5,155 | 4,464 (86.6%) | | 2,604 | 2,298 (88.3%) | 2,349 | | | | | 2,080 (88.6%) |
| *Mild* |  | 471 (9.1%) | |  | 220 (8.5%) |  |  |  |  |  | 199 (8.5%) |
| *Moderate* |  | 137 (2.7%) | |  | 51 (2.0%) |  |  |  |  |  | 39 (1.7%) |
| *Severe* |  | 83 (1.6%) | |  | 35 (1.3%) |  |  |  |  |  | 31 (1.3%) |
| **Smoking status** | | | | | | | | | | | |
| *Never smoker* | 7,468 | 2,744 (36.7%) | | 2,876 | 1,111 (38.6%) | 2,619 | | | | | 1,034 (39.5%) |
| *Former smoker* |  | 2,481 (33.2%) | |  | 1,041 (36.2%) |  |  |  |  |  | 937 (35.8%) |
| *Current smoker* |  | 2,243 (30.0%) | |  | 724 (25.2%) |  |  |  |  |  | 648 (24.7%) |
| ***Anthropometric factors*** | | | | | | | | | | | |
| **BMI** | 6,069 | 24.4 (5.1) | | 2,575 | 24.7 (5.3) | 2,338 | | | | | 24.7 (5.3) |
| **SBP** | 5,842 | 117.1 (11.5) | | 2,536 | 115.3 (10.8) | 2,304 | | | | | 115.2 (10.9) |
| **DBP** | 5,842 | 66.0 (7.8) | | 2,536 | 66.7 (7.8) | 2,304 | | | | | 66.9 (7.8) |
| ***Comorbidities*** | | | | | | | | | | | |
| **Cardiometabolic disorders** | | | | | | | | | | | |
| *Any* | 6,585 | 202 (3.1%) | | 2,755 | 92 (3.3%) | 2,499 | | | | | 86 (3.4%) |
| *None* |  | 6,383 (96.9%) | |  | 2,663 (96.7%) |  |  |  |  |  | 2,413 (96.6%) |
| **Respiratory disorders** | | | | | | | | | | | |
| *Any* | 10,088 | 3,140 (31.1%) | | 2,847 | 926 (32.5%) | 2,600 | | | | | 873 (33.6%) |
| *None* |  | 6,948 (68.9%) | |  | 1,921 (67.5%) |  |  |  |  |  | 1,727 (66.4%) |
| **Autoimmune disorders** | | | | | | | | | | | |
| *Any* | 3,994 | 80 (2.0%) | | 2,198 | 35 (1.6%) | 1,990 | | | | | 37 (1.9%) |
| *None* |  | 3,914 (98.0%) | |  | 2,163 (98.4%) |  |  |  |  |  | 1,953 (98.1%) |
| **Adverse mental health outcomes** | | | | | | | | | | | |
| *Any* | 7,494 | 3,344 (44.6%) | | 2,887 | 1,488 (51.5%) | 2,626 | | | | | 1,383 (52.7%) |
| *None* |  | 4,150 (55.4%) | |  | 1,399 (48.5%) |  |  |  |  |  | 1,243 (47.3%) |

The distribution of the COVID-19 related variables in ALSPAC is given for the self-reported data in each COVID-19 questionnaire. The distribution of the candidate predictors of selection is given for the whole cohorts and for the selected subsample with SARS-CoV-2 data from each questionnaire. Alcohol abuse in the ALSPAC G1 cohort was assessed as the DSM-5 alcohol use disorder symptoms (no, mild, moderate, severe).

**b) ALSPAC G0 Mothers**

| Variables | Whole sample | | | Selected subsample Q1 | | Selected subsample Q2 | | | | | |
| --- | --- | --- | --- | --- | --- | --- | --- | --- | --- | --- | --- |
|  | N | Mean (SD) / N (%) | | N | Mean (SD) / N (%) | N | | | | | Mean (SD) / N (%) |
| ***SARS-CoV-2/COVID-19 outcomes*** | | | | | | | | | | | |
| **Assessed for SARS-CoV-2 infection** | | | | | | | | | | | |
| Non-assessed | 14,282 | Q1:  11,597  (81.2%)  Q2:  11,650  (81.6%) | 2,685 | | 0 (0.0%) | | | 2,632 | | 0 (0.0%) | |
| Assessed |  | Q1: 2,685 (18.8%)  Q2: 2,632 (18.4%) |  |  | 2,685 (100.0%) | | |  |  | 2,632 (100.0%) | |
| **SARS-CoV-2 infection** | | | | | | | | | | | |
| *SARS-CoV-2 (+)* | 14,282 | Q1: 347 (2.4%)  Q2: 332 (2.3%) | 2,685 | | 347 (12.9%) | | 2,632 | | 332 (12.6%) | | |
| *SARS-CoV-2 (-)* |  | Q1: 2,338 (16.4%)  Q2:2,300 (16.1%) |  |  | 2,338 (87.1%) | |  |  | 2,300 (87.4%) | | |
| ***Sociodemographic variables*** | | | | | | | | | | | |
| **Age** | 14,267 | Q1: 56.1 (5.0)  Q2: 56.3 (5.0) | | 2,685 | 57.9 (4.4) | 2,632 | | | | | 58.0 (4.4) |
| **Ethnicity** | | | | | | | | | | | |
| *White* | 12,025 | 11,708 (97.4%) | | 2,551 | 2,511 (98.4%) | 2,511 | | | | | 2,471 (98.4%) |
| *Other than White* |  | 317 (2.6%) | |  | 40 (1.6%) |  |  |  |  |  | 40 (1.6%) |
| **Education** | | | | | | | | | | | |
| *CSE/O level/none* | 12,114 | 6,634 (54.8%) | | 2,557 | 1,036 (40.5%) | 2,516 | | | | | 1,017 (40.4%) |
| *Vocational* |  | 1,198 (9.9%) | |  | 149 (5.8%) |  |  |  |  |  | 140 (5.6%) |
| *A level* |  | 2,720 (22.5%) | |  | 761 (29.8%) |  |  |  |  |  | 760 (30.2%) |
| *Degree* |  | 1,562 (9.6%) | |  | 611 (23.9%) |  |  |  |  |  | 599 (23.8%) |
| **Index of Multiple Deprivation** | | | | | | | | | | | |
| *Quintile 1 (least deprived)* | 12,661 | 3,867 (30.5%) | | 2,489 | 1,046 (42.0%) | 2,450 | | | | | 1,038 (42.4%) |
| *Quintile 2* |  | 2,885 (22.8%) | |  | 663 (26.6%) |  |  |  |  |  | 658 (26.9%) |
| *Quintile 3* |  | 2,245 (17.7%) | |  | 410 (16.5%) |  |  |  |  |  | 400 (16.3%) |
| *Quintile 4* |  | 2,067 (16.3%) | |  | 253 (10.2%) |  |  |  |  |  | 241 (9.8%) |
| *Quintile 5 (most deprived)* |  | 1,597 (12.6%) | |  | 117 (4.7%) |  |  |  |  |  | 113 (4.6%) |
| **Urban/Rural Index** | | | | | | | | | | | |
| *Urban* | 12,866 | 10,678 (83.0%) | | 2,530 | 1,952 (77.2%) | 2,486 | | | | | 1,922 (77.3%) |
| *Non-urban* |  | 2,188 (17.0%) | |  | 578 (22.8%) |  |  |  |  |  | 564 (22.7%) |
| ***Behavioural factors*** | | | | | | | | | | | |
| **Alcohol abuse** | | | | | | | | | | | |
| *Low risk* | 4,740 | 2,444 (51.6%) | | 2,137 | 1,067 (49.9%) | 2,126 | | | | | 1,065 (50.1%) |
| *Hazardous* |  | 2,140 (45.1%) | |  | 1,004 (47.0%) |  |  |  |  |  | 994 (46.8%) |
| *Harmful* |  | 104 (2.2%) | |  | 46 (2.2%) |  |  |  |  |  | 47 (2.2%) |
| *High risk* |  | 52 (1.1%) | |  | 20 (0.9%) |  |  |  |  |  | 20 (0.9%) |
| **Smoking status** | | | | | | | | | | | |
| *Never smoker* | 5,269 | 2,766 (52.5%) | | 2,346 | 1,306 (55.7%) | 2,327 | | | | | 1,287 (55.3%) |
| *Former smoker* |  | 1,906 (36.2%) | |  | 839 (35.8%) |  |  |  |  |  | 849 (36.5%) |
| *Current smoker* |  | 597 (11.3%) | |  | 201 (8.6%) |  |  |  |  |  | 191 (8.2%) |
| ***Anthropometric factors*** | | | | | | | | | | | |
| **BMI** | 4,890 | 26.8 (5.5) | | 2,152 | 26.3 (5.0) | 2,138 | | | | | 26.3 (5.1) |
| **SBP** | 4,828 | 119.9 (14.3) | | 2,140 | 119.4 (14.1) | 2,128 | | | | | 119.3 (14.1) |
| **DBP** | 4,828 | 71.4 (9.6) | | 2,140 | 70.6 (9.3) | 2,128 | | | | | 70.5 (9.3) |
| ***Comorbidities*** | | | | | | | | | | | |
| **Cardiometabolic disorders** | | | | | | | | | | | |
| *Any* | 13,021 | 3,245 (24.9%) | | 2,684 | 722 (26.9%) | 2,631 | | | | | 719 (27.3%) |
| *None* |  | 9,776 (75.1%) | |  | 1,962 (73.1%) |  |  |  |  |  | 1,912 (72.7%) |
| **Respiratory disorders** | | | | | | | | | | | |
| *Any* | 12,765 | 2,461 (19.3%) | | 2,679 | 625 (23.3%) | 2,625 | | | | | 613 (23.4%) |
| *None* |  | 10,304 (80.7%) | |  | 2,054 (76.7%) |  |  |  |  |  | 2,012 (76.6%) |
| **Cancer** | | | | | | | | | | | |
| *Any* | 9,908 | 491 (5.0%) | | 2,658 | 174 (6.5%) | 2,607 | | | | | 177 (6.8%) |
| *None* |  | 9,417 (95.0%) | |  | 2,484 (93.5%) |  |  |  |  |  | 2,430 (93.2%) |
| **Adverse mental health outcomes** | | | | | | | | | | | |
| *Any* | 9,165 | 4,168 (45.5%) | | 2,647 | 1,333 (50.4%) | 2,599 | | | | | 1,324 (50.9%) |
| *None* |  | 4,997 (54.5%) | |  | 1,314 (49.6%) |  |  |  |  |  | 1,275 (49.1%) |

The distribution of the COVID-19 related variables in ALSPAC is given for the self-reported data in each COVID-19 questionnaire. The distribution of the candidate predictors of selection is given for the whole cohorts and for the selected subsample with SARS-CoV-2 data from each questionnaire. Alcohol abuse in the G0 mothers cohort as AUDIT groups (low risk, hazardous, harmful, high risk).

**c) UK Biobank participants**

|  | Full eligible UK Biobank | | | | Assessed participants | | | |
| --- | --- | --- | --- | --- | --- | --- | --- | --- |
|  | Pre-mass testing | | Post-mass testing | | Pre-mass testing | | Post-mass testing | |
| Variable | N | Mean (SE)/Frequency (%) | N | Mean (SE)/Frequency (%) | N | Mean (SE)/Frequency (%) | N | Mean (SE)/Frequency (%) |
| ***SARS-CoV-2/COVID-19 outcomes*** | | | | | | | | |
| **Assessed for SARS-CoV-2 infection** | | | | | | | | |
| *Not assessed* | 421,037 | 416,168 (98.84) | 419,226 | 362,610 (86.5) | 4,869 | 0 (0%) | 56,616 | 0 (0%) |
| *Assessed* |  | 4,869 (1.16) |  | 56,616 (13.5) |  | 4,869 (100%) |  | 56,616 (100%) |
| **SARS-CoV-2 infection** | | | | | | | | |
| *Negative* | 421,037 | 419,597 (99.66) | 419,226 | 406,252 (96.91) | 4,869 | 3,429 (70.43) | 56,616 | 43,642 (77.08) |
| *Positive* |  | 1,440 (0.34) |  | 12,974 (3.09) |  | 1,440 (29.57) |  | 12,974 (22.92) |
| **Severe COVID-19** | | | | | | | | |
| *No death-with-COVID-19* | 421,037 | 420,695 (99.92) | 419,226 | 418,978 (99.94) | 4,869 | 4,527 (92.98) | 56,616 | 56,368 (99.56) |
| *Death-with-COVID-19* |  | 342 (0.08) |  | 248 (0.06) |  | 342 (7.02) |  | 248 (0.44) |
| ***Sociodemographic variables*** | | | | | | | | |
| **Age at baseline** | 421,037 | 56.28 (8.09) | 419,226 | 56.26 (8.09) | 4,869 | 57.12 (8.85) | 56,616 | 56.87 (8.29) |
| **Sex** | | | | | | | | |
| *Female* | 421,037 | 231,984 (55.10) | 419,226 | 231,209 (55.15) | 4,869 | 2,496 (51.26) | 56,616 | 30,313 (53.54) |
| *Male* |  | 189,053 (44.90) |  | 188,017 (44.85) |  | 2,373 (48.74) |  | 26,303 (46.46) |
| **Ethnicity** | | | | | | | | |
| *White ethnicity* | 421,037 | 393,816 (93.53) | 419,226 | 392,113 (93.53) | 4,869 | 4,410 (90.57) | 56,616 | 52,546 (92.81) |
| *non-white ethnicity* |  | 27,221 (6.47) |  | 27,113 (6.47) |  | 459 (9.43) |  | 4,070 (7.19) |
| **Education** | | | | | | | | |
| *GCSE/O level or less* | 413,333 | 137,367 (33.23) | 411,578 | 136,550 (33.18) | 4,744 | 1,795 (37.84) | 55,427 | 20,211 (36.46) |
| *AS/A level* |  | 22,360 (5.41) |  | 22,289 (5.42) |  | 210 (4.43) |  | 2,823 (5.09) |
| *Vocational qualifications* |  | 115,874 (28.03) |  | 115,423 (28.04) |  | 1,387 (29.24) |  | 16,055 (28.97) |
| *Degree level or higher* |  | 137,732 (33.32) |  | 137,316 (33.36) |  | 1,352 (28.5) |  | 16,338 (29.48) |
| ***Townsend deprivation index*** | 420,540 | -1.33 (3.05) | 418,732 | -1.33 (3.05) | 4,861 | -0.53 (3.39) | 56,540 | -1.12 (3.17) |
| **Urban/rural index** | | | | | | | | |
| *Urban* | 416,989 | 386,520 (92.69) | 415,194 | 384,822 (92.68) | 4,830 | 4,608 (95.4) | 56,107 | 52,216 (93.07) |
| *Rural* |  | 30,469 (7.31) |  | 30,372 (7.32) |  | 222 (4.6) |  | 3,891 (6.93) |
| ***Behavioural factors*** | | | | | | | | |
| **Alcohol consumption** | | | | | | | | |
| *<15 units of alcohol per week* | 421,037 | 253,775 (60.27) | 419,226 | 252,726 (60.28) | 4,869 | 3,037 (62.37) | 56,616 | 34,255 (60.5) |
| *≥15 units of alcohol per week* |  | 167,262 (39.73) |  | 166,500 (39.72) |  | 1,832 (37.63) |  | 22,361 (39.5) |
| **Smoking status** | | | | | | | | |
| *Never smoker* | 418,690 | 234,557 (56.02) | 416,898 | 233,795 (56.08) | 4,832 | 2,363 (48.9) | 56,202 | 29,460 (52.42) |
| *Former smoker* |  | 142,740 (34.09) |  | 141,995 (34.06) |  | 1,846 (38.2) |  | 20,597 (36.65) |
| *Current smoker* |  | 41,393 (9.89) |  | 41,108 (9.86) |  | 623 (12.89) |  | 6,145 (10.93) |
| ***Physical measures*** | | | | | | | | |
| **BMI** | 418,599 | 27.36 (4.75) | 416,818 | 27.36 (4.75) | 4,809 | 28.42 (5.44) | 56,179 | 27.92 (4.99) |
| **SBP** | 399,170 | 137.44 (18.5) | 397,470 | 137.42 (18.49) | 4,561 | 137.97 (19.37) | 53,495 | 137.73 (18.64) |
| **DBP** | 399,177 | 82.11 (10.08) | 397,477 | 82.11 (10.08) | 4,561 | 82.11 (10.3) | 53,496 | 82.14 (10.15) |
| ***Comorbidities*** | | | | | | | | |
| **Cardiometabolic disorders** | | | | | | | | |
| *None* | 421,037 | 235,735 (55.99) | 419,226 | 235,476 (56.17) | 4,869 | 1,413 (29.02) | 56,616 | 23,043 (40.7) |
| *Any* |  | 185,302 (44.01) |  | 183,750 (43.83) |  | 3,456 (70.98) |  | 33,573 (59.3) |
| **Hypertension** | | | | | | | | |
| *Yes* | 421,037 | 264,627 (62.85) | 419,226 | 264,023 (62.98) | 4,869 | 2,196 (45.1) | 56,616 | 29,733 (52.52) |
| *No* |  | 156,410 (37.15) |  | 155,203 (37.02) |  | 2,673 (54.9) |  | 26,883 (47.48) |
| **Respiratory disorders** | | | | | | | | |
| *Yes* | 421,037 | 319,384 (75.86) | 419,226 | 318,692 (76.02) | 4,869 | 2,480 (50.93) | 56,616 | 37,819 (66.8) |
| *No* |  | 101,653 (24.14) |  | 100,534 (23.98) |  | 2,389 (49.07) |  | 18,797 (33.2) |
| **Autoimmune disorders** | | | | | | | | |
| *None* | 421,037 | 365,411 (86.79) | 419,226 | 364,081 (86.85) | 4,869 | 3,763 (77.28) | 56,616 | 46,398 (81.95) |
| *Any* |  | 55,626 (13.21) |  | 55,145 (13.15) |  | 1,106 (22.72) |  | 10,218 (18.05) |
| **Cancer** | | | | | | | | |
| *None* | 421,037 | 339,880 (80.72) | 419,226 | 339,045 (80.87) | 4,869 | 3,244 (66.63) | 56,616 | 40,922 (72.28) |
| *Any* |  | 81,157 (19.28) |  | 80,181 (19.13) |  | 1,625 (33.37) |  | 15,694 (27.72) |
| **Adverse mental health outcomes** | | | | | | | | |
| *None* | 421,037 | 260,750 (61.93) | 419,226 | 259,753 (61.96) | 4,869 | 2,633 (54.08) | 56,616 | 32,658 (57.68) |
| *Any* |  | 160,287 (38.07) |  | 159,473 (38.04) |  | 2,236 (45.92) |  | 23,958 (42.32) |

The distributions of the COVID-19 related variables and the candidate predictors of selection in UKBB are given separately for the pre-mass testing and post-mass testing periods, for the whole cohorts and for the selected subsamples. The deprivation index presented for UKBB is the Townsend deprivation Index.

#### Supplementary Table 3: Missing data in the candidate predictors of selection

| Variables | ALSPAC - G1  N missing (%)  (N_assessed_ = 14,849) | ALSPAC – G0 Mothers  N missing (%)  (N_assessed_ = 14,282) | UK Biobank  N missing (%)  (N_assessed_ = 421,037) |
| --- | --- | --- | --- |
| *Sociodemographic factors* | | | |
| Age | 0 | 15 (0.11%) | 0 |
| Sex | 0 | NA | 0 |
| Ethnicity | 2,804 (18.9%) | 2,257 (15.8%) | 0 |
| Education | 10,964 (73.8%) | 2,168 (15.2%) | 7,704 (1.8%) |
| Deprivation Index | 1,826 (12.3%) | 1,621 (11.4%) | 497 (0.1%) |
| Urban/Rural Index | 1,625 (10.9%) | 1,416 (9.9%) | 4,048 (1.0%) |
| *Behavioural factors* | | | |
| Alcohol abuse | 9,694 (65.3%) | 9,542 (66.8%) | 0 |
| Smoking status | 7,381 (49.7%) | 9,013 (63.1%) | 2,347 (0.6%) |
| *Anthropometric factors* | | | |
| BMI | 8,780 (59.1%) | 9,392 (65.8%) | 2,438 (0.6%) |
| SBP | 9,007 (60.7%) | 9,454 (66.2%) | 21,867 (5.2%) |
| DBP | 9,007 (60.7%) | 9,454 (66.2%) | 21,860 (5.2%) |
| *Comorbidities* | | | |
| Cardiometabolic disorders | 8,264 (55.6%) | 1,261 (8.8%) | 0 |
| Respiratory disorders | 4,761 (32.1%) | 1,517 (10.6%) | 0 |
| Autoimmune disorders | 10,855 (73.1%) | NA | 0 |
| Cancer | NA | 4,374 (30.6%) | 0 |
| Adverse mental health outcomes | 7,355 (49.5%) | 5,117 (35.8%) | 0 |

#### Supplementary Table 4: Data generating mechanism used in simulations

**a) Simulation of the association of BMI with SARS-CoV-2 infection**

| **Exposure** | **Outcome** | **Simulated effect sizes** |
| --- | --- | --- |
| BMI | SARS-CoV-2 infection | *Two versions: no effect, and OR=3.* ^1^ |
| Confounders | SARS-CoV-2 infection | Estimated in cohort in assessed^2^ subsample. Logistic regression of SARS-CoV-2 infection (positive versus negative test results) on confounders. ^1^ |
| Confounders | BMI | Estimated in cohort in whole sample. Linear regression of BMI on confounding factors. |
| Confounders and BMI | Assessed | Estimated in cohort in whole sample. Logistic regression of selection on confounders and BMI. ^3^ |
| SARS-CoV-2 infection | Assessed | *RR=5.80 in ALSPAC and 5.05 in UKB* |
| SARS-CoV-2 infection and BMI interaction | Assessed | *RR=5.80 in ALSPAC and 5.05 in UKB* |

**b) Simulation of the association of BMI with death-with-COVID-19 (UK Biobank only)**

| **Exposure** | **Outcome** | **Simulated effect sizes** |
| --- | --- | --- |
| Confounders | SARS-CoV-2 infection | As above. |
| Confounders and BMI | Assessed | As above. |
| Confounders | BMI | As above. |
| SARS-CoV-2 infection | Assessed | As above. |
| SARS-CoV-2 infection and BMI interaction | Assessed | As above. |
| Death-with-COVID-19 * | Assessed | Deterministic: All participants who died with COVID-19 are assessed. |
| BMI | Death-with-COVID-19 | Two versions: no effect, and OR=3. ^5^ |
| SARS-CoV-2 infection | Death-with-COVID-19 | Deterministic: Only participants with a SARS-CoV-2 infection can have a death-with-COVID-19 value. ^5^ |
| Confounders | Death-with-COVID-19 | Estimated in UK Biobank in assessed ^4^ subsample. Logistic regression of death-with-COVID-19 on confounders. ^5^ |

These tables detail how the parameters in the data generating mechanism of the simulations of BMI and (a) SARS-CoV-2 infection and (b) death-with-COVID-19. Each row corresponds to one or more edges in the respective DAG shown in Figure 1 in the main paper.

^1^ Intercept of logistic model set so that proportion infected was 3.16% in ALSPAC and 7.20% in UK Biobank.

^2^ Assessed subsample: Responded to COVID-19 questionnaire item in ALSPAC, or with a SARS-CoV-2 PCR test result or COVID-19 death in UK Biobank.

^3^ Intercept of logistic model used to generate ‘assessed’ set so that the proportion selected that seen in each cohort (19.97% responded to ALSPAC questionnaire and 1.156% tested in UK Biobank).

^4^ Assessed subsample in severity simulations: Having had a positive SARS-CoV-2 PCR test result or a death-with-COVID-19.

^5^ Intercept of logistic model generating death-with-COVID-19 set so to assume an infection fatality ratio (% COVID-19 deaths among all with a SARS-CoV-19 infection) of 3.13% [1].

#### Supplementary Table 5: Variable distributions of data generating mechanism used in simulations

| **Variable** | **ALSPAC variable distributions** | **UK Biobank variable distributions** |
| --- | --- | --- |
| Age | $\mathcal{\sim N}(0 1)$ | $\mathcal{\sim N}(0 1)$ |
| Sex | P_male_= 0.3226, P_female_= 0.6774 | P_male_= 0.4490, P_female_= 0.5510 |
| Education level | P_gcse_or_lower_= 0.1169, P_alevel_ = 0.2789, P_vocational_= 0.1394, P_degree_=0.4648 | P_alevel_ = 0.0541, P_vocational_=0.2803, P_degree_=0.3332, P_none_of_above_=0.3324 |
| BMI | $\mathcal{\sim N}(0 1)$ | $\mathcal{\sim N}(0 1)$ |
| Smoking | P_never_=0.3881, P_previous_= 0.3595, P_current_*=* 0.2524 | P_never_=0.5602, P_previous_=0.3409, P_current_*=*0.0989 |
| Deprivation | P_1_=0.3841, P_2_=0.2587, P_3_=0.1717, P_4_=0.1263, P_5_=0.0592 | $\mathcal{\sim N}(0 1)$ |
| SARS-CoV-2 infection | P_infection_=0.072, P_no_infection_=0.928 | P_infection_=0.0316, P_no_infection_=0.9684 |
| Assessed | P_assessed_=0.0984, P_not_assessed_=0.9016 * | P_assessed_=0.01156, P_not_assessed_=0.98844 |
| Death-with-COVID-19 | *NA* | P_death_=0.0316, P_no_death_=0.9684 |

#### * Probability of assessed in the ALSPAC simulations is lower than in the empirical data (P_assessed_=0.1974, P_not_assessed_=0.8026) because we are using a Poisson model and needed to ensure all probabilities generated using this model were between 0 and 1. Supplementary table 6: Results of simulations of SARS-CoV-2 infection based on ALSPAC G1 cohort, including unadjusted associations and bias-eliminated coverage

|  |  | **Bias (MCSE), coverage (MCSE) and bias-eliminated coverage (MCSE) of estimated effect of BMI on SARS-CoV-2 infection** | | | | |
| --- | --- | --- | --- | --- | --- | --- |
|  |  | SARS-CoV-2 (+) versus SARS-CoV-2 (-) | | | | SARS-CoV-2 (+) versus everyone else |
| ***Assumed interaction size of effect of BMI with SARS-CoV-2 infection on selection*** | Performance measure | All participants, unadjusted | All participants, confounder adjusted | Selected subsample, unadjusted | Selected subsample, confounder adjusted | All participants |
| **Assuming no effect of BMI on SARS-CoV-2 infection** | | | | | | |
| No interaction (log RR=0) | Bias (MCSE) | -0.0041 (0.0010) | -0.0017 (0.0010) | -0.0017 (0.0018) | -0.0002 (0.0019) | 0.0213 (0.0016) |
|  | Coverage (MCSE) | 0.945 (0.0072) | 0.945 (0.0072) | 0.940 (0.0075) | 0.944 (0.0073) | 0.922 (0.0085) |
|  | Bias-eliminated coverage (MCSE) | 0.943 (0.0073) | 0.942 (0.0074) | 0.940 (0.0075) | 0.943 (0.0073) | 0.949 (0.0070) |
| Plausible (log RR=0.0527) | Bias (MCSE) | -0.0022 (0.0010) | 0.0001 (0.0010) | 0.0501 (0.0018) | 0.0517 (0.0018) | 0.0595 (0.0015) |
|  | Coverage (MCSE) | 0.955 (0.0066) | 0.958 (0.0063) | 0.847 (0.0114) | 0.851 (0.0113) | 0.761 (0.0135) |
|  | Bias-eliminated coverage (MCSE) | 0.955 (0.0066) | 0.958 (0.0063) | 0.952 (0.0068) | 0.959 (0.0063) | 0.953 (0.0067) |
| Extreme (log RR=0.135) | Bias (MCSE) | -0.0022 (0.0010) | 0.0001 (0.0010) | 0.1323 (0.0018) | 0.1343 (0.0019) | 0.1194 (0.0015) |
|  | Coverage (MCSE) | 0.955 (0.0066) | 0.958 (0.0063) | 0.354 (0.0151) | 0.369 (0.0153) | 0.297 (0.0144) |
|  | Bias-eliminated coverage (MCSE) | 0.955 (0.0066) | 0.958 (0.0063) | 0.946 (0.0071) | 0.950 (0.0069) | 0.949 (0.0070) |
| **Assuming OR=3 for effect of BMI on SARS-CoV-2 infection** | | | | | | |
| No interaction (log RR=0) | Bias (MCSE) | -0.0092 (0.0012) | -0.0001 (0.0012) | -0.0042 (0.0023) | 0.0090 (0.0024) | -0.0802 (0.0017) |
|  | Coverage (MCSE) | 0.938 (0.0076) | 0.954 (0.0066) | 0.953 (0.0067) | 0.948 (0.0070) | 0.667 (0.0149) |
|  | Bias-eliminated coverage (MCSE) | 0.948 (0.0070) | 0.954 (0.0066) | 0.956 (0.0065) | 0.949 (0.0070) | 0.953 (0.0067) |
| Plausible (log RR=0.0527) | Bias (MCSE) | -0.0069 (0.0011) | 0.0018 (0.0012) | 0.0522 (0.0024) | 0.0652 (0.0024) | -0.0353 (0.0016) |
|  | Coverage (MCSE) | 0.948 (0.0070) | 0.955 (0.0066) | 0.910 (0.0090) | 0.886 (0.0101) | 0.914 (0.0089) |
|  | Bias-eliminated coverage (MCSE) | 0.953 (0.0067) | 0.954 (0.0066) | 0.948 (0.0070) | 0.949 (0.0070) | 0.958 (0.0063) |
| Extreme (log RR=0.135 | Bias (MCSE) | -0.0069 (0.0011) | 0.0018 (0.0012) | 0.1341 (0.0024) | 0.1479 (0.0025) | 0.0283 (0.0016) |
|  | Coverage (MCSE) | 0.948 (0.0070) | 0.955 (0.0066) | 0.604 (0.0155) | 0.543 (0.0158) | 0.933 (0.0079) |
|  | Bias-eliminated coverage (MCSE) | 0.953 (0.0067) | 0.954 (0.0066) | 0.952 (0.0068) | 0.953 (0.0067) | 0.957 (0.0064) |

#### Supplementary table 7: Results of simulations of SARS-CoV-2 infection and COVID-19 severity based on UK Biobank data, including unadjusted associations and bias-eliminated coverage

**a) Results of simulations estimating effect of BMI on SARS-CoV-2 infection**

|  |  | **Bias (MCSE), coverage (MCSE) and bias-eliminated coverage (MCSE) of estimated effect of BMI on SARS-CoV-2 infection** | | | | |
| --- | --- | --- | --- | --- | --- | --- |
|  |  | SARS-CoV-2 (+) versus SARS-CoV-2 (-) | | | | SARS-CoV-2 (+) versus everyone else |
| ***Interaction size of effect of BMI with SARS-CoV-2 infection on selection*** | **Performance measure** | All participants, unadjusted | All participants, confounder adjusted | Selected subsample, unadjusted | Selected subsample, confounder adjusted | All participants |
| **No effect of BMI on SARS-CoV-2 infection** | | | | | | |
| No interaction (log RR=0) | Bias (MCSE) | 0.0385 (0.0003) | 0.0001 (0.0003) | 0.0398 (0.0013) | 0.0020 (0.0013) | 0.1637 (0.0012) |
|  | Coverage (MCSE) | 0.007 (0.0026) | 0.956 (0.0065) | 0.825 (0.0120) | 0.961 (0.0061) | 0.012 (0.0034) |
|  | Bias-eliminated coverage (MCSE) | 0.953 (0.0067) | 0.956 (0.0065) | 0.956 (0.0065) | 0.963 (0.0060) | 0.956 (0.0065) |
| Plausible (log RR=-0.162) | Bias (MCSE) | 0.0386 (0.0003) | 0.0001 (0.0003) | -0.1226 (0.0013) | -0.1614 (0.0013) | 0.0259 (0.0012) |
|  | Coverage (MCSE) | 0.007 (0.0026) | 0.947 (0.0071) | 0.164 (0.0117) | 0.030 (0.0054) | 0.901 (0.0094) |
|  | Bias-eliminated coverage (MCSE) | 0.952 (0.0068) | 0.948 (0.0070) | 0.951 (0.0068) | 0.955 (0.0066) | 0.957 (0.0064) |
| Extreme (log RR=-0.245) | Bias (MCSE) | 0.0386 (0.0003) | 0.0001 (0.0003) | -0.2050 (0.0013) | -0.2440 (0.0014) | -0.0386 (0.0012) |
|  | Coverage (MCSE) | 0.007 (0.0026) | 0.947 (0.0071) | 0.000 (0.0000) | 0.000 (0.0000) | 0.834 (0.0118) |
|  | Bias-eliminated coverage (MCSE) | 0.952 (0.0068) | 0.948 (0.0070) | 0.946 (0.0071) | 0.946 (0.0071) | 0.951 (0.0068) |
| **OR=3 for effect of BMI on SARS-CoV-2 infection** | | | | | | |
| No interaction (log RR=0) | Bias (MCSE) | 0.0333 (0.0003) | 0.0002 (0.0003) | 0.0356 (0.0015) | 0.0046 (0.0015) | 0.0604 (0.0011) |
|  | Coverage (MCSE) | 0.075 (0.0083) | 0.938 (0.0076) | 0.896 (0.0097) | 0.950 (0.0069) | 0.608 (0.0154) |
|  | Bias-eliminated coverage (MCSE) | 0.941 (0.0075) | 0.937 (0.0077) | 0.946 (0.0071) | 0.948 (0.0070) | 0.955 (0.0066) |
| Plausible (log RR=-0.162) | Bias (MCSE) | 0.0333 (0.0003) | 0.0001 (0.0003) | -0.1283 (0.0015) | -0.1599 (0.0015) | -0.0702 (0.0011) |
|  | Coverage (MCSE) | 0.076 (0.0084) | 0.946 (0.0071) | 0.211 (0.0129) | 0.072 (0.0082) | 0.493 (0.0158) |
|  | Bias-eliminated coverage (MCSE) | 0.945 (0.0072) | 0.946 (0.0071) | 0.944 (0.0073) | 0.951 (0.0068) | 0.958 (0.0063) |
| Extreme (log RR=-0.245) | Bias (MCSE) | 0.0333 (0.0003) | 0.0001 (0.0003) | -0.2108 (0.0014) | -0.2427 (0.0015) | -0.1305 (0.0011) |
|  | Coverage (MCSE) | 0.076 (0.0084) | 0.946 (0.0071) | 0.004 (0.0020) | 0.002 (0.0014) | 0.046 (0.0066) |
|  | Bias-eliminated coverage (MCSE) | 0.945 (0.0072) | 0.946 (0.0071) | 0.944 (0.0073) | 0.942 (0.0074) | 0.949 (0.0070) |

**b) Results of simulations estimating effect of BMI on death-with-COVID-19**

|  |  | **Bias (MCSE), coverage (MCSE) and bias-eliminated coverage (MCSE) of estimated effect of BMI on death-with-COVID-19** | | | | |
| --- | --- | --- | --- | --- | --- | --- |
| ***Interaction size of effect of BMI with SARS-CoV-2 infection on selection*** |  | Death-with-COVID-19 versus SARS-CoV-2 (+) not resulting in death-with-COVID-19 | | | | Death-with- COVID-19 versus controls=everyone (i.e. whole sample) |
|  | **Performance measure** | SARS-CoV-2 (+) subsample, unadjusted | SARS-CoV-2 (+) subsample, confounder adjusted | SARS-CoV-2 (+) and assessed subsample, unadjusted | SARS-CoV-2 (+) and assessed subsample, confounder adjusted | All participants |
| **No effect of BMI on death-with-COVID-19** | | | | | | |
| No interaction (log RR=0) | Bias (MCSE) | 0.0296 (0.0016) | -0.0013 (0.0017) | -0.1567 (0.0020) | -0.1652 (0.0022) | -0.0018 (0.0016) |
|  | Coverage (MCSE) | 0.900 (0.0095) | 0.951 (0.0068) | 0.279 (0.0142) | 0.326 (0.0148) | 0.945 (0.0072) |
|  | Bias-eliminated coverage (MCSE) | 0.943 (0.0073) | 0.949 (0.0070) | 0.948 (0.0070) | 0.941 (0.0075) | 0.944 (0.0073) |
| Plausible (log RR=-0.162) | Bias (MCSE) | 0.0305 (0.0015) | -0.0004 (0.0016) | -0.0193 (0.0019) | -0.0233 (0.0022) | -0.0004 (0.0015) |
|  | Coverage (MCSE) | 0.917 (0.0087) | 0.952 (0.0068) | 0.946 (0.0071) | 0.935 (0.0078) | 0.957 (0.0064) |
|  | Bias-eliminated coverage (MCSE) | 0.957 (0.0064) | 0.954 (0.0066) | 0.953 (0.0067) | 0.950 (0.0069) | 0.957 (0.0064) |
| Extreme (log RR=-0.245) | Bias (MCSE) | 0.0305 (0.0015) | -0.0004 (0.0016) | 0.0458 (0.0019) | 0.0423 (0.0022) | -0.0004 (0.0015) |
|  | Coverage (MCSE) | 0.917 (0.0087) | 0.952 (0.0068) | 0.904 (0.0093) | 0.909 (0.0091) | 0.957 (0.0064) |
|  | Bias-eliminated coverage (MCSE) | 0.957 (0.0064) | 0.954 (0.0066) | 0.958 (0.0063) | 0.953 (0.0067) | 0.957 (0.0064) |
| **OR=3 for effect of BMI on death-with-COVID-19** | | | | | | |
| No interaction (log RR=0) | Bias (MCSE) | -0.0340 (0.0017) | 0.0012 (0.0019) | -0.2027 (0.0024) | -0.1525 (0.0027) | -0.1404 (0.0016) |
|  | Coverage (MCSE) | 0.896 (0.0097) | 0.933 (0.0079) | 0.237 (0.0134) | 0.535 (0.0158) | 0.204 (0.0127) |
|  | Bias-eliminated coverage (MCSE) | 0.935 (0.0078) | 0.936 (0.0077) | 0.945 (0.0072) | 0.937 (0.0077) | 0.942 (0.0074) |
| Plausible (log RR=-0.162) | Bias (MCSE) | -0.0334 (0.0016) | 0.0016 (0.0018) | -0.0636 (0.0025) | -0.0100 (0.0028) | -0.1381 (0.0015) |
|  | Coverage (MCSE) | 0.913 (0.0089) | 0.953 (0.0067) | 0.864 (0.0108) | 0.952 (0.0068) | 0.190 (0.0124) |
|  | Bias-eliminated coverage (MCSE) | 0.965 (0.0058) | 0.953 (0.0067) | 0.949 (0.0070) | 0.955 (0.0066) | 0.961 (0.0061) |
| Extreme (log RR=-0.245) | Bias (MCSE) | -0.0334 (0.0016) | 0.0016 (0.0018) | 0.0029 (0.0025) | 0.0564 (0.0028) | -0.1381 (0.0015) |
|  | Coverage (MCSE) | 0.913 (0.0089) | 0.953 (0.0067) | 0.950 (0.0069) | 0.916 (0.0088) | 0.190 (0.0124) |
|  | Bias-eliminated coverage (MCSE) | 0.965 (0.0058) | 0.953 (0.0067) | 0.952 (0.0068) | 0.956 (0.0065) | 0.961 (0.0061) |

MCSE: Monte Carlo standard error (across 1000 repetitions). Simulations based on UK Biobank PCR test results from national testing pre-mass testing.

Bias given is the difference in the estimated versus true effect of BMI on (a) SARS-CoV-2 infection, and (b) death-with-COVID-19. Coverage is the proportion of simulation repetitions with confidence intervals containing the true effect.

Sample sizes: a) All N = 421,027; selected subsample N~18,000, and b) SARS-CoV-2 (+) subsample N~13,300K; SARS-CoV-2 (+) and assessed subsample N~1,600; whole sample N=421,037.

Example biases: A bias of -0.1618 ^$^ when no effect of BMI on SARS-CoV-2 infection (plausible scenario) is equivalent to an estimated odds ratio of 0.85 per 1SD higher BMI (compared with true odds ratio = 1). A bias of 0.0735 ^#^ when BMI effect on death-with-COVID-19 is OR=3 is equivalent to an estimated odds ratio of 3.23 (compared with true odds ratio =3).

Histograms of simulation results are shown in Supplementary figure 11.

#### Supplementary table 8: Results of simulations of SARS-CoV-2 infection based on ALSPAC G1 cohort using larger sample size of 5 million

|  |  | **Bias (MCSE) or coverage (MCSE) of estimated effect of BMI on SARS-CoV-2 infection** | | | | |
| --- | --- | --- | --- | --- | --- | --- |
|  |  | SARS-CoV-2 (+) versus SARS-CoV-2 (-) | | | | SARS-CoV-2 (+) versus everyone else |
| ***Interaction size of effect of BMI with SARS-CoV-2 infection on selection*** | Performance measure | All participants, unadjusted | All participants, confounder adjusted | Selected subsample, unadjusted | Selected subsample, confounder adjusted | All participants |
| **No effect of BMI on SARS-CoV-2 infection** | | | | | | |
| No interaction (log RR=0) | Bias (MCSE) | -0.0027 (0.0001) | -0.0000 (0.0001) | -0.0019 (0.0001) | -0.0002 (0.0001) | 0.0214 (0.0001) |
|  | Coverage (MCSE) | 0.672 (0.0148) | 0.958 (0.0063) | 0.907 (0.0092) | 0.954 (0.0066) | 0.000 (0.0000) |
| Plausible (log RR=0.0527) | Bias (MCSE) | -0.0025 (0.0001) | 0.0001 (0.0001) | 0.0511 (0.0001) | 0.0528 (0.0001) | 0.0597 (0.0001) |
|  | Coverage (MCSE) | 0.683 (0.0147) | 0.959 (0.0063) | 0.000 (0.0000) | 0.000 (0.0000) | 0.000 (0.0000) |
| Extreme (log RR=0.135) | Bias (MCSE) | -0.0025 (0.0001) | 0.0001 (0.0001) | 0.1334 (0.0001) | 0.1351 (0.0001) | 0.1200 (0.0001) |
|  | Coverage (MCSE) | 0.683 (0.0147) | 0.959 (0.0063) | 0.000 (0.0000) | 0.000 (0.0000) | 0.000 (0.0000) |
| **Assuming, OR=3 for effect of BMI on SARS-CoV-2 infection** | | | | | | |
| No interaction (log RR=0) | Bias (MCSE) | -0.0081 (0.0001) | -0.0000 (0.0001) | -0.0061 (0.0001) | 0.0000 (0.0001) | -0.0795 (0.0001) |
|  | Coverage (MCSE) | 0.027 (0.0051) | 0.949 (0.0070) | 0.668 (0.0149) | 0.956 (0.0065) | 0.000 (0.0000) |
| Plausible (log RR=0.0527) | Bias (MCSE) | -0.0081 (0.0001) | -0.0000 (0.0001) | 0.0466 (0.0001) | 0.0527 (0.0001) | -0.0403 (0.0001) |
|  | Coverage (MCSE) | 0.019 (0.0043) | 0.958 (0.0063) | 0.000 (0.0000) | 0.000 (0.0000) | 0.000 (0.0000) |
| Extreme (log RR=0.135 | Bias (MCSE) | -0.0081 (0.0001) | -0.0000 (0.0001) | 0.1288 (0.0001) | 0.1350 (0.0001) | 0.0231 (0.0001) |
|  | Coverage (MCSE) | 0.019 (0.0043) | 0.958 (0.0063) | 0.000 (0.0000) | 0.000 (0.0000) | 0.000 (0.0000) |

#### Supplementary table 9: Results of simulations of SARS-CoV-2 infection and COVID-19 severity based on UK Biobank data using larger sample size of 5 million

**a) Results of simulations estimating effect of BMI on SARS-CoV-2 infection**

|  |  | **Bias (MCSE) or coverage (MCSE) of estimated effect of BMI on SARS-CoV-2 infection** | | | | |
| --- | --- | --- | --- | --- | --- | --- |
|  |  | SARS-CoV-2 (+) versus SARS-CoV-2 (-) | | | | SARS-CoV-2 (+) versus everyone else |
| ***Interaction size of effect of BMI with SARS-CoV-2 infection on selection*** | **Performance measure** | All participants, unadjusted | All participants, confounder adjusted | Selected subsample, unadjusted | Selected subsample, confounder adjusted | All participants |
| **No effect of BMI on SARS-CoV-2 infection** | | | | | | |
| No interaction (log RR=0) | Bias (MCSE) | 0.0384 (0.0001) | 0.0000 (0.0001) | 0.0387 (0.0004) | 0.0006 (0.0004) | 0.1623 (0.0004) |
|  | Coverage (MCSE) | 0.000 (0.0000) | 0.956 (0.0065) | 0.082 (0.0087) | 0.950 (0.0069) | 0.000 (0.0000) |
| Plausible (log RR=-0.162) | Bias (MCSE) | 0.0384 (0.0001) | 0.0000 (0.0001) | -0.1231 (0.0004) | -0.1614 (0.0004) | 0.0259 (0.0004) |
|  | Coverage (MCSE) | 0.000 (0.0000) | 0.962 (0.0060) | 0.000 (0.0000) | 0.000 (0.0000) | 0.366 (0.0152) |
| Extreme (log RR=-0.245) | Bias (MCSE) | 0.0384 (0.0001) | 0.0000 (0.0001) | -0.2061 (0.0004) | -0.2444 (0.0004) | -0.0393 (0.0004) |
|  | Coverage (MCSE) | 0.000 (0.0000) | 0.962 (0.0060) | 0.000 (0.0000) | 0.000 (0.0000) | 0.065 (0.0078) |
| **Assuming, OR=3 for effect of BMI on SARS-CoV-2 infection** | | | | | | |
| No interaction (log RR=0) | Bias (MCSE) | 0.0332 (0.0001) | 0.0001 (0.0001) | 0.0333 (0.0004) | 0.0004 (0.0004) | 0.0588 (0.0003) |
|  | Coverage (MCSE) | 0.000 (0.0000) | 0.971 (0.0053) | 0.313 (0.0147) | 0.951 (0.0068) | 0.000 (0.0000) |
| Plausible (log RR=-0.162) | Bias (MCSE) | 0.0331 (0.0001) | -0.0000 (0.0001) | -0.1287 (0.0004) | -0.1619 (0.0004) | -0.0697 (0.0003) |
|  | Coverage (MCSE) | 0.000 (0.0000) | 0.963 (0.0060) | 0.000 (0.0000) | 0.000 (0.0000) | 0.000 (0.0000) |
| Extreme (log RR=-0.245) | Bias (MCSE) | 0.0331 (0.0001) | -0.0000 (0.0001) | -0.2116 (0.0004) | -0.2449 (0.0004) | -0.1305 (0.0003) |
|  | Coverage (MCSE) | 0.000 (0.0000) | 0.963 (0.0060) | 0.000 (0.0000) | 0.000 (0.0000) | 0.000 (0.0000) |

**b) Results of simulations estimating effect of BMI on COVID-19 severity**

|  |  | **Bias (MCSE) or coverage (MCSE) of estimated effect of BMI on COVID-19 severity** | | | | |
| --- | --- | --- | --- | --- | --- | --- |
| ***Interaction size of effect of BMI with SARS-CoV-2 infection on selection*** |  | Death-with-COVID-19 versus SARS-CoV-2 (+) not resulting in death-with-COVID-19 | | | | Death-with-COVID-19 versus controls=everyone (i.e. whole sample) |
|  | **Performance measure** | SARS-CoV-2 (+) subsample, unadjusted | SARS-CoV-2 (+) subsample, confounder adjusted | SARS-CoV-2 (+) and assessed subsample, unadjusted | SARS-CoV-2 (+) and assessed subsample, confounder adjusted | All participants |
| **No effect of BMI on COVID-19 severity** | | | | | | |
| No interaction (log RR=0) | Bias (MCSE) | 0.0302 (0.0005) | -0.0005 (0.0005) | -0.1565 (0.0006) | -0.1626 (0.0006) | -0.0005 (0.0005) |
|  | Coverage (MCSE) | 0.466 (0.0158) | 0.956 (0.0065) | 0.000 (0.0000) | 0.000 (0.0000) | 0.950 (0.0069) |
| Plausible (log RR=-0.162) | Bias (MCSE) | 0.0308 (0.0005) | -0.0000 (0.0005) | -0.0187 (0.0006) | -0.0243 (0.0007) | -0.0002 (0.0004) |
|  | Coverage (MCSE) | 0.422 (0.0156) | 0.966 (0.0057) | 0.818 (0.0122) | 0.770 (0.0133) | 0.960 (0.0062) |
| Extreme (log RR=-0.245) | Bias (MCSE) | 0.0308 (0.0005) | -0.0000 (0.0005) | 0.0464 (0.0006) | 0.0409 (0.0007) | -0.0002 (0.0004) |
|  | Coverage (MCSE) | 0.422 (0.0156) | 0.966 (0.0057) | 0.295 (0.0144) | 0.481 (0.0158) | 0.960 (0.0062) |
| **Assuming, OR=3 for effect of BMI on COVID-19 severity** | | | | | | |
| No interaction (log RR=0) | Bias (MCSE) | -0.0319 (0.0005) | 0.0008 (0.0005) | -0.2035 (0.0007) | -0.1604 (0.0008) | -0.1379 (0.0004) |
|  | Coverage (MCSE) | 0.470 (0.0158) | 0.948 (0.0070) | 0.000 (0.0000) | 0.000 (0.0000) | 0.000 (0.0000) |
| Plausible (log RR=-0.162) | Bias (MCSE) | -0.0340 (0.0005) | -0.0014 (0.0005) | -0.0650 (0.0007) | -0.0237 (0.0008) | -0.1397 (0.0005) |
|  | Coverage (MCSE) | 0.435 (0.0157) | 0.932 (0.0080) | 0.199 (0.0126) | 0.826 (0.0120) | 0.000 (0.0000) |
| Extreme (log RR=-0.245) | Bias (MCSE) | -0.0340 (0.0005) | -0.0014 (0.0005) | 0.0012 (0.0008) | 0.0416 (0.0008) | -0.1397 (0.0005) |
|  | Coverage (MCSE) | 0.435 (0.0157) | 0.932 (0.0080) | 0.950 (0.0069) | 0.628 (0.0153) | 0.000 (0.0000) |

### Supplementary figures

#### Supplementary Figure 1: Flow chart for eligible participants in the studies in ALSPAC and UK Biobank

**a) ALSPAC study flow chart for eligible participants in the G1 cohort**

G1 cohort

N = 14,849

Withdrawn from study or declined questionnaires: N = 4,842

No email address: N = 3,870

**G1 eligible for Q1 analyses**

**having received Q1 questionnaire**

Responded to COVID-19 question

(i.e. assessed): N = 2,966

Reported having a SARS-CoV-2 infection: N = 472 (52 positive test or suspected by doctor, 420 own-suspected)

**G1 eligible for Q2 analyses**

**having received Q2 questionnaire**

Responded to COVID-19 question

(i.e. assessed): N = 2,704

Reported having a SARS-CoV-2 infection: N = 478 (76 positive test or suspected by doctor, 402 own-suspected)

Withdrawn from study or declined questionnaires: N = 4,832

No email address: N = 4,173

**b) ALSPAC study flow chart for eligible participants in the G0 mothers cohort**

G0 mothers cohort

N = 14,282

Withdrawn from study or declined questionnaires: N = 3,941

No email address: N = 5,697

**G0 mothers eligible for Q1 analyses having received Q1 questionnaire**

Responded to COVID-19 question

(i.e. assessed): N = 2,685

Reported having a SARS-CoV-2 infection: N = 347 (25 positive test or suspected by doctor, 322 own-suspected)

**G0 mothers eligible Q2 analyses**

**having received Q2 questionnaire**

Responded to COVID-19 question

(i.e. assessed): N = 2,632

Reported having a SARS-CoV-2 infection: N = 332 (35 positive test or suspected by doctor, 297 own-suspected)

Withdrawn from study or declined questionnaires: N = 3,937

No email address: N = 5,745

**c) UK Biobank study flow chart for eligible participant**

Recruited at baseline

N = 503,317

In study sample

N = 502,117

Withdrawn N = 829

Pregnant at baseline N = 371

Alive in 2020

N = 473,580

Residing in England at baseline

N = 421,037

**Eligible for analyses pre-mass testing N = 421,037**

Covid Test N = 4,804

SARS-CoV-2 test or COVID-19 death = 4,869

SARS-CoV-2 (+) confirmed via test N = 1,352

SARS-CoV-2 (+) and/or death-with-COVID-19 = 1,440

Death-with-COVID-19 and SARS-CoV-2 (+) N = 254

Death-with-COVID-19 N = 342

Died before 01/01/2020 N = 28,537

Attended baseline assessment centre in Wales or Scotland

N = 52,543

Died between 01/01/2020 and 18/05/2020 N = 1,811

**Eligible for analyses post-mass testing N = 419,226**

Covid Test N = 56,589

SARS-CoV-2 test or COVID-19 death = 56,616

SARS-CoV-2 (+) confirmed via test N = 12,940

SARS-CoV-2 (+) and/or death-with-COVID-19 = 12,974

Death-with-COVID-19 and SARS-CoV-2 (+) N = 214

Death-with-COVID-19 N = 248

#### Supplementary Figure 2: Forest plots of the association between the candidate predictors of selection and outcomes related to SARS-CoV-2 infection in the ALSPAC G1 cohort – questionnaires 1 and 2

**a) Categorical predictors of selection**

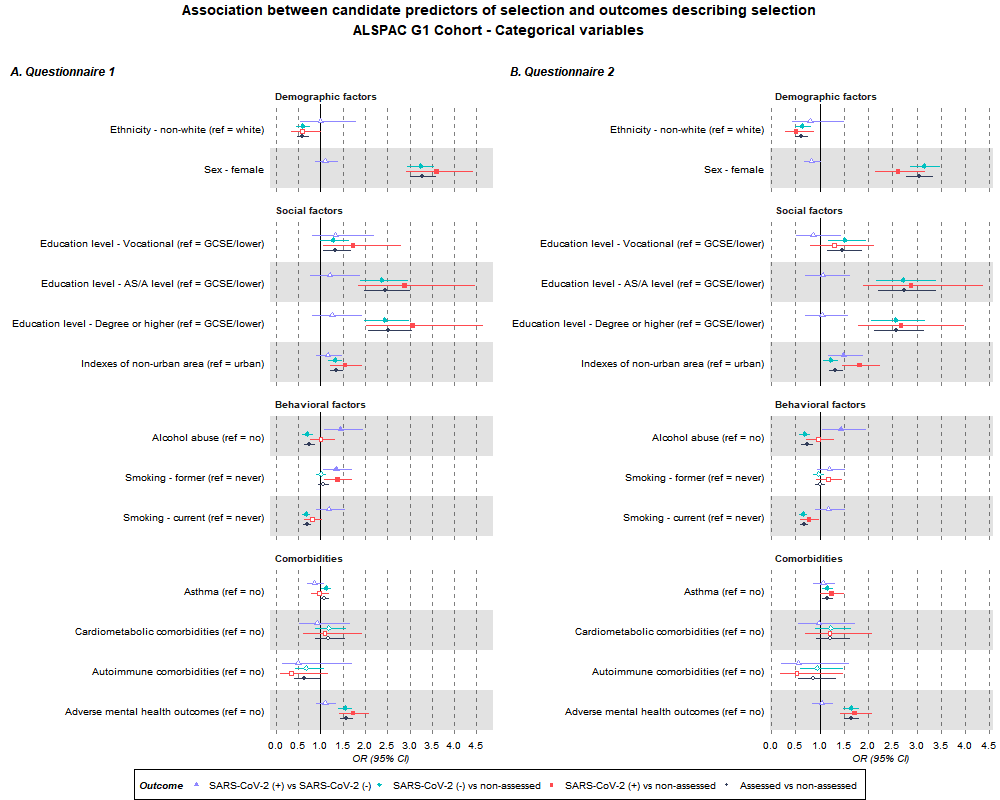

**b) Continuous predictors of selection**

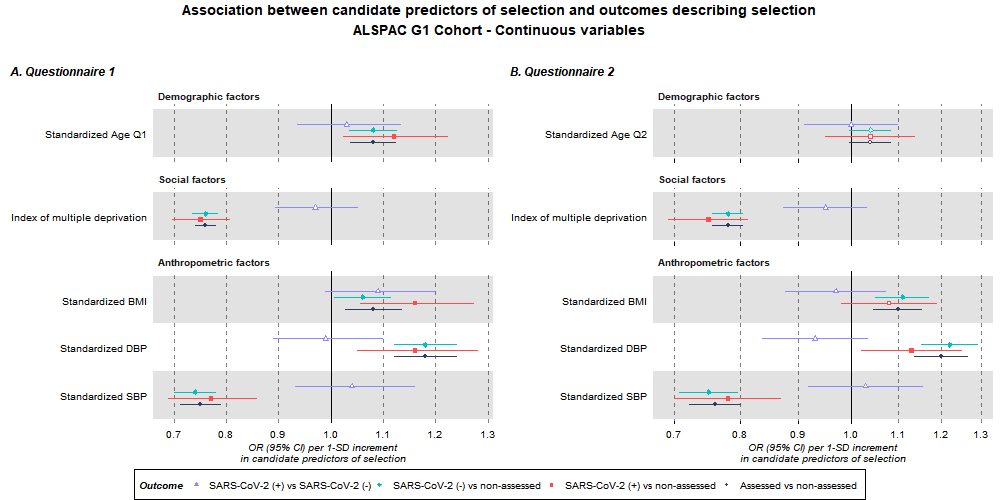

ORs and their 95% confidence intervals are shown for categorical variables (a) and continuous variables (b). Estimates for continuous candidate predictors are per 1 standard deviation for each predictor except for IMD which is given per 1 higher quantile.

#### Supplementary Figure 3: Forest plots of the association between the candidate predictors of selection and outcomes related to SARS-CoV-2 infection in the ALSPAC G0 Mothers cohort

**a. Categorical predictors of selection**

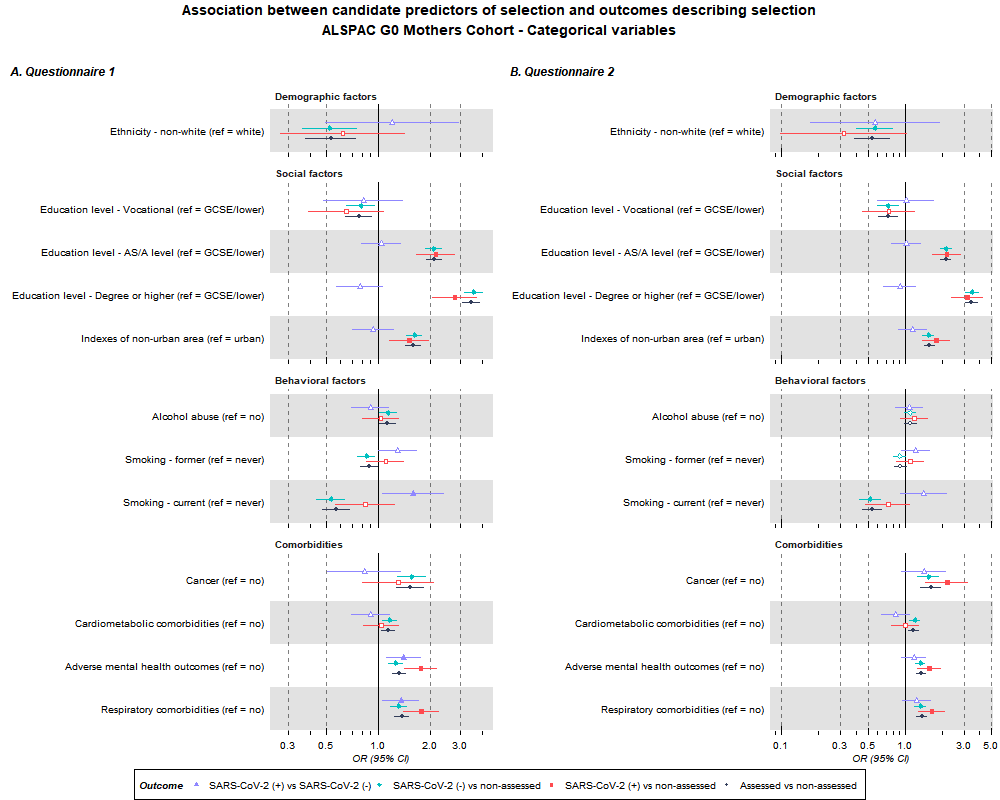

**b) Continuous predictors of selection**

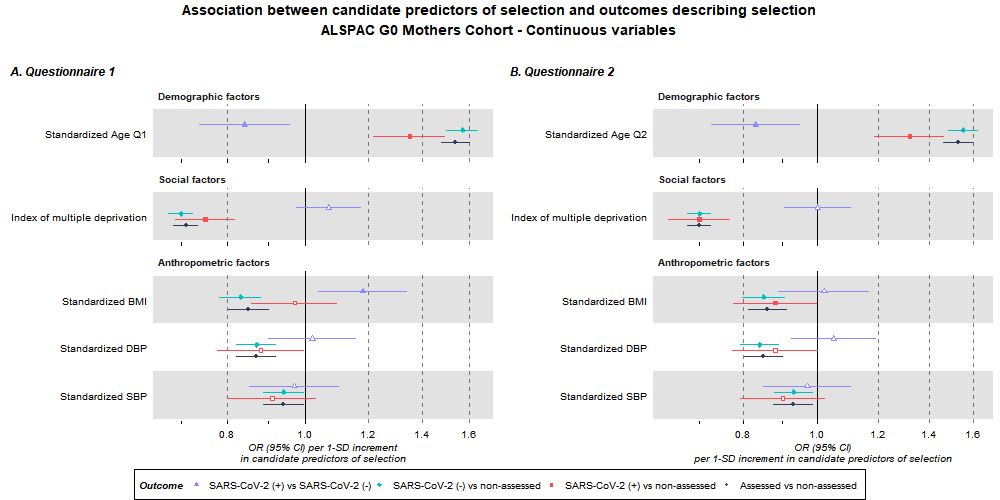

ORs and their 95% confidence intervals are shown for categorical variables (a) and continuous variables (b). Estimates for continuous candidate predictors are per 1 standard deviation for each predictor except for IMD which is given per 1 higher quantile.

#### Supplementary Figure 4: Forest plots of the association between the candidate predictors of selection and outcomes related to SARS-CoV-2 infection and COVID-19 severity in UK Biobank – pre- and post-mass testing

a) SARS-CoV-2 infection, pre mass testing, categorical variables

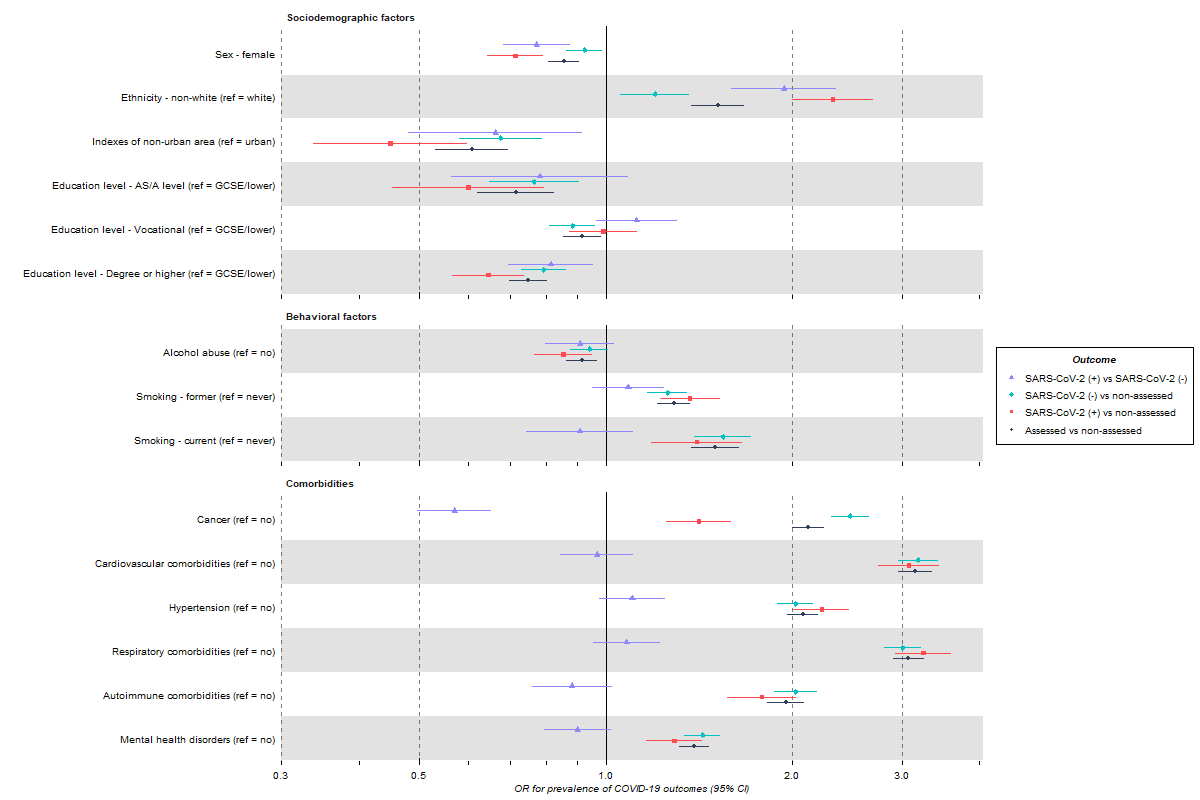

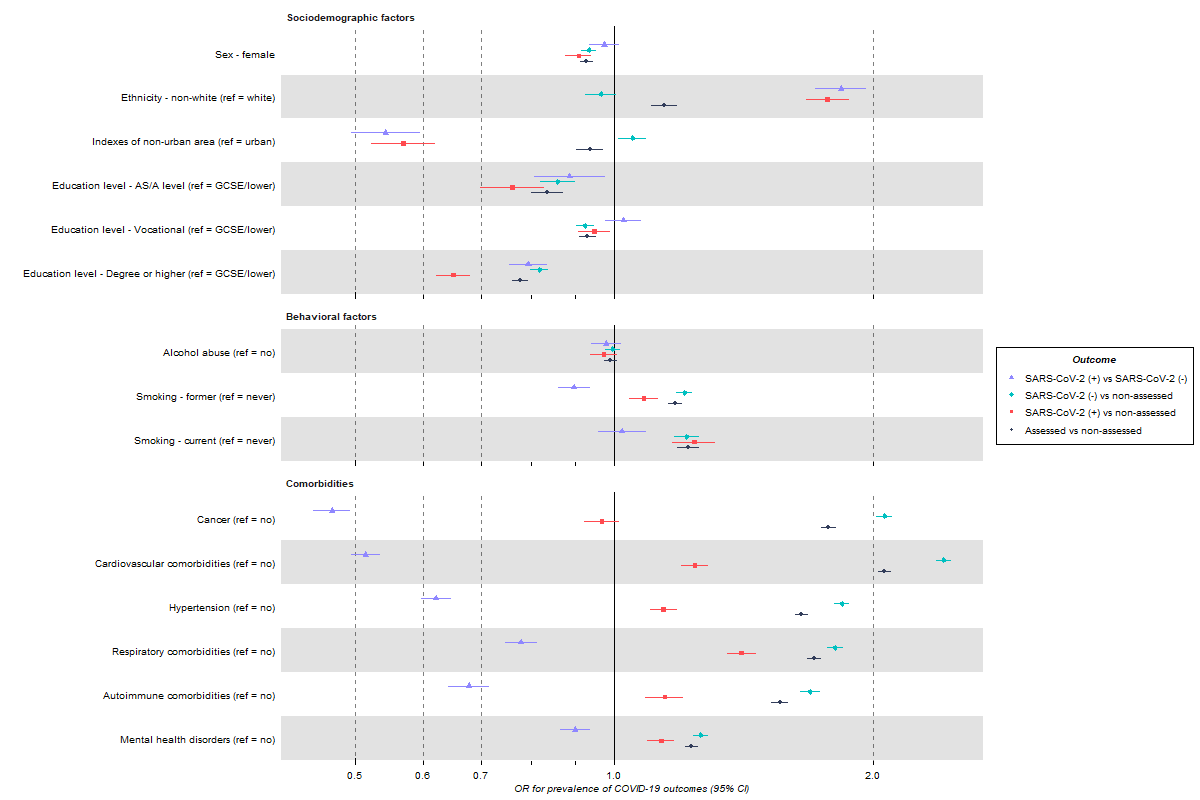

b) SARS-CoV-2 infection, post mass testing, categorical variables

,
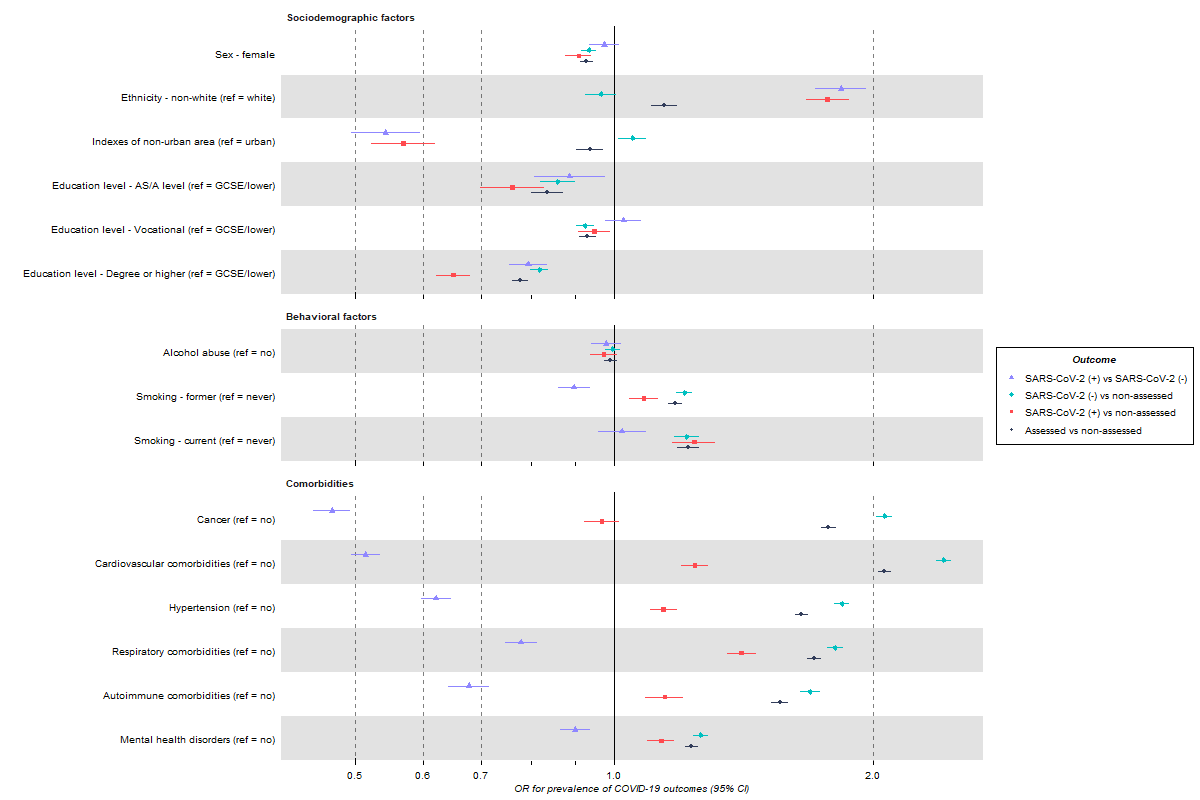

c) SARS-CoV-2 infection, pre mass testing, continuous variables

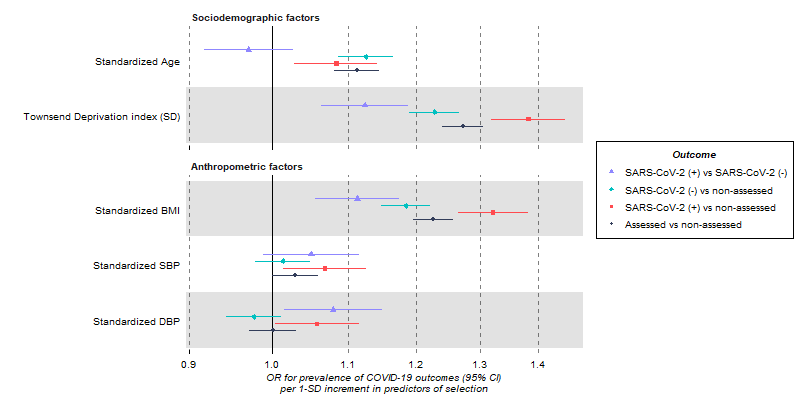

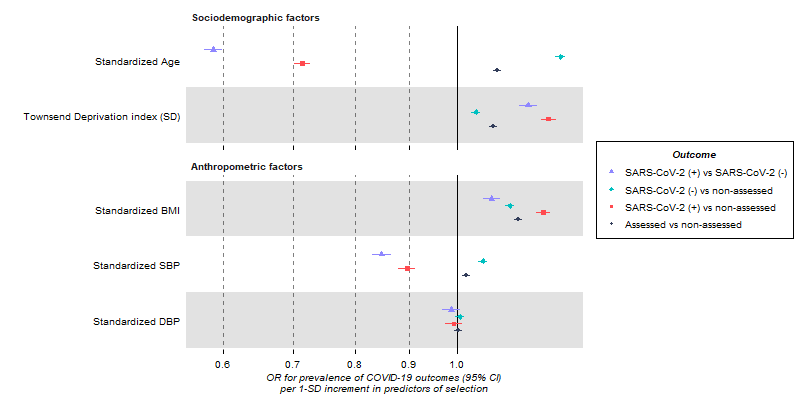

d) SARS-CoV-2 infection, post mass testing, continuous variables

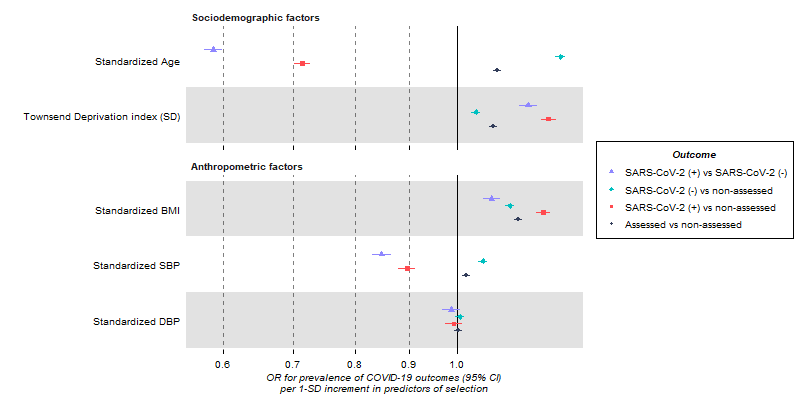

e) COVID-19 severity, pre mass testing, categorical variables

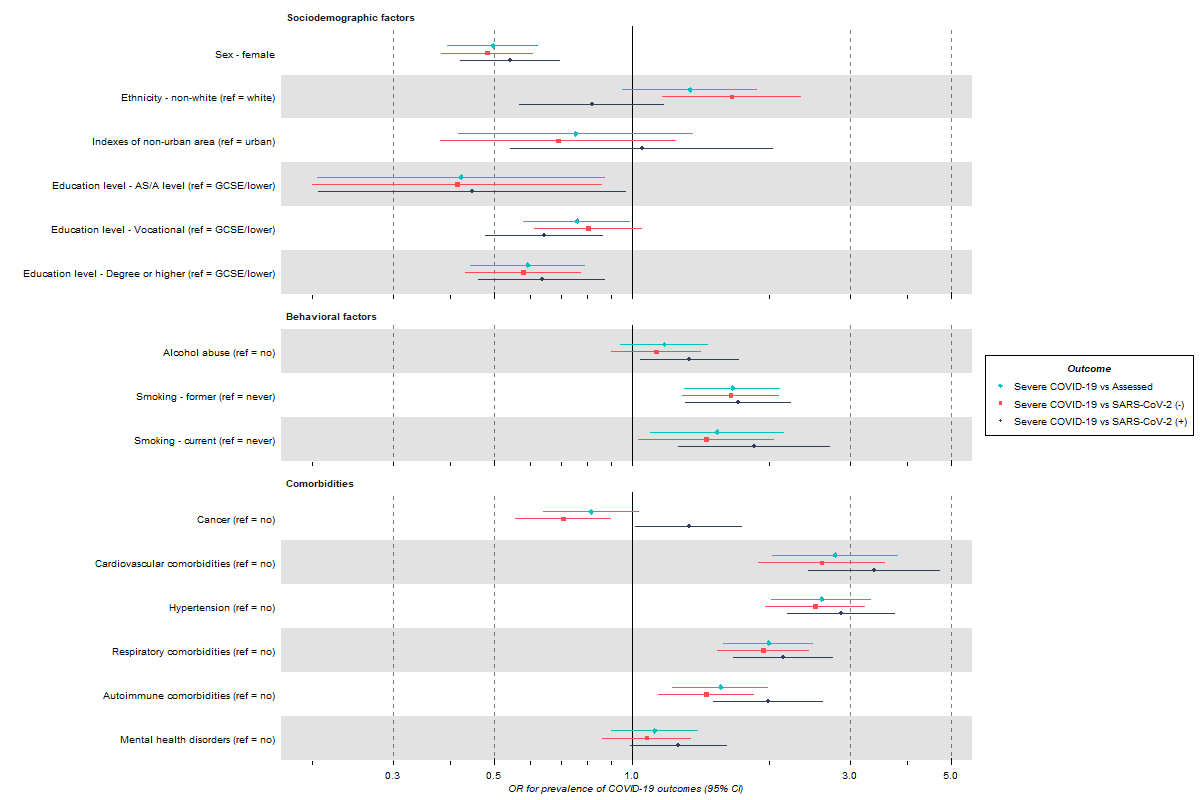

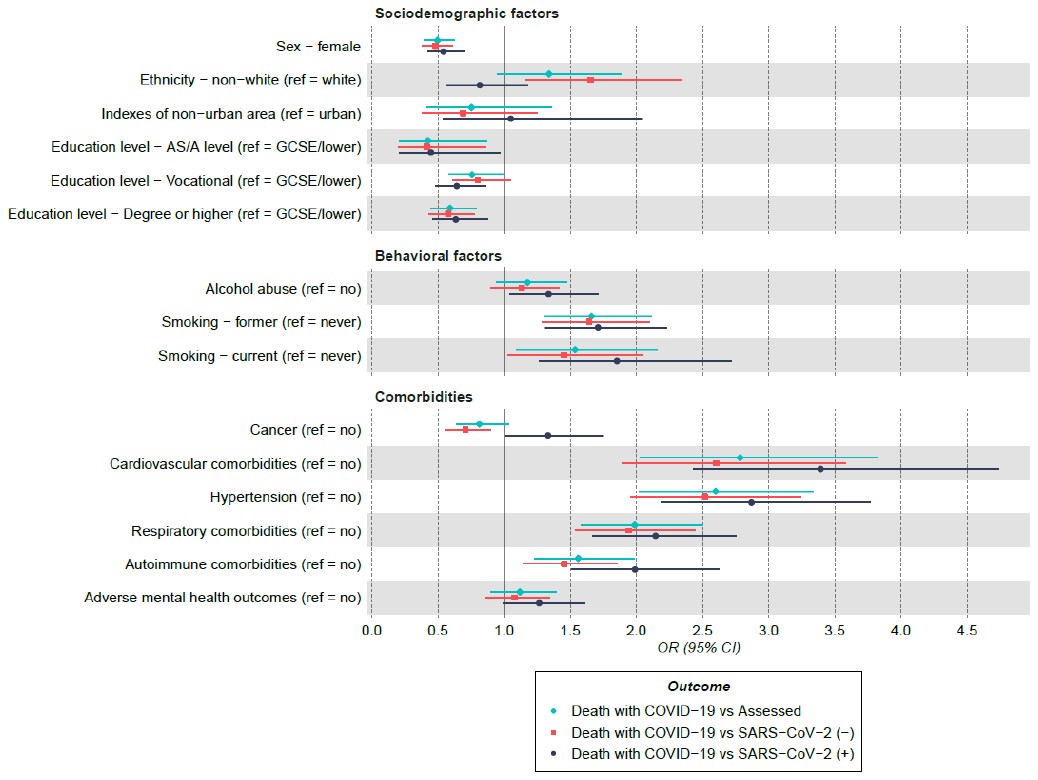

f) COVID-19 severity, post mass testing, categorical variables

,
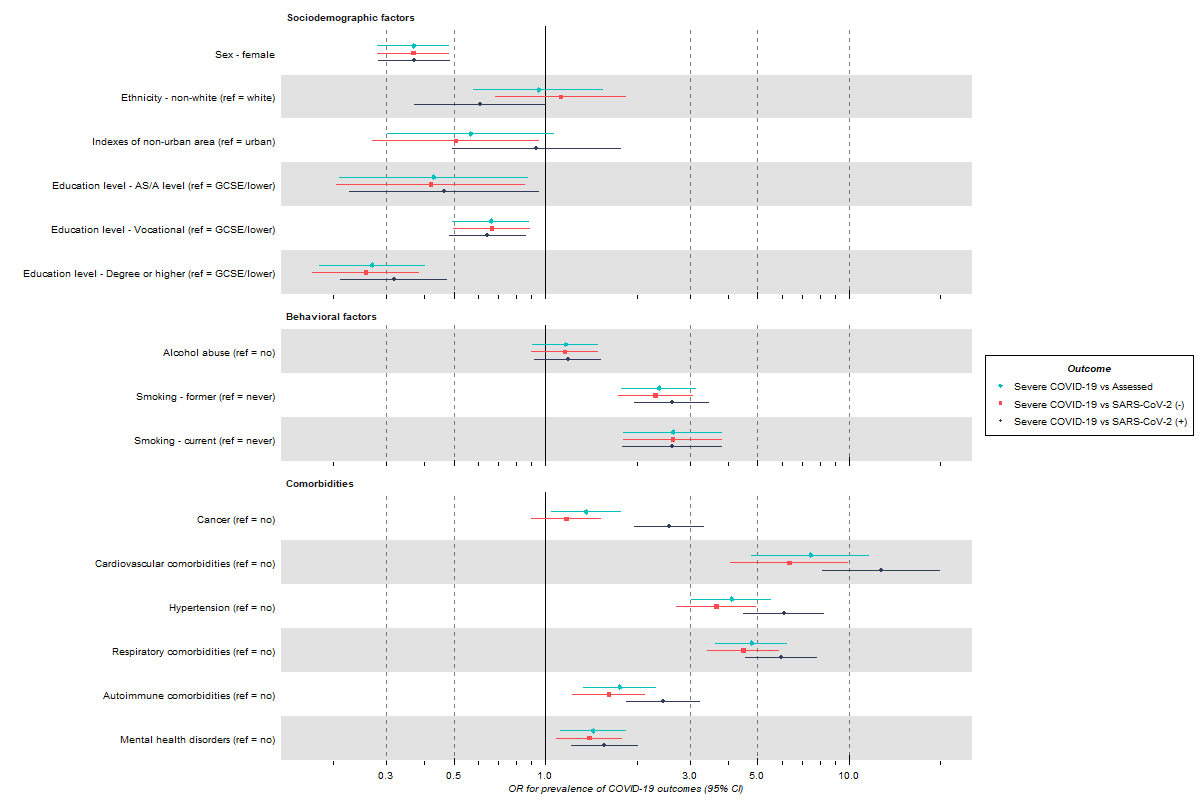

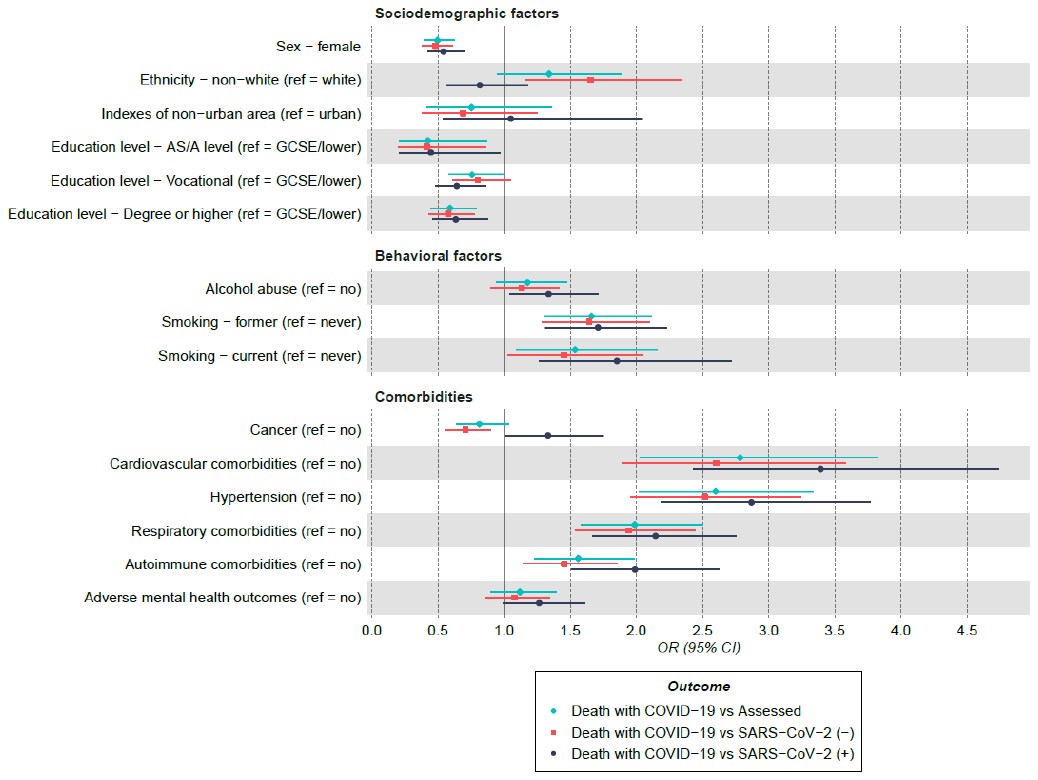

g) COVID-19 severity, pre mass testing, continuous variables

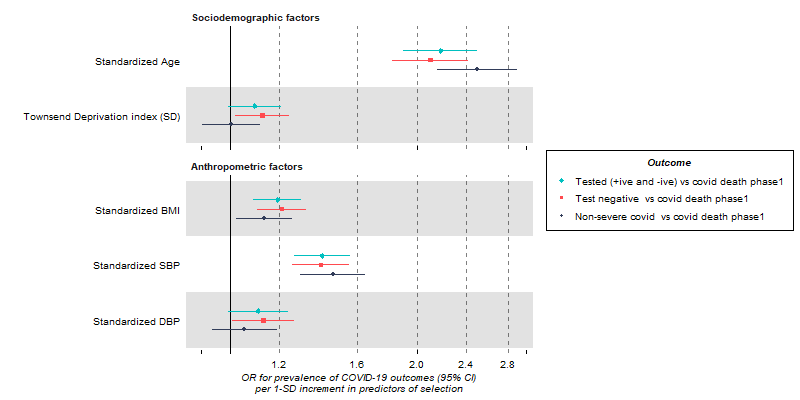

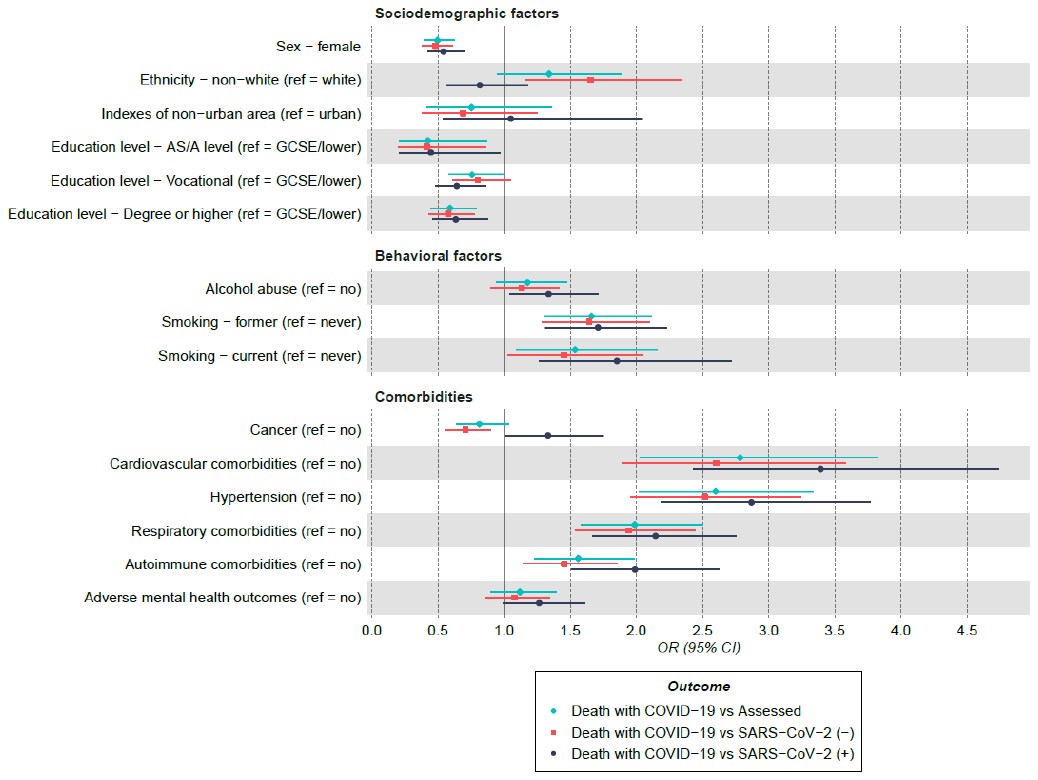

h) COVID-19 severity, post mass testing, continuous variables

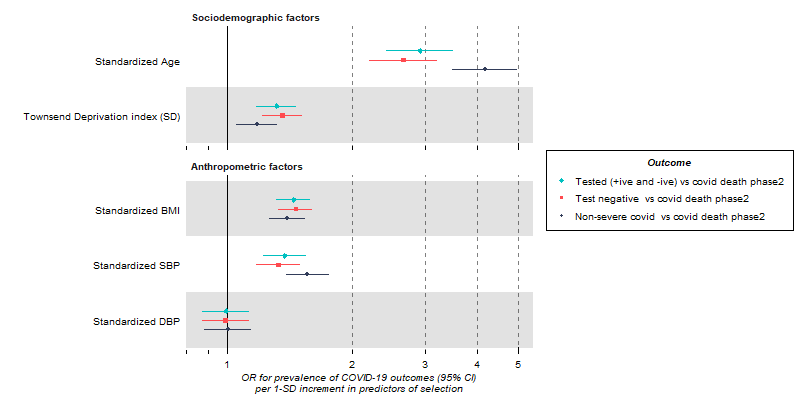

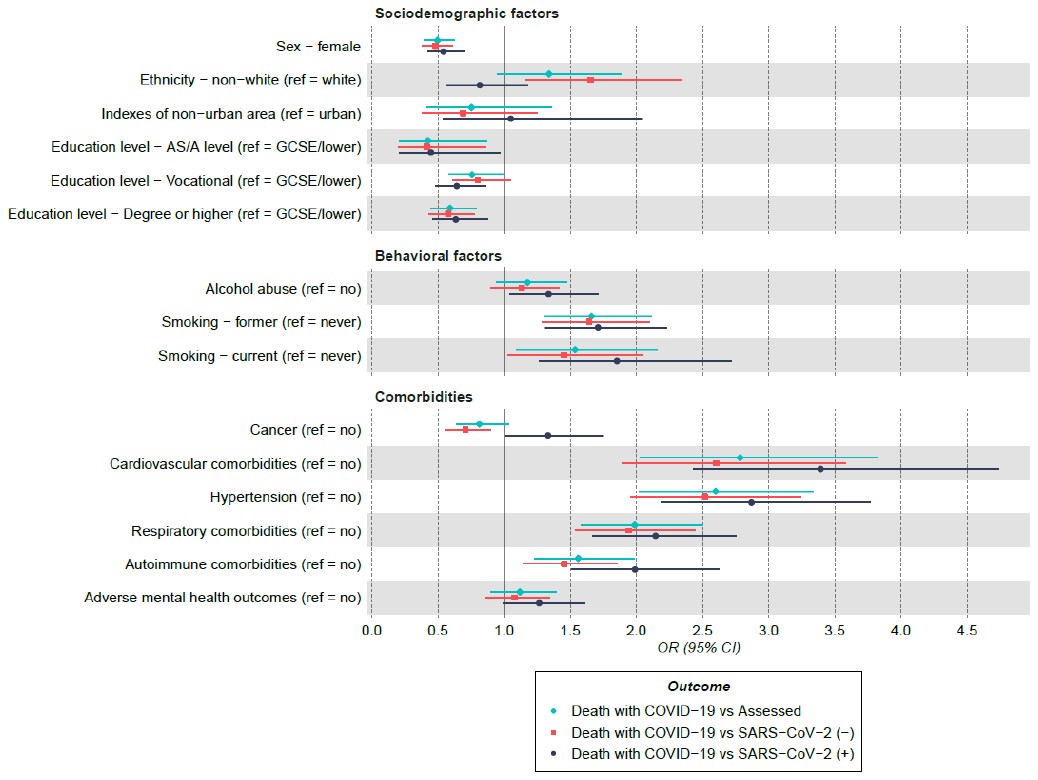

#### Supplementary Figure 5: Forest plots of the association between BMI and SARS-CoV-2 infection in the ALSPAC G1 cohort

Odds ratio and 95% confidence intervals using data from questionnaire 1 (panel A) and questionnaire 2 (panel B), for the whole cohort and the subsamples stratified by sex. Models were adjusted for age, sex, smoking, education and deprivation (index of multiple deprivation). ‘Everyone else’ control group includes either tested and SARS-CoV-2 (-) or those not tested. Sample sizes for SARS-CoV-2 (+) vs SARS-CoV-2 (-) were: N_Q1-all_ = 1 915; N_Q2-all_ = 1 744; N_Q1-females_ = 1 401; N_Q2-females_ = 1 265; N_Q1-males_ = 514; N_Q2-males_ = 479. Sample sizes for SARS-CoV-2 (+) vs everyone else were: N_all_ = 2 983; N_females_ = 2 020; N_males_ = 963.

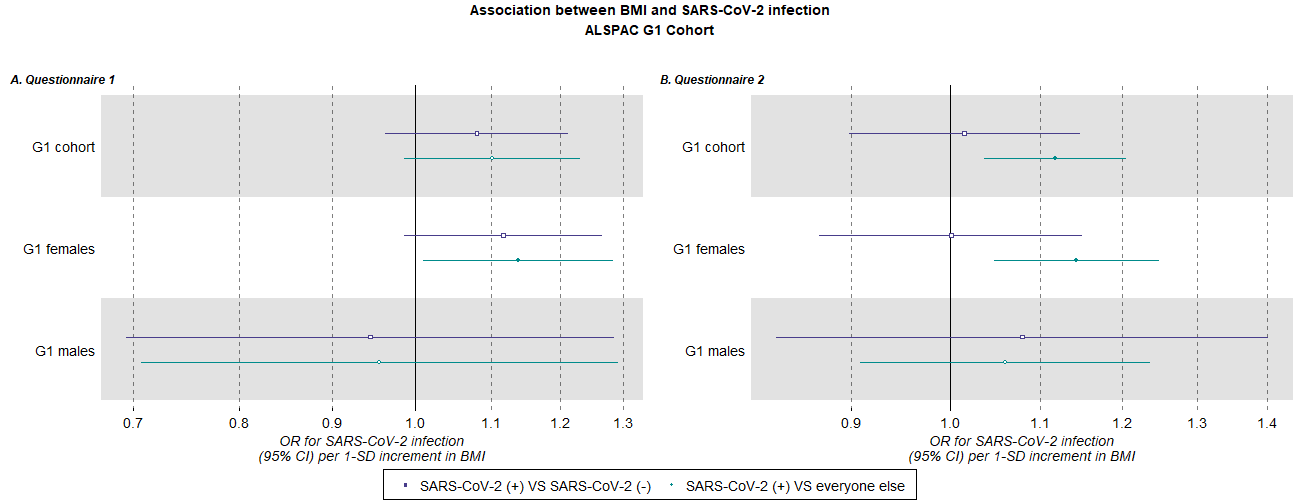

#### Supplementary Figure 6: Forest plots of the association between BMI and SARS-CoV-2 infection in the ALSPAC G0 Mothers cohort

ORs and their 95% confidence intervals are for questionnaire 1 (panel A) and questionnaire 2 (panel B). Models were adjusted for age, smoking, education and deprivation (index of multiple deprivation). ‘Everyone else’ control group includes those either tested and SARS-CoV-2 (-) or those not tested. Sample sizes for SARS-CoV-2 (+) vs SARS-CoV-2 (-) were: N_Q1_ = 1 929; N_Q2_ = 1 926. Sample size for SARS-CoV-2 (+) vs everyone else was: N = 4 261.

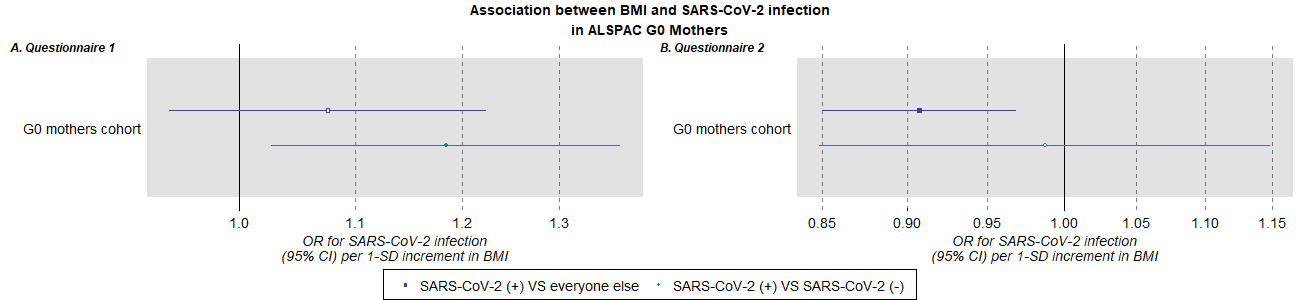

#### Supplementary Figure 7: Forest plots of the association between BMI and SARS-CoV-2 infection in UK Biobank

Estimates are shown separately for pre-mass testing (a) and post-mass testing (b)**.** ORs and their 95% confidence intervals are given for the whole sample and the subsamples stratified by sex. Models were adjusted for age, sex, smoking, education and deprivation (Townsend deprivation index). ‘Everyone else’ control group includes those either tested and SARS-CoV-2 (-) or those not tested. Sample sizes for SARS-CoV-2 (+) vs SARS-CoV-2 (-) were: N_pre-all_ = 4 662; N_post-all_ = 54 739; N_pre-females_ = 2 399; N_post-females_ = 29 372; N_pre-males_ = 2 263; N_post-males_ = 25 367. Sample sizes for SARS-CoV-2 (+) vs everyone else were: N_pre-all_ = 409 487; N_post-all_ = 407 768; N_pre-females_ = 225 876; N_post-females_ = 225 143; N_pre-males_ = 183 611; N_post-males_ = 182 625.

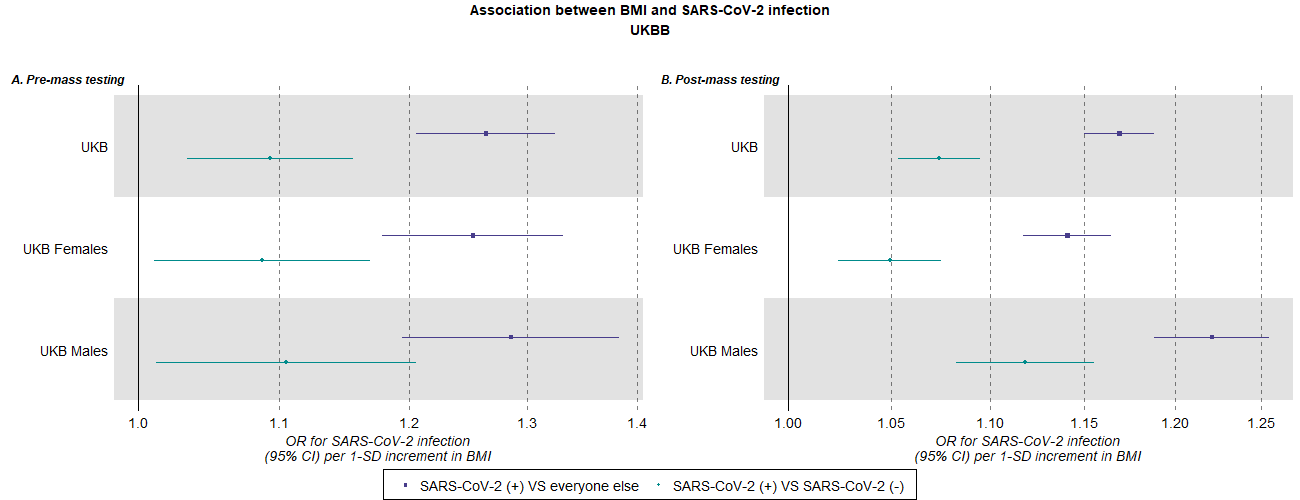

#### Supplementary Figure 8: Forest plots of the association between BMI and COVID-19 severity in UK Biobank

Estimates are shown separately for pre-mass testing (a) and post-mass testing (b). ORs and their 95% confidence intervals are given for the whole sample and the subsamples stratified by sex. Models were adjusted for age, sex, smoking, education and deprivation (Townsend deprivation index). ‘Everyone else’ control group includes those either tested and SARS-CoV-2 (-) or those not tested. Sample sizes for death-with-COVID-19 (“severe COVID-19”) vs SARS-CoV-2 (+) not resulting in death-with-COVID-19 (“non-severe COVID-19”) were: N_pre-all_ = 1 375; N_post-all_ = 12 533; N_pre-females_ = 644; N_post-females_ = 6 658; N_pre-males_ = 731; N_post-males_ = 5 875. Sample sizes for death-with-COVID-19 vs everyone: N_pre-all_ = 409 487; N_post-all_ = 407 768; N_pre-females_ = 225 845; N_post-females_ = 225 143; N_pre-males_ = 183 560; N_post-males_ = 182 625.

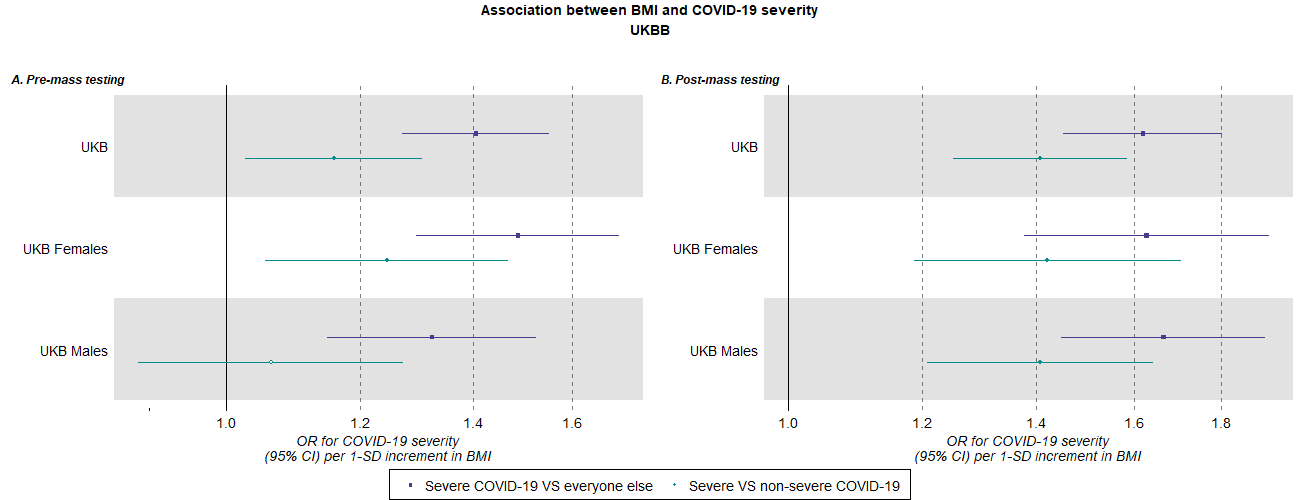

#### Supplementary figure 9: Simulated estimates of the effect of BMI on SARS-CoV-2 infection based on ALSPAC G1 cohort, questionnaire 1

|  | ***BMI, SARS-CoV-2 infection interaction for their effect on being tested*** | | |
| --- | --- | --- | --- |
| **Assumed effect of BMI on SARS-CoV-2 infection** | ***None*** | ***Plausible*** | ***Extreme*** |
| ***No effect*** | 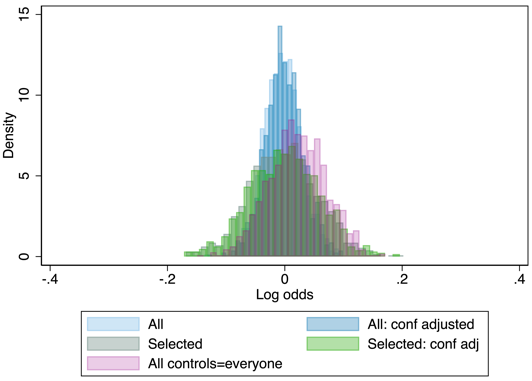 | 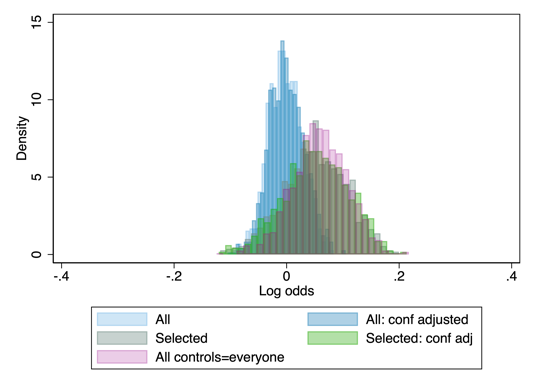 | 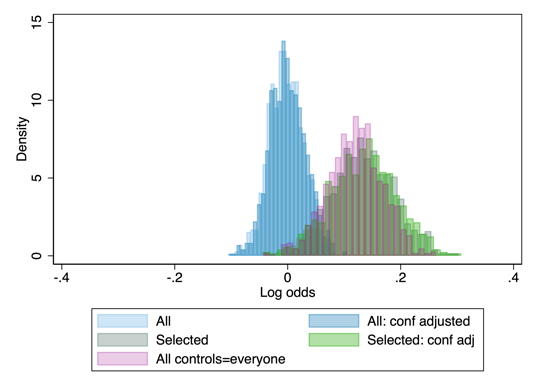 |
| ***OR=3 (log odds=1.1)*** | 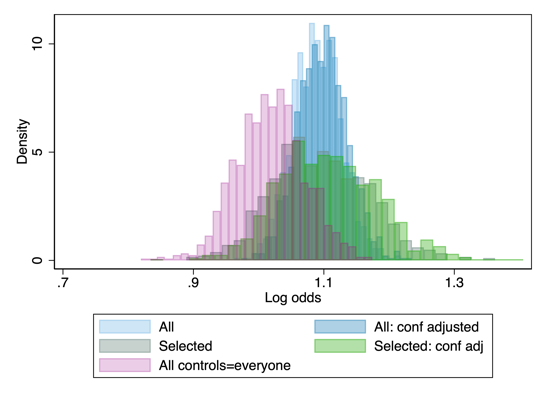 | 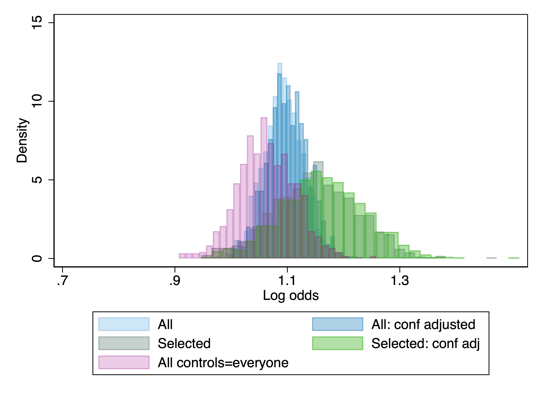 | 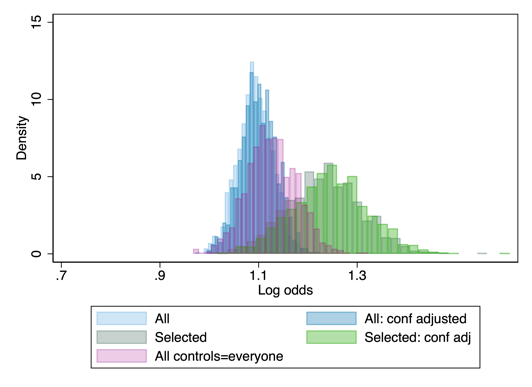 |

Estimate is the log odds of having a SARS-CoV-2 infection per 1 SD higher BMI.

Confounders: age, sex, education level, Townsend deprivation index and smoking status (never, former, current).

#### Supplementary figure 10: Illustration of bias directions in simulations

**a) Derivation of bias formula for binary case, for SARS-CoV-2 (+) versus SARS-CoV-2 (-) outcome definition**

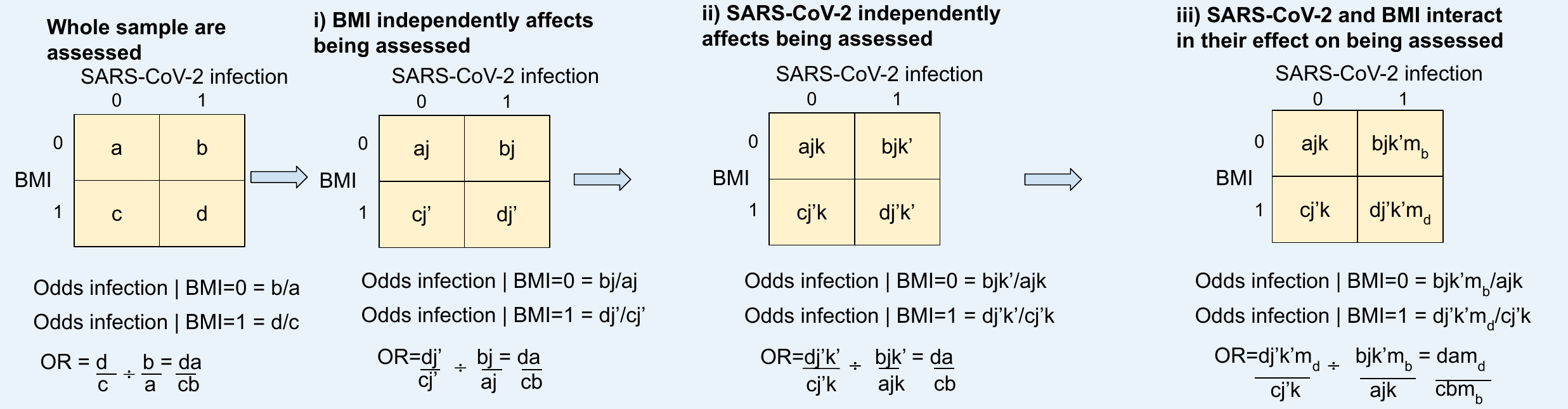
**b) Derivation of bias formula for binary case, for SARS-CoV-2 (+) versus everyone else outcome definition**

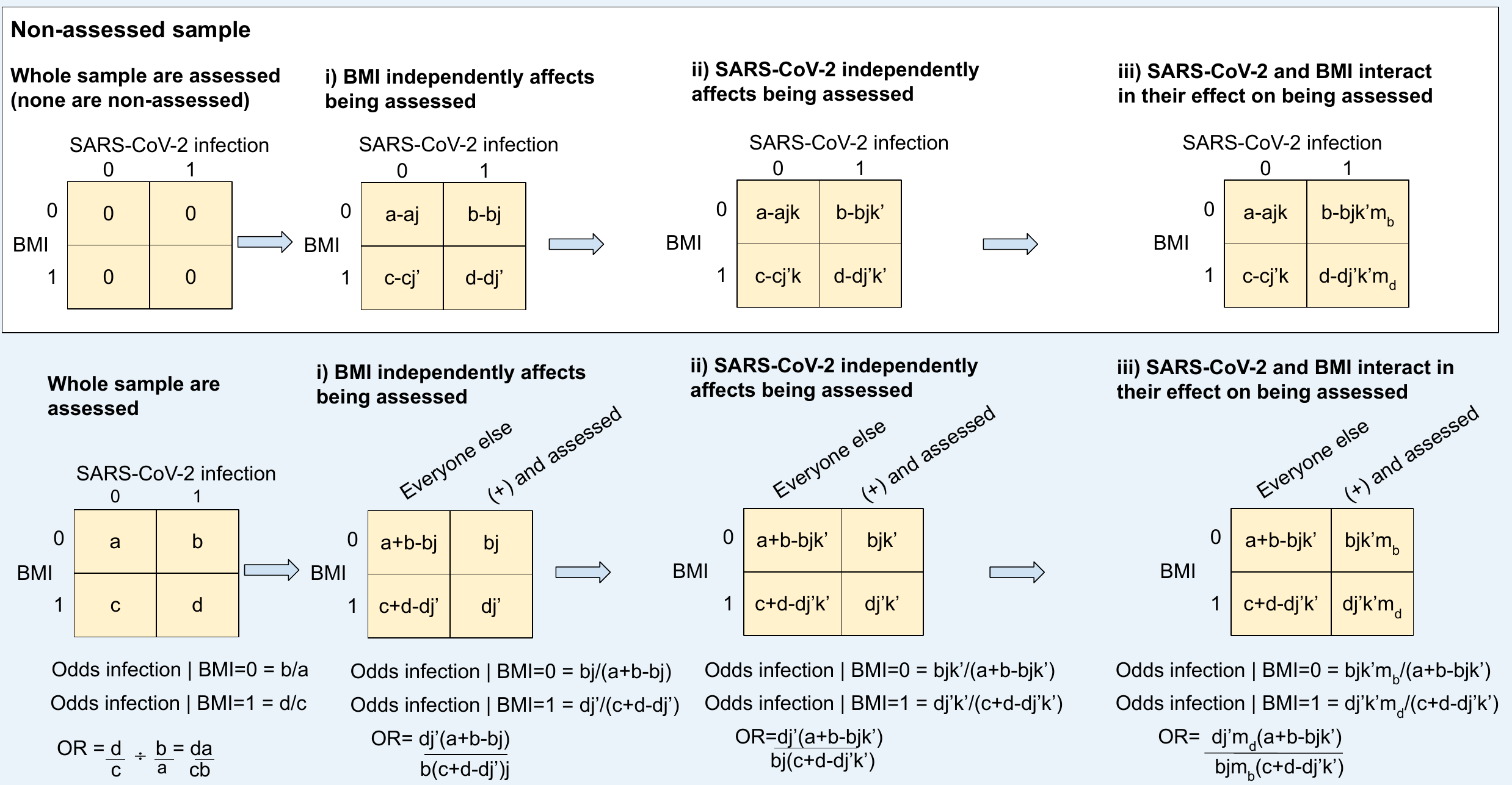
**c) Example scenarios for SARS-CoV-2 (+) versus SARS-CoV-2 (-) outcome definition**

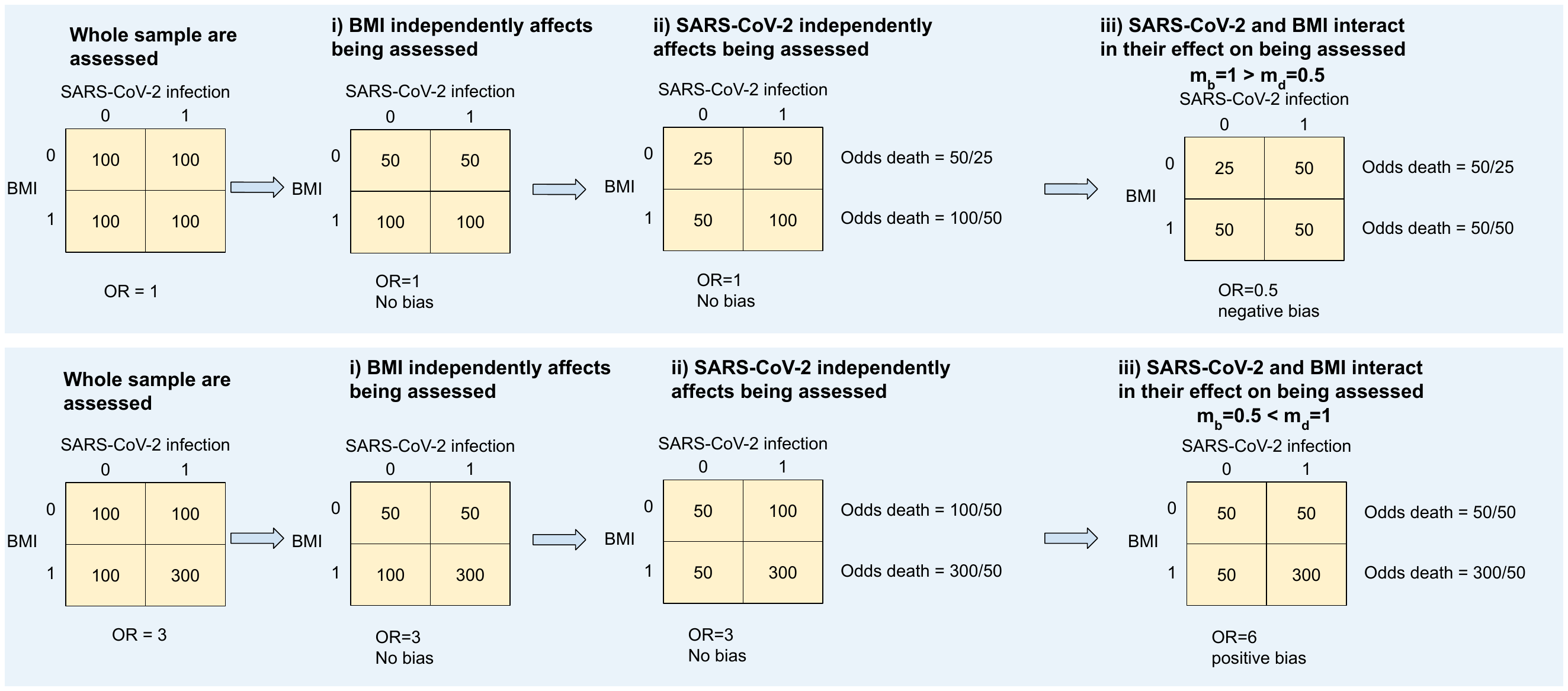

**d) Illustrating impact of parameter settings on bias direction**

**d.i) Example 1: Positive bias due to selection only, negative bias due to misclassification only, and positive bias when both selection and misclassification are at play**

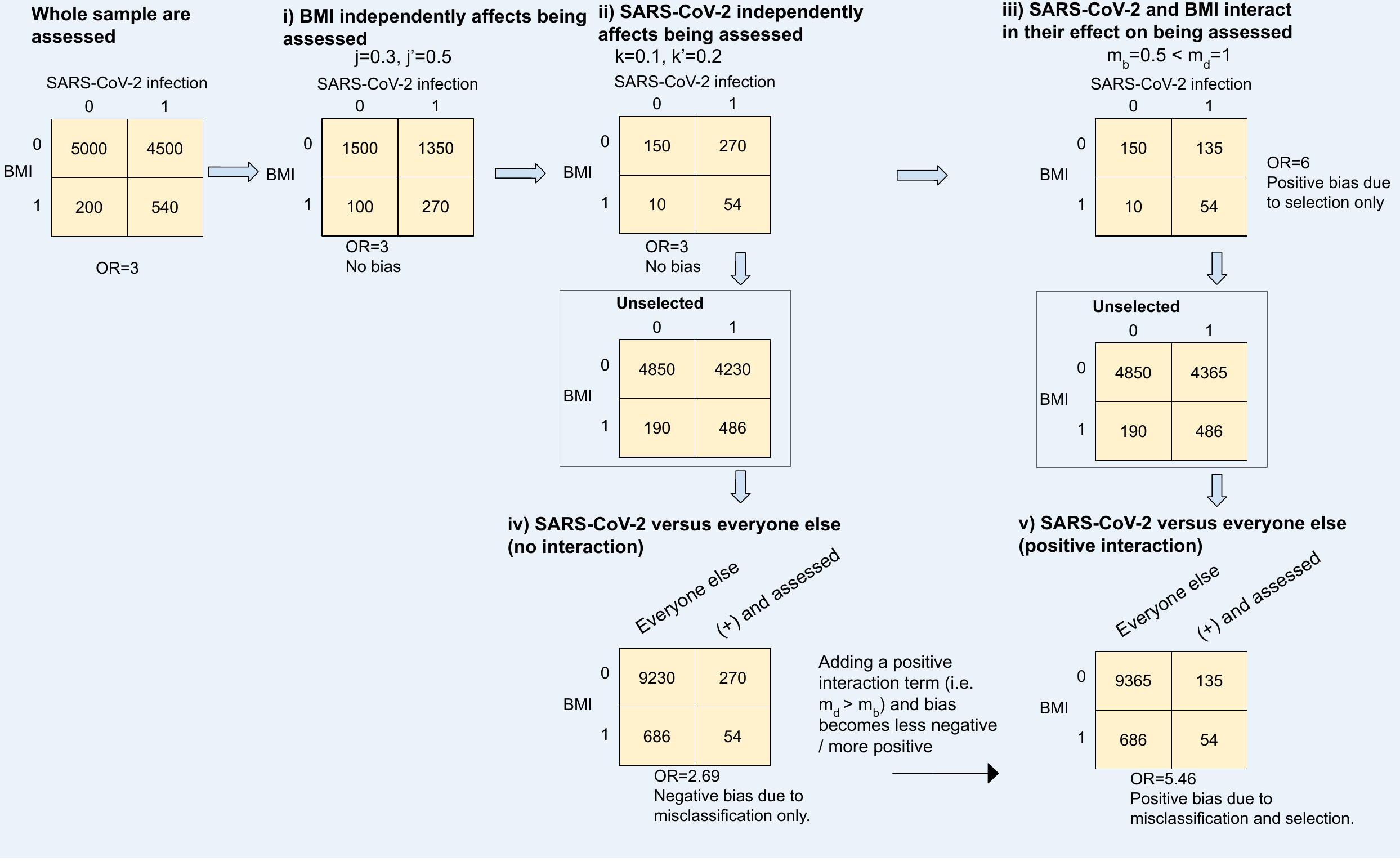

**d.ii) Example 2: Negative bias due to selection only, positive bias due to misclassification only, and negative bias when both selection and misclassification are at play**

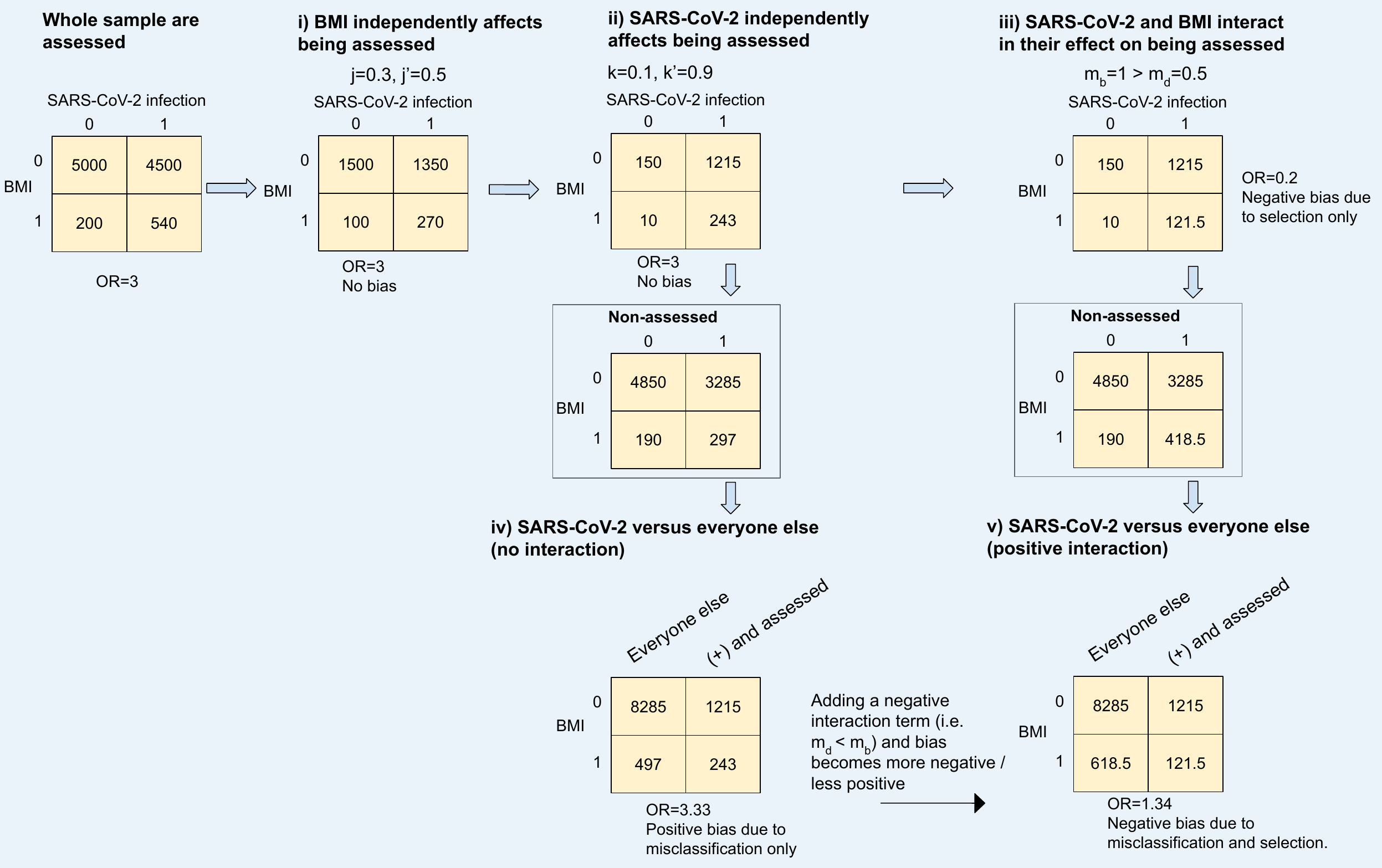

**e) Illustrating bias when all participants with a death-with-COVID-19 are selected into the analytical sample**

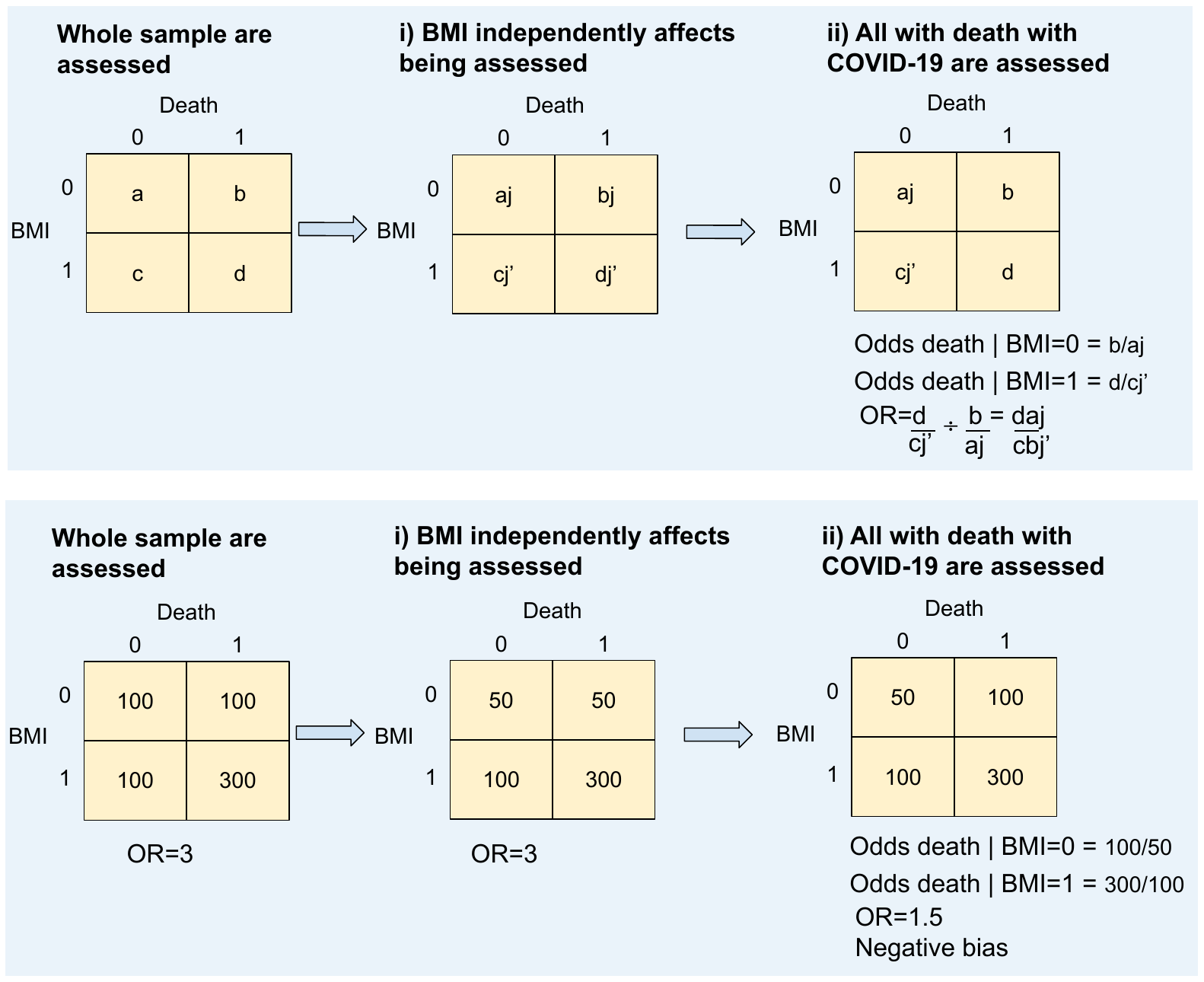

Illustrations assuming a binary exposure (BMI) and outcome (SARS-CoV-2 infection). Variables j and j’ denote the probability of selection when BMI=0 and BMI=1 respectively (i.e. the main effect of BMI in a poisson model). Variables k and k’ denote the probability of selection when SARS-CoV-2 infection = 0 and SARS-CoV-2 infection = 1 respectively (i.e. the main effect of infection in a poisson model). Variables m_b_ and m_d_ denote the probability of selection when BMI=0 and BMI=1, respectively, among the infected (i.e. representing the interaction term in a poisson model).

Illustration (a) demonstrates what can happen when BMI and SARS-CoV-2 infection affect selection, for the case vs control outcome definition. If BMI and infection independently affect selection on the probability scale ((i) and (ii)), then estimated OR will be unbiased. If BMI and infection interact in their effect on selection then, if the interaction term is positive (i.e. m_d_ > m_b_ in (iii)) bias will be positive (and vice versa if m_d_ < m_b_).

Illustration (b) demonstrates what can happen when BMI and SARS-CoV-2 infection affect selection, for the case vs everyone else outcome definition. If BMI and infection independently affect selection on the probability scale ((i) and (ii)), then bias occurs due to the reassignment of unselected cases as controls. If BMI and infection interact in their effect on selection then the total bias is a combination of bias due to misclassification, and bias due to this interaction (iii).

Illustration (c) demonstrates with a numeric example, bias for the case versus control outcome definition. Irrespective of effect of BMI on SARS-CoV-2 infection, if BMI and SARS-CoV-2 infection independently affect selection, estimates of effect with logistic regression will be unbiased.

Illustration (d) demonstrates with two numeric examples, bias for the SARS-CoV-2 (+) versus everyone else outcome definition. When there is misclassification bias but not selection bias (iv) bias can be positive or negative depending on the BMI distribution and the selection probability across this distribution. When there is additionally selection bias (v) total bias is a combination of bias due to misclassification and bias due to selection.

Illustration (e) demonstrates what can happen when all participants with a covid death are selected, such that statistically there is an interaction between death and BMI in their effects on selection, since the effect of BMI on selection depends on death-with-COVID-19. Since bias is proportional to j/j’ (i.e. one over the risk ratio of the independent effect of BMI on being assessed), if a higher BMI causes an increased probability of being assessed (as in our simulations) then the bias is negative (and vice versa if BMI causes a decreased probability of being assessed).

#### Supplementary figure 11: Simulated estimates of the effect of BMI on SARS-CoV-2 infection and COVID-19 severity based on UK Biobank, pre-mass testing

**a) Results of simulations estimating effect of BMI on SARS-CoV-2 infection**

|  | ***BMI, SARS-CoV-2 infection interaction for their effect on being tested*** | | |
| --- | --- | --- | --- |
| **Assumed effect of BMI on SARS-CoV-2 infection** | ***None*** | ***Plausible*** | ***Extreme*** |
| ***No effect*** | 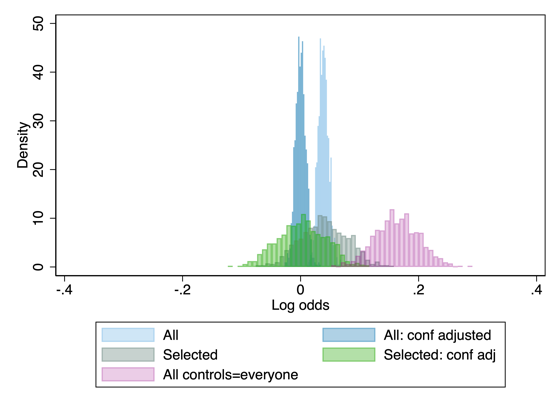 | 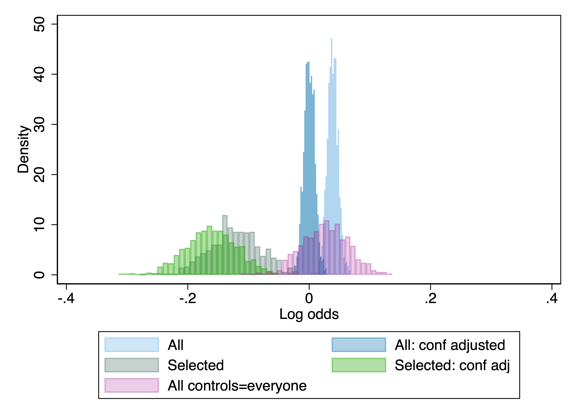 | 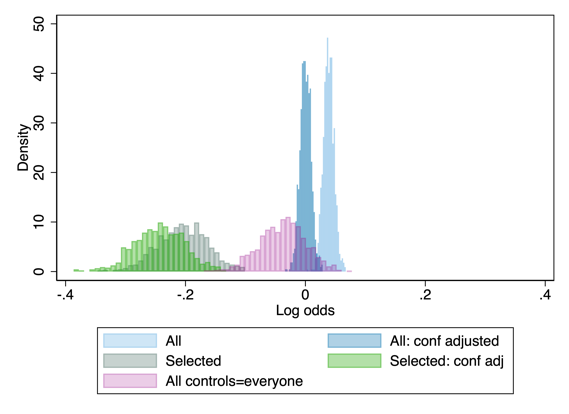 |
| ***OR=3 (log odds=1.1)*** | 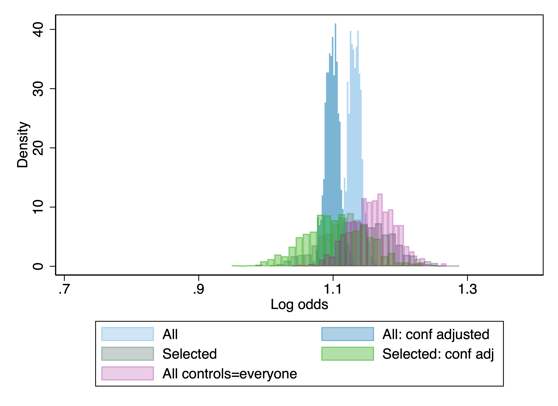 | 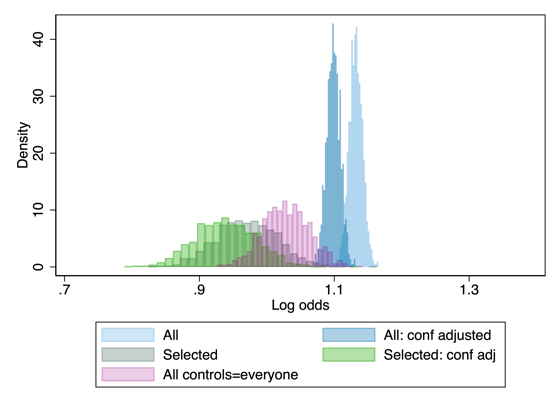 | 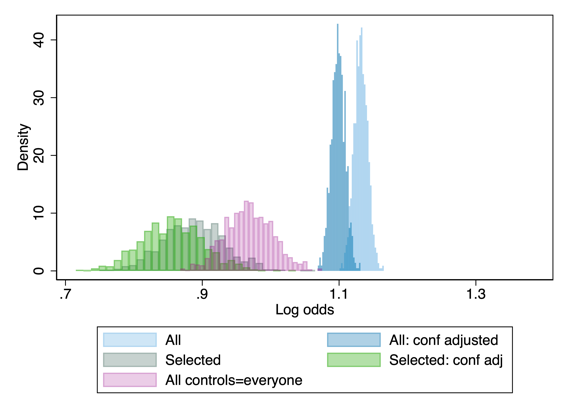 |

Estimate is the log odds of having a SARS-CoV-2 infection per 1 SD higher BMI.

Confounders: age, sex, education level, Townsend deprivation index and smoking status (never, former, current).

**b) Results of simulations estimating effect of BMI on COVID-19 severity**

|  | ***BMI, SARS-CoV-2 infection interaction for their effect on being tested*** | | |
| --- | --- | --- | --- |
| **Assumed effect of BMI on SARS-CoV-2 infection** | ***None*** | ***Plausible*** | ***Extreme*** |
| ***No effect*** | 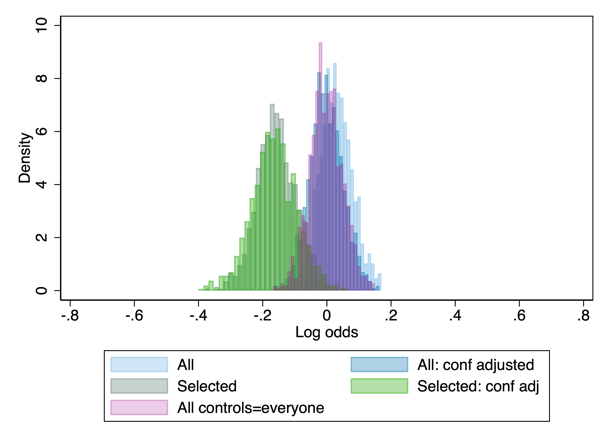 | 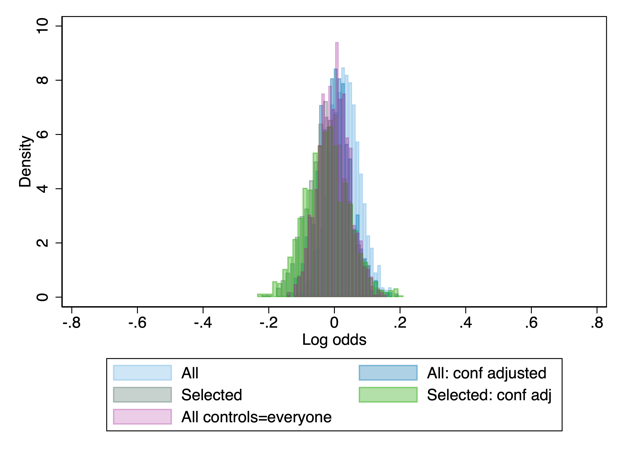 | 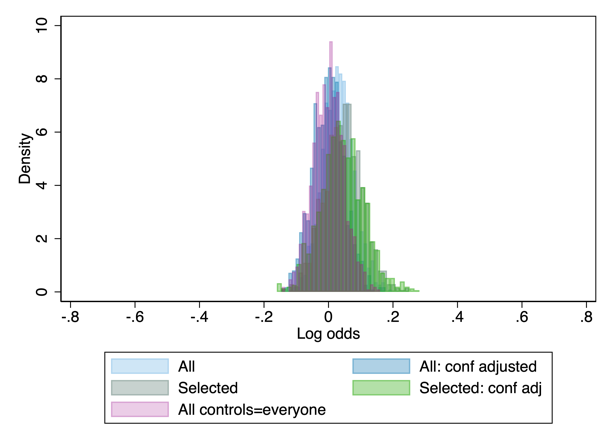 |
| ***OR=3 (log odds=1.1)*** | 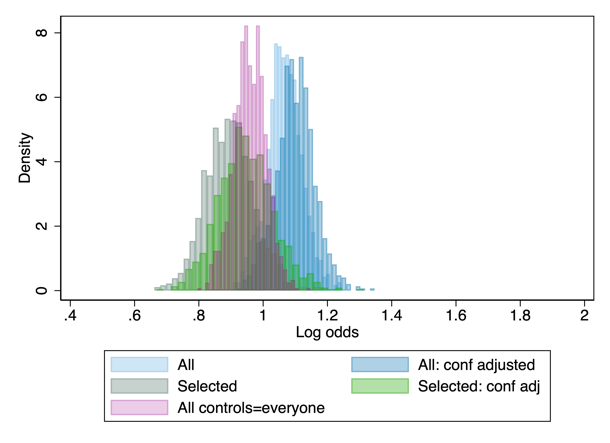 | 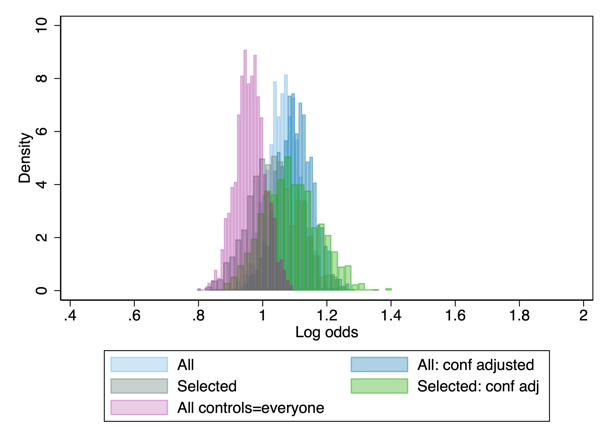 | 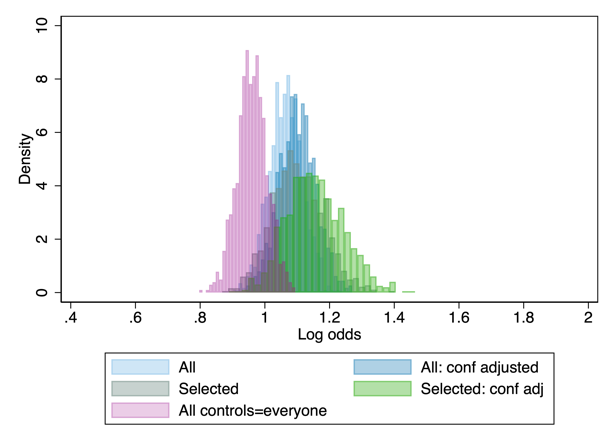 |

Estimate is the log odds of death-with-COVID-19 versus SARS-CoV-2 infection not resulting in death-with-COVID-19 per 1 SD higher BMI.

Confounders: age, sex, education level, Townsend deprivation index and smoking status (never, former, current).

All simulations include 1000 repetitions.

### REFERENCES

1.	Ward H, Atchison C, Whitaker M, Ainslie KEC, Elliott J, Okell L, et al. SARS-CoV-2 antibody prevalence in England following the first peak of the pandemic. Nat Commun. 2021;12: 905. doi:10.1038/s41467-021-21237-w

2.	Pollock AM, Lancaster J. Asymptomatic transmission of covid-19. BMJ. 2020;371: m4851. doi:10.1136/bmj.m4851

3.	Howe LJ, Lawson DJ, Davies NM, St. Pourcain B, Lewis SJ, Davey Smith G, et al. Genetic evidence for assortative mating on alcohol consumption in the UK Biobank. Nat Commun. 2019. doi:10.1038/s41467-019-12424-x
